# A multivariable Mendelian randomisation study of serum lipids and dementia risk within the UK Biobank

**DOI:** 10.1101/2024.05.27.24307728

**Authors:** Kitty Pham, Anwar Mulugeta, Amanda Lumsden, Elina Hyppӧnen

## Abstract

**Background and aims:** An unfavourable lipid profile has been associated with increased risk of dementia. However, it is challenging to investigate each serum lipid measure individually due to the high correlation between the traits. We tested for genetic evidence supporting associations between serum lipid measures and risk of dementia.

**Methods:** We conducted multivariable and univariable Mendelian randomisation (MR) analyses on 329,896 UK Biobank participants (age 37–73 years) to examine the associations between low-density lipoprotein cholesterol (LDL-C), high-density lipoprotein cholesterol (HDL-C), triglycerides, apolipoprotein-A1 (ApoA1) and apolipoprotein-B (ApoB), and the risk of dementia. The multivariable approach allows us to assess the association of each lipid measure with the outcome, including where the genetic variant-exposure associations are mediated by one another.

**Results:** In the univariable MR analyses, we observed no association between genetically determined serum lipids and risk of dementia. However, in a multivariable MR model containing LDL-C, triglycerides, and ApoB, ApoB was associated with a higher risk of dementia (OR per 1 SD higher ApoB 1.63, 95% CI 1.12, 2.37). Multivariable findings were consistent across IVWMR and MR-Egger, but not weighted median MR or MR-Lasso. HDL-C and ApoA1 were not associated with dementia in univariable or multivariable MR.

**Conclusions:** Our findings suggest that when considering the correlation between lipid measures, ApoB may play a role in the previously reported association between serum lipids and increased risk of dementia. Future studies should aim to confirm the findings in clinical/experimental studies and further explore the role of ApoB in dementia pathophysiology.

## Introduction

Lipids have been implicated in the pathology of dementia due to the role of lipid homeostasis dysregulation in disruption of the blood brain barrier, β-amyloid precursor protein (APP) processing, cellular remodelling, myelination, receptor-mediated signalling, inflammation, oxidative stress and the immune response [1]. Both observational and genetic studies largely agree that an adverse serum lipid profile is associated with increased risk of dementia, or specifically Alzheimer’s disease.

In the most recent cohort study, higher low-density lipoprotein cholesterol (LDL-C), measured in participants <65 years (n = 953,635), was reported to be associated with increased risk of dementia in later life [2]. Similarly, higher LDL-C, but not triglycerides or high-density lipoprotein cholesterol (HDL-C), was associated with incident dementia in a cohort study of participants >65 years [3]. However, there are some earlier studies that report contradictory findings. A Mendelian randomisation (MR) study of multiple disease factors found no association between LDL-C, HDL-C or triglycerides and dementia [4]. Meanwhile, two studies in cohorts of participants aged >50 years found that higher LDL-C was inversely associated with dementia risk [5], and lower LDL-C levels were associated with slower cognitive decline [6]. However, these studies had notably smaller sample sizes of *n*=3,467 and *n*=7,129, respectively, than the most recent cohort study. In another study conducted using a Japanese cohort, lower HDL-C, also considered an adverse lipid measurement, was associated with greater cognitive impairment (*n*=1,114) and dementia in later life (*n*=781) [7]. Meta-analysis of genetic studies supported this observational study, suggesting that the association between lower HDL-C and Alzheimer’s disease may be causal [8].

The evidence on the relationship between apolipoproteins and risk of dementia is less clear. Since apolipoproteins are bound to both LDL-C and HDL-C, which have opposing relationships with dementia, some prior studies have attempted to differentiate between the effects of apolipoprotein subtypes and other lipid traits [9]. A study of apolipoprotein B (ApoB) within the cerebrospinal fluid (CSF) found that higher ApoB concentration in the CSF was associated with higher T-tau and P-tau, which are considered markers of neurodegeneration and Alzheimer’s disease [10, 11]. In a study of serum ApoB however, higher ApoB was associated with protective markers for Alzheimer’s disease. Studies on serum apolipoprotein A1 (ApoA1) reported that lower ApoA1 was a risk factor for cognitive decline and Alzheimer’s disease [12, 13].

Mendelian randomisation (MR) analysis uses genetic variants that are associated with an exposure of interest to assess genetic evidence for a causal effect on a disease or phenotype outcome [14]. Since genetic variants are randomly assigned at conception, this method is less susceptible to misleading inferences caused by reverse causation and bias from confounding factors. Multivariable MR (MVMR) allows the concepts of conventional MR to be applied in cases with pleiotropic variants [15]. MVMR additionally assumes that each genetic variant may be associated with one or more exposures and that the association of one genetic variant with the outcome may be mediated by another exposure.

The high correlation between lipid traits raises challenges for the investigation of the independent relationships between each serum lipid and dementia, including for the selection of appropriate genetic instruments in genetic studies [16]. Differentiating between the effects of different lipids can be important in understanding how each directly affects the outcome of interest, and hence in better understanding their pathophysiological relevance. A prior multivariable MR investigating causal effects of serum lipids on cardiovascular disease risk highlighted ApoB as the main driver, showing that its association with cardiovascular disease was stronger than that seen for other atherogenic lipid traits. We sought to understand whether similar patterns are seen for brain health, which is often closely linked to cardiovascular health [17].

In this study, we tested for genetic evidence supporting causal associations between genetically instrumented serum lipid profile (LDL-C, HDL-C, triglycerides, ApoA1 and ApoB) and risk of dementia, using a multivariable MR approach.

## Methods

### Study populations

Our study uses exposure data from the UK Biobank and summary outcome data from the Lambert et al. meta-analysis of four Alzheimer’s GWAS studies [18]. We included up to 329 896 individuals from the UK Biobank database and up to 54,162 individuals (17,008 cases and 37,154 controls) from the Lambert et al. GWAS.

The UK Biobank is a population-based prospective cohort comprised of over 500,000 participants aged 37 – 73 years [19]. As a secondary analysis of UK Biobank data, this study relies on the consents of subjects at their participation with the UK Biobank data collection studies [20]. Ethics approval of the UK Biobank data is managed by the National Information Governance Board for Health and Social Care and North West Multicentre Research Ethics Committee (11/NW/0382). Participants of the UK Biobank study have provided informed consent for use of their anonymised data and have the right to withdraw at any point without explanation or penalty. Access and use of the database have been granted to researchers of this study under UK Biobank application number 10171.

### Serum lipids

All serum lipids traits including LDL-C (measured as LDL direct), HDL-C, triglycerides, ApoA1 and ApoB, were measured from blood samples taken from the participants at baseline assessment according to a standard protocol [21, 22]. Samples from aliquot 4 (*n*= 673) were excluded due to severe dilution issues. Values underwent rank-based inverse normal transformation, with a mean of zero and standard deviation of one. Full details on the sample collection and processing protocols have been reported elsewhere [21].

### Dementia risk

For phenotypic analyses, we used data from the UK Biobank. Dementia cases were collected using primary care data, hospital admission records, national death registers and self-reported touchscreen questionnaire answers [23]. For genetic analyses, we used summary data from the International Genomics of Alzheimer’s Project (IGAP) GWAS by Lambert and colleagues [18], which was performed as a meta-analysis of four large consortia totalling 74,046 participants: 17,008 cases and 37,154 controls, including no overlap with the UK Biobank. Dementia cases satisfied the DSM-IV (Diagnostic and Statistical Manual of Mental Disorders, 4^th^ edition) or NINCDS-ADRDA (National Institute of Neurological and Communicative Diseases and Stroke/Alzheimer’s Disease and Related Disorders Association) criteria for Alzheimer’s disease, confirmed by clinical assessment or autopsy. Controls were free of dementia, including diagnoses of mild cognitive impairment.

### Genetic instruments for serum lipids

The discovery and selection of genetic instruments to proxy the five lipid exposures have been reported by Richardson and colleagues [16]. The full SNP list includes novel SNPs identified from a genome wide association study within the UK Biobank and SNPs previously reported in the Global Lipids Genetics Consortium [24] (for LDL-C, HDL-C and triglycerides) and Kettunen and colleagues [25] (for ApoA1 and ApoB). Selected SNPs were associated with the exposure at a significance threshold of *P<*5×10^-8^ and linkage disequilibrium of r^2^<0.001.

We excluded 38 LDL-C SNPs, 96 HDL-C SNPs, 68 triglyceride SNPs, 88 ApoA1 SNPs and 58 ApoB SNPs which were not available within the IGAP dementia GWAS (Supplementary Table 1). We additionally excluded 7 tri-allelic SNPs and 28 palindromic SNPs during the data harmonisation process. Given that *APOE* genotype is a very strong determinant of dementia risk [26] and also affects traits that are influential in the lipid-dementia pathways (e.g. BMI), we excluded SNPs from the *APOE* gene and neighbouring gene regions (*NECTIN2*, *APOC1, BCL3*) in the main analyses [27]. This allowed us to measure the effects of lipid traits when excluding the strong influence of *APOE*. To account for pleiotropic effects, we also checked for any remaining SNPs associated with dementia at a threshold of *P*<5×10^-8^ within the IGAP GWAS. In the main analyses we included 173 SNPs for LDL-C, 423 SNPs for HDL-C, 361 SNPs for triglycerides, 338 SNPs for ApoA1 and 193 SNPs for ApoB (Figure 1).

**Figure 1:**
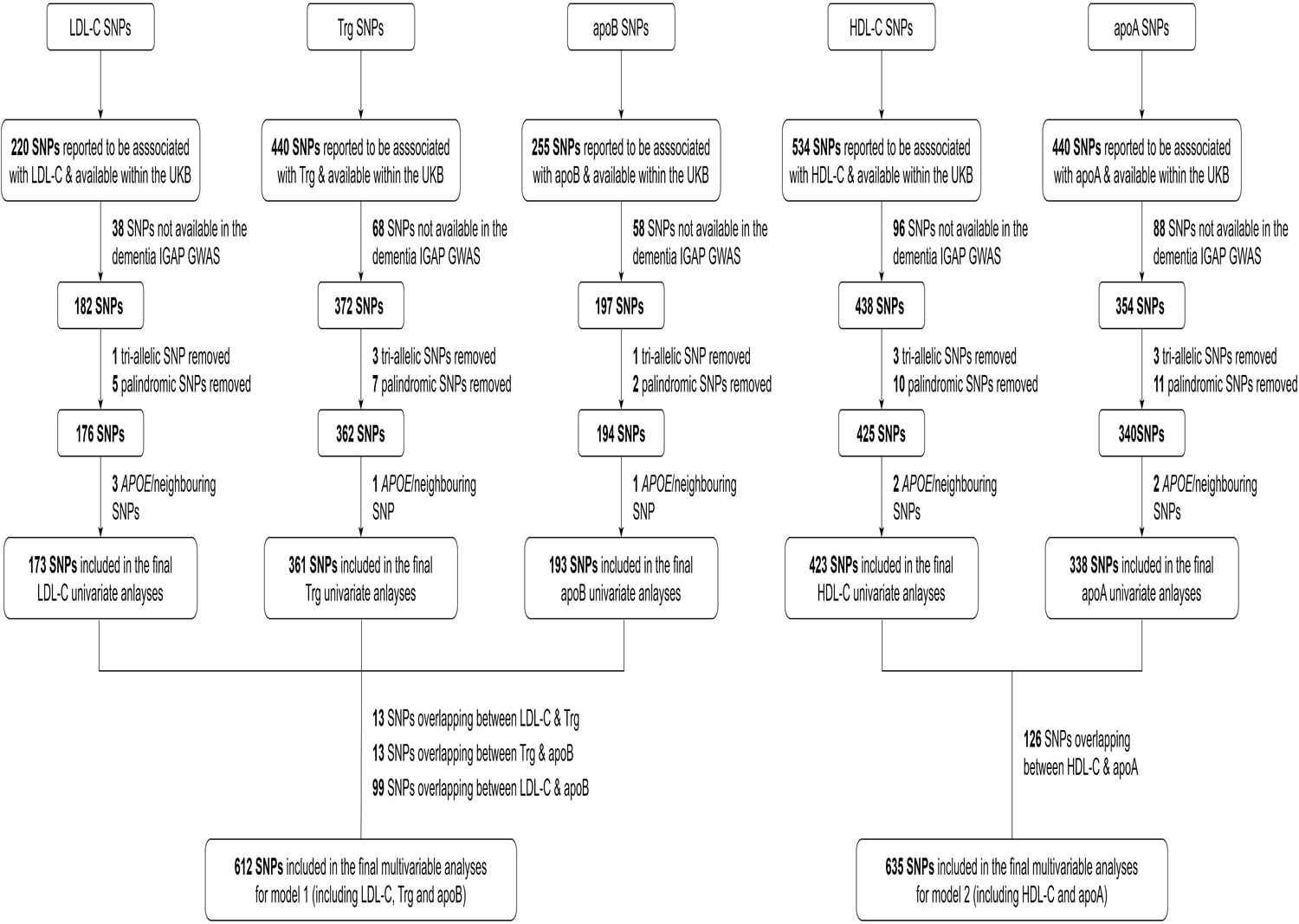
Flowchart showing the SNP selection process for instrumental variables of the five lipid traits (LDL-C, HDL-C, triglycerides, ApoA1 and ApoB), and SNP overlap between lipid traits in each multivariable MR model.

### Statistical analyses

Firstly, we conducted phenotypic analyses using lipid measures from the UK Biobank. We tabulated the baseline characteristics of our study population and showed the correlations of each lipid trait with one another in a correlation matrix. The phenotypic associations of the lipid traits and dementia incidence were tested with Cox proportional hazard models, adjusted for age, sex, UK Biobank assessment centre, fasting time before blood collection and lipid-lowering medication use.

We conducted univariable and multivariable two-sample MR analyses using the TwoSampleMR [28], MR-PRESSO [29] and MendelianRandomization [30] R packages. SNP-exposure estimates were collected from a sample of 329,896 unrelated white British individuals within the UK Biobank, by regressing the exposure against the selected SNPs, adjusting for age, sex, SNP array and 40 principal components. SNP-exposure estimates for each lipid trait were calculated using individual models. The full list of SNPs used for each lipid trait are provided in Supplementary Tables 2 – 6. As described above, summary data from the IGAP GWAS was used for the SNP-outcome estimates. SNP outcome-estimates were adjusted for age, sex, and principal components. Univariate MR was used to assess genetic evidence for a causal association between each lipid trait and the risk of dementia. Meanwhile, multivariable MR allowed us to assess these associations, while adjusting for the mediating effect of other traits included in the model [31]. The lipid traits were split into two models containing correlated lipid traits for multivariable analyses: the first included LDL-C, triglycerides and ApoB (model 1); and the other with HDL-C and ApoA1 (model 2) [16]. After accounting for the overlap of SNPs between lipid traits in the same model, the main analyses included 612 SNPs in model 1 and 635 SNPs in model 2. Univariable analyses used inverse variance weighted MR (IVWMR), MR-Egger, weighted median, and weighted mode MR methods, while multivariable analyses were conducted with IVWMR, MR-Egger, weighted median MR, and MR-Lasso (MR methods available in the MendelianRandomization R package).

The different MR methods allow us to assess the presence of horizontal pleiotropy, which may indicate that the observed effect is through a non-direct pathway [15]. IVWMR assumes that there is no pleiotropy or balanced horizontal pleiotropy in the genetic instruments. MR-Egger relaxes the pleiotropic assumptions and allows pleiotropic instruments if the InSIDE assumption is met (the InSIDE assumption states that the associations of the genetic instruments on the exposure must be independent from any pleiotropic effects of the genetic instruments on the outcome) [32]. Weighted median assumes that a majority of the genetic instruments are valid (no horizontal pleiotropy) and weighted mode assumes that the largest cluster of genetic instruments is valid [33, 34]. The MR-Lasso method applies lasso-type penalisation and performs IVWMR using only genetic instruments deemed valid in the lasso procedure (which shrinks the regression coefficients towards zero, forcing some coefficients to be zero) [35].

As sensitivity analyses, we repeated the main analyses accounting for fasting time and history of lipid-lowering medication use (in separate models) by including fasting time and medication use as a covariate when collecting the SNP-exposure estimates. We also repeated the multivariable analyses for model 1 adjusting for the effects of traits in model 2 (HDL-C, ApoA1) and for model 2 adjusting for the effects of traits in model 1 (LDL-C, triglycerides, ApoB). To test associations when including the effects of *APOE*, we repeated the multivariable analyses including SNPs in *APOE* and neighbouring gene regions (which were excluded from the main analyses). Finally, multivariable analyses were replicated using summary data from the IGAP GWAS conducted by Kunkle and colleagues [36], which is an extension of the IGAP GWAS used in the main analyses. All analyses were reported according to the STROBE-MR guidelines.

## Results

Our full analysis sample was comprised of 329,896 individuals from the UK Biobank, of whom 53.4% were female and 75.1% self-reported above average health (Table 1). Concentration of all serum lipid traits were higher in older age groups, while participants with higher BMI groups had higher LDL-C, triglycerides and ApoB, but lower HDL-C and ApoA1 levels. Participants with below average self-reported health and those using lipid-lowering medication tended to have higher triglyceride values, but lower LDL-C, HDL-C, ApoA1 and ApoB. Longer fasting times before blood sample collection tended to occur in conjunction with higher LDL-C and ApoB measurements, and lower HDL-C, triglycerides and ApoA1. As expected, dementia incidence within the population was the highest in older age groups and participants with poorer self-reported health.

**Table 1:** Distribution of average lipid traits (LDL-C, HDL-C, triglycerides, ApoA1 and ApoB) across population characteristics, within our analysis sub-sample of the UK Biobank.

|  | *n* (%) | LDL cholesterol |  | HDL cholesterol |  | Triglycerides |  | Apolipoprotein A1 |  | Apolipoprotein B |  | Dementia |  |
| --- | --- | --- | --- | --- | --- | --- | --- | --- | --- | --- | --- | --- | --- |
|  |  | Median (IQR), mmol/L | *P* value | Median (IQR), mmol/L | *P* value | Median (IQR), mmol/L | *P* value | Median (IQR), mmol/L | *P* value | Median (IQR), mmol/L | *P* value | Incidence rate^ (n) | *P* value |
| **Total population** | 329,896 | 3.53 (1.17) |  | 1.40 (0.51) |  | 1.50 (1.11) |  | 1.52 (0.35) |  | 1.02 (0.32) |  | 7.8 (3,117) |  |
| **Sex** |  |  | <1 x 10<sup>-300</sup> |  | <1 x 10<sup>-300</sup> |  | <1 x 10<sup>-300</sup> |  | <1 x 10<sup>-300</sup> |  | 1.4 x 10<sup>-49</sup> |  | 5.8 x 10<sup>-33</sup> |
| Women | 176,172 (53.4) | 3.59 (1.17) |  | 1.56 (0.50) |  | 1.34 (0.93) |  | 1.62 (0.34) |  | 1.02 (0.32) |  | 6.1 (1,302) |  |
| Men | 153,724 (46.6) | 3.47 (1.17) |  | 1.24 (0.39) |  | 1.70 (1.27) |  | 1.41 (0.29) |  | 1.02 (0.32) |  | 9.6 (1,815) |  |
| **Age (in years)** |  |  | 9.2 x 10<sup>-67</sup> |  | 1.5 x 10<sup>-82</sup> |  | 5.2 x 10<sup>-236</sup> |  | <1 x 10<sup>-300</sup> |  | 2.8 x 10<sup>-147</sup> |  | 6.1 x 10<sup>-272</sup> |
| 39 – 49 years | 72,005 (21.8) | 3.39 (1.06) |  | 1.37 (0.47) |  | 1.32 (1.10) |  | 1.47 (0.33) |  | 0.98 (0.30) |  | 1.2 (131) |  |
| 50 – 59 years | 125,346 (38.0) | 3.62 (1.13) |  | 1.42 (0.52) |  | 1.50 (1.11) |  | 1.52 (0.35) |  | 1.04 (0.31) |  | 3.7 (621) |  |
| 60 – 73 years | 132,545 (40.2) | 3.53 (1.26) |  | 1.41 (0.51) |  | 1.58 (1.07) |  | 1.53 (0.35) |  | 1.02 (0.33) |  | 15.3 (2,365) |  |
| **BMI (in kg/m<sup>2</sup>)** |  |  | <1 x 10<sup>-300</sup> |  | <1 x 10<sup>-300</sup> |  | <1 x 10<sup>-300</sup> |  | <1 x 10<sup>-300</sup> |  | <1 x 10<sup>-300</sup> |  | 1.6 x 10<sup>-9</sup> |
| Underweight, <18.5 kg/m<sup>2</sup> | 1,632 (0.5) | 3.20 (1.06) |  | 1.79 (0.56) |  | 0.94 (0.47) |  | 1.72 (0.39) |  | 0.91 (0.27) |  | 8.5 (20) |  |
| Normal, [18.5, 25) kg/m<sup>2</sup> | 107,003 (32.5) | 3.44 (1.09) |  | 1.59 (0.52) |  | 1.12 (0.75) |  | 1.62 (0.36) |  | 0.98 (0.29) |  | 6.7 (875) |  |
| Overweight, [25, 30) kg/m<sup>2</sup> | 140,635 (42.6) | 3.61 (1.18) |  | 1.37 (0.46) |  | 1.59 (1.11) |  | 1.49 (0.33) |  | 1.04 (0.32) |  | 7.4 (1,262) |  |
| Obese, ≥30 kg/m<sup>2</sup> | 79,555 (24.1) | 3.53 (1.25) |  | 1.24 (0.40) |  | 1.90 (1.27) |  | 1.42 (0.31) |  | 1.04 (0.34) |  | 9.5 (916) |  |
| Missing | 1,071 (0.3) | 3.38 (1.29) |  | 1.32 (0.52) |  | 1.62 (1.13) |  | 1.45 (0.36) |  | 1.00 (0.34) |  | 31.9 (44) |  |
| **Self-reported general health** |  |  | <1 x 10<sup>-300</sup> |  | <1 x 10<sup>-300</sup> |  | <1 x 10<sup>-300</sup> |  | <1 x 10<sup>-300</sup> |  | 7.5 x 10<sup>-109</sup> |  | 1.5 x 10<sup>-169</sup> |
| Excellent | 55,150 (16.7) | 3.56 (1.09) |  | 1.51 (0.51) |  | 1.29 (0.91) |  | 1.57 (0.35) |  | 1.02 (0.30) |  | 3.6 (241) |  |
| Good | 192,690 (58.4) | 3.57 (1.15) |  | 1.42 (0.50) |  | 1.47 (1.07) |  | 1.53 (0.35) |  | 1.03 (0.31) |  | 6.0 (1,365) |  |
| Fair | 67,207 (20.4) | 3.44 (1.27) |  | 1.30 (0.46) |  | 1.69 (1.24) |  | 1.46 (0.34) |  | 1.01 (0.34) |  | 12.1 (1,004) |  |
| Poor | 13,696 (4.2) | 3.27 (1.33) |  | 1.23 (0.46) |  | 1.81 (1.38) |  | 1.41 (0.35) |  | 0.98 (0.35) |  | 27.1 (477) |  |
| Missing | 1,153 (0.3) | 3.41 (1.28) |  | 1.32 (0.47) |  | 1.68 (1.21) |  | 1.47 (0.34) |  | 0.99 (0.34) |  | 20.5 (30) |  |
| **Fasting time before sample collection (in hours)** |  |  | 1.6 x 10<sup>-54</sup> |  | 4.2 x 10<sup>-5</sup> |  | 5.1 x 10<sup>-116</sup> |  | 0.0003 |  | 3.6 x 10<sup>-95</sup> |  | 1.4 x 10<sup>-8</sup> |
| <8 hours | 319,070 (96.7) | 3.53 (1.17) |  | 1.40 (0.51) |  | 1.50 (1.11) |  | 1.52 (0.35) |  | 1.02 (0.32) |  | 7.7 (2,990) |  |
| 8 – 12 hours | 4,104 (1.2) | 3.61 (1.20) |  | 1.36 (0.49) |  | 1.30 (0.96) |  | 1.48 (0.35) |  | 1.05 (0.35) |  | 7.4 (39) |  |
| >12 hours | 6,707 (2.0) | 3.65 (1.25) |  | 1.37 (0.50) |  | 1.30 (0.99) |  | 1.49 (0.37) |  | 1.07 (0.34) |  | 11.1 (88) |  |
| Missing | 15 (0.0001) | 3.67 (1.06) |  | 1.37 (0.93) |  | 2.10 (0.94) |  | 1.53 (0.47) |  | 1.00 (0.22) |  | 0.0 (0) |  |
| **Lipid-lowering medication** |  |  | <1 x 10<sup>-300</sup> |  | <1 x 10<sup>-300</sup> |  | 6.3 x 10<sup>-183</sup> |  | <1 x 10<sup>-300</sup> |  | <1 x 10<sup>-300</sup> |  | 6.1 x 10<sup>-25</sup> |
| No | 273,778 (83.0) | 3.69 (1.08) |  | 1.43 (0.51) |  | 1.46 (1.07) |  | 1.53 (0.35) |  | 1.06 (0.30) |  | 6.0 (1,999) |  |
| Yes | 56,116 (17.0) | 2.72 (0.84) |  | 1.25 (0.44) |  | 1.70 (1.21) |  | 1.46 (0.34) |  | 0.85 (0.24) |  | 16.5 (1,118) |  |
| Missing | 2 (0.00001) | 2 (0.72) |  | 1.08 (0.01) |  | 2.46 (0.56) |  | 1.38 (0.23) |  | 1.18 (0.31) |  | 0.0 (0) |  |

IQR: interquartile range
^ per 10,000 person-years.

In phenotypic analyses, we saw the expected strong correlations between LDL-C and ApoB, and between HDL-C and ApoA1 (Supplementary table 7). Analyses between the lipid traits and dementia showed protective associations for LDL-C, HDL-C and ApoA1 with dementia (Table 2).

**Table 2:** Phenotypic associations of each lipid trait (LDL-C, HDL-C, triglycerides, ApoA1 and ApoB) with incident dementia risk, using Cox proportional hazard models.

|  | **Cases** | **Controls** | **HR (95% CI)** | **P value** |
| --- | --- | --- | --- | --- |
| **LDL-cholesterol** | 2,634 | 311,387 | 0.92 (0.87, 0.97) | 0.002 |
| **HDL-cholesterol** | 2,459 | 292,014 | 0.88 (0.78, 0.99) | 0.032 |
| **Triglycerides** | 2,638 | 311,694 | 1.01 (0.97, 1.05) | 0.73 |
| **Apolipoprotein A1** | 2,433 | 290,421 | 0.62 (0.52, 0.73) | 2.0 x 10<sup>-8</sup> |
| **Apolipoprotein B** | 2,622 | 310,438 | 0.87 (0.72, 1.04) | 0.13 |

All analyses adjusted for age, sex, assessment centre, fasting time before blood sample collection, lipid lowering medication use.

### Association of genetically determined serum lipids with dementia

Our univariate MR analyses showed no associations between any lipid traits and dementia (Figure 2). In the multivariable MR analyses, in a model containing LDL-C, triglycerides and ApoB, we observed an association between ApoB and increased risk of dementia (OR per 1 SD higher ApoB 1.63, 95% CI 1.12, 2.37), while LDL-C had a protective effect (OR 0.63, 95% CI 0.43, 0.94). Multivariable analyses did not reveal any significant associations for HDL-C and ApoA1 with dementia. Estimates were consistent between all MR methods in the univariate analyses. In the multivariable analyses, estimates were broadly similar between IVWMR and MR-Egger, but not consistent with weighted median MR or MR-Lasso. Analyses conducted in the Kunkle et al. GWAS were consistent with our main analyses in the Lambert et al. GWAS (Supplementary Figure 1).

**Figure 2:**
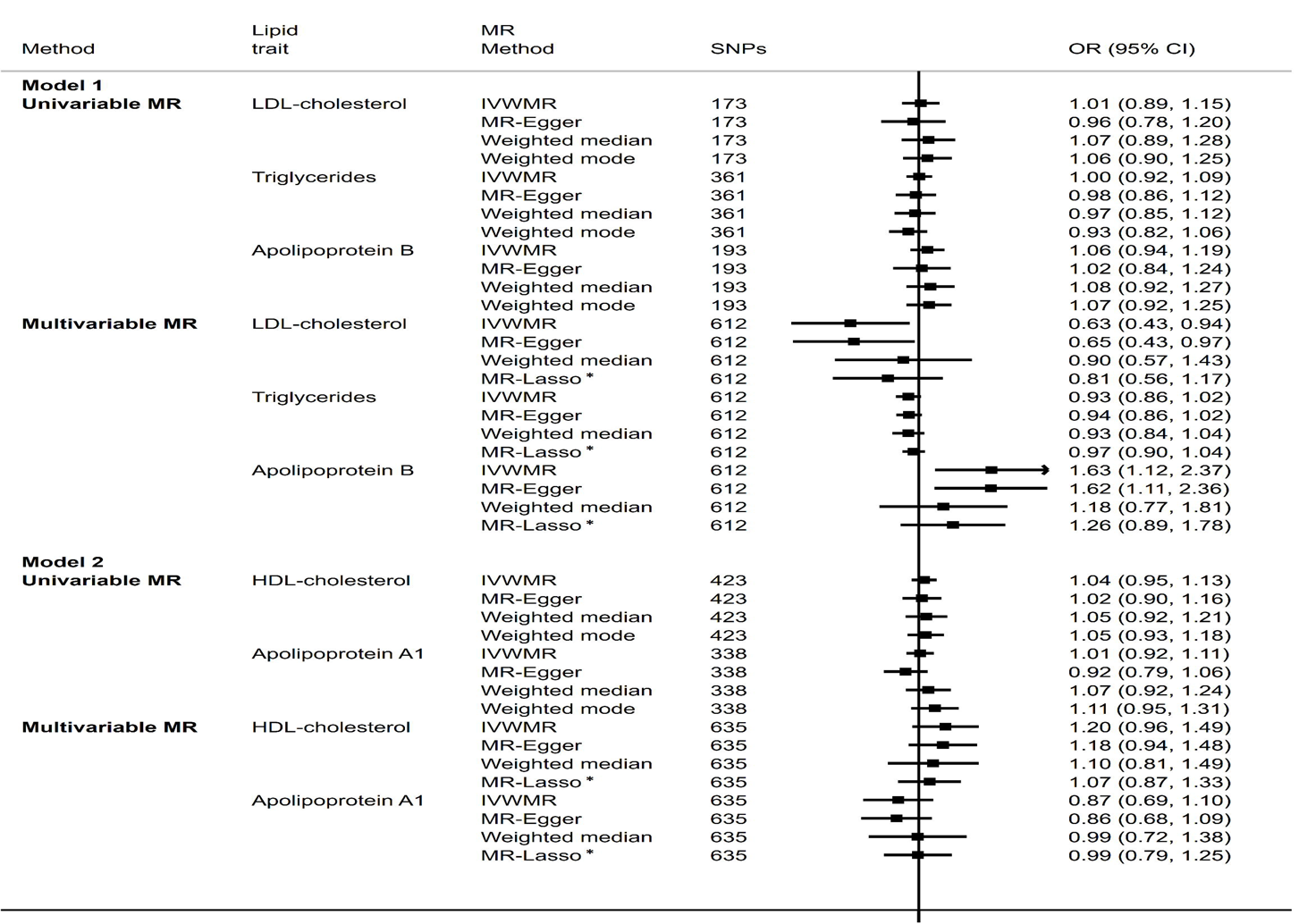
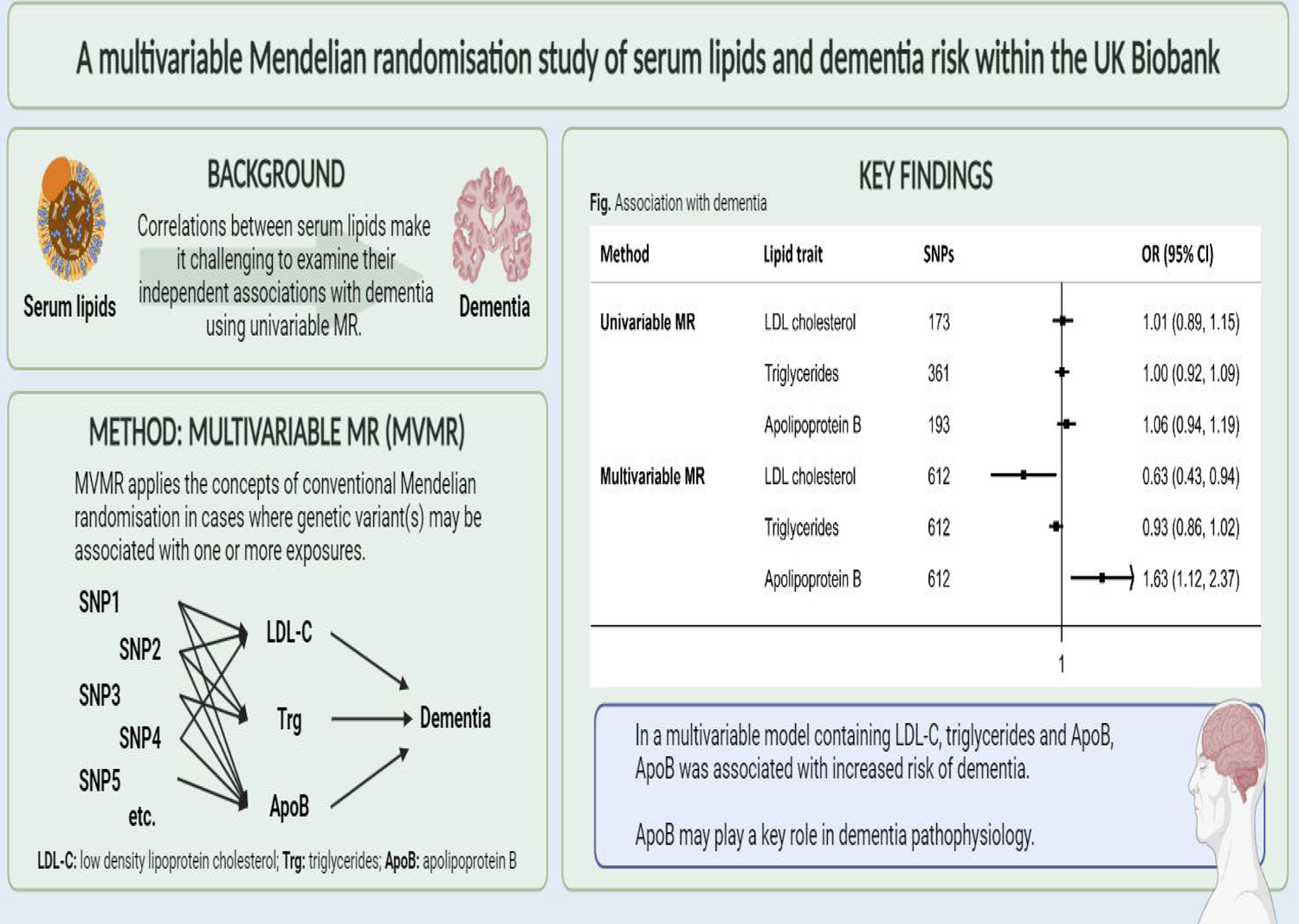
Forest plot showing the associations between genetically instrumented lipid traits and dementia risk, using univariable and multivariable MR, for model 1 (LDL-C, triglycerides and ApoB) and model 2 (HDL-C and ApoA1). Estimates are shown for IVWMR, MR-Egger, weighted median, weighted mode, and MR-PRESSO for univariable MR, and for IVWMR, MR-Egger, weighted median, and MR-Lasso for multivariable MR. MR-Egger p-intercept for model 1 >0.57. MR-Egger p-intercept for model 2 >0.10. * 581/612 SNPs in model 1 were considered valid instruments for the multivariable MR-Lasso analyses 599/635 SNPs in model 2 were considered valid instruments for the multivariable MR-Lasso analyses.

### Sensitivity analyses

In sensitivity analyses additionally adjusting for fasting time (before blood sample collection) and history of lipid-lowering medication use provided similar estimates to the main results (Supplementary Figure 2).

When accounting for alternative pathways through lipid traits in model 2 (HDL-C, ApoA1) in the model 1 multivariable analyses and for lipid traits in model 1 (LDL-C, triglycerides, ApoB) in the model 2 multivariable analyses, estimates were almost identical to the main results (Supplementary Figure 3).

Multivariable MR analyses including the *APOE*-related SNPs provided directionally consistent estimates, but of stronger magnitude (Supplementary Figure 4). Estimates for model 1 were directionally consistent with the main results, while IVWMR and MR-Egger estimates for model 2 suggested that when controlling for the effects of one another, higher HDL-C is associated with higher dementia risk and higher ApoA1 is protective.

## Discussion

Correlations between different lipid traits make it challenging to examine their independent associations with disease risk. In our study despite the phenotypic analyses suggesting inverse associations with dementia for LDL-C, HDL-C and ApoB, there was no evidence for an association between any of the genetically determined lipid traits and dementia in univariate MR analyses. In contrast, in the multivariable analyses which accounted for intercorrelations, a higher ApoB was associated with an increased risk of dementia, while higher LDL-C had an inverse association. This suggests that ApoB may play a role in the previously reported association between serum lipids and increased risk of dementia. Our findings are compatible with a previous study that used similar multivariable MR methodology to estimate the direct effects between serum lipids and cardiovascular disease [16], with both studies suggesting that that ApoB may at least partly explain some of the adverse associations observed for higher LDL-C.

The association between ApoB and dementia may occur through the same pathway as cardiovascular disease, which is a known risk factor of dementia [37]. ApoB has been implicated in the formation of atherosclerotic plaques [38]. ApoB is a protein found on chylomicrons, very-low-density lipoproteins (VLDL), intermediate-density lipoproteins (IDL) and low-density lipoproteins (LDL) that carry fat and cholesterol to, and within, the bloodstream [39]. ApoB binds to LDLR receptor on cells, allowing LDLs to enter the cell and release cholesterol, which is used in various cellular processes. However, LDLR is downregulated when cells have sufficient cholesterol, and excess LDLs in the bloodstream undergo oxidation and get taken up by scavenger cells, which can lead to the formation of foam cells; an early step of atherosclerosis [40]. While this is a plausible pathway for the effect between ApoB and dementia, the effects do not appear to be relevant for two rare conditions affecting the *APOB* gene, abetalipoproteinemia and homozygous familial hypercholesterolemia (HoFH) [41, 42]. Both conditions cause deficiencies in ApoB but are known to increase neurological deterioration (abetalipoproteinemia) and atherosclerotic cardiovascular disease (HoFH), which opposes the direction of association reported in our study.

Our findings support previous evidence which suggests divergent relationships for LDL-C and ApoB on disease risk. Discordance analyses conducted in a multicentre cohort study of young adults found that risk of coronary artery calcification was more strongly influenced by ApoB, than by LDL-C or non-HDL-C [43].

Another study also found that elevated LDL-C, but normal ApoB was associated with lower risk of carotid atherosclerosis [44]. In our analyses, some of the multivariable MR methods suggested an association between higher LDL-C and lower risk of dementia, when accounting for the effects of ApoB and excluding the effects of *APOE*. This could be as LDL-C is known to have antioxidant properties and may relieve oxidative stress within the system, as seen in studies on type 2 diabetes [45, 46] and in animal studies on dementia [47, 48]. An alternative explanation is that the protective association seen between LDL-C and dementia is reflective of LDL-C particle size. When controlling for ApoB, higher LDL-C levels may represent larger LDL-C particle size, which are thought to be less atherogenic than smaller particles and less likely to penetrate into the arterial walls [49]. In an earlier cohort study also within the UK Biobank, higher LDL/ApoB ratio was found to be associated with decreased risk of all-cause dementia [50].

A major strength of our study is the availability of a large sample of phenotypic serum lipid data and individual-level genetic data within the UK Biobank. The multivariable MR analysis method allows us to investigate the individual effects of highly correlated serum lipid traits without introducing collider bias [31]. To our knowledge, this is the first study to explore the direct causal associations between both lipoprotein cholesterols and apolipoproteins with the risk of dementia, using multivariable MR. However, there are some limitations of our study which should be discussed. Firstly, the analyses were conducted in white British populations so the findings cannot be directly generalised to other ethnic populations. Selection bias could be present given that the disease outcome was assessed in an elderly population (survivor bias) and as the outcome population included prevalent cases of dementia whereby cases with relatively milder forms of dementia may have been overrepresented (prevalence-incidence bias). The summary statistics used for the outcome associations are from one of the largest publicly available GWAS meta-analyses with no overlap with the UK Biobank. However, the use of summary-level data limits our ability to adjust for certain covariates or conduct stratified analyses. While we conducted analyses to test for pleiotropic effects and excluded known pleiotropic SNPs, we are unable to account for the effects of residual confounding and cannot completely exclude bias caused by pleiotropy. Furthermore, the weighted median and MR-Lasso estimates were not consistent with the IVWMR and MR-Egger methods in the multivariable analyses so we cannot discount the potential bias from directional pleiotropy [51]. Further investigation is required to understand how each method performs when applied to a group of highly correlated exposures. Multivariable MR is designed to estimate linear associations, so we may be unable to accurately describe any non-linear relationships. In addition, MR relies on genetic instruments to proxy the effect of our exposures, which only represents the average effects of each serum lipid, not the complex changes that may occur throughout an individual’s lifetime. Our estimates reflect a situation where the lipids are controlled for each other, therefore these findings should not be taken to discount the utility of LDL-C as a biomarker for dementia.

In summary, our study suggests a potentially harmful effect of higher ApoB on dementia. It supports the existing body of knowledge on ApoB and cardiovascular disease, where ApoB has been suggested as a more accurate biomarker for the prediction of atherogenic risk than LDL-C. The genetic evidence for a strong role of ApoB in dementia risk suggested by this study should be confirmed and further explored in clinical/experimental studies.

## Author contributions

KP conceptualized the study with EH, analysed the data and prepared the first draft. AM and EH advised on data analyses and supervised the study with AL. KP, AM, AL and EH interpreted results, revised the paper and approved the manuscript for submission.

## Ethical approval

Ethics approval for the UK Biobank was granted by the National Information Governance Board for Health and Social Care and North West Multi-centre Research Ethics Committee (11/NW/0382). All researchers have gained approval for use of the database under UK Biobank application number 10171.

## Use of AI and AI-assisted Technologies Statement

AI has not been used in the writing process.

## URLS

**UK Biobank data access:** https://www.ukbiobank.ac.uk/enable-your-research/apply-for-access

**Dementia GWAS database:** https://gwas.mrcieu.ac.uk/datasets/ieu-a-297/

**PLINK software:** https://zzz.bwh.harvard.edu/plink/dataman.shtml

**R MVMR package:** https://cran.r-project.org/web/packages/MendelianRandomization/vignettes/Vignette_MR.pdf

## Data Availability

All data supporting this study will be available to approved users of the UK Biobank upon application.

https://www.ukbiobank.ac.uk/enable-your-research/apply-for-access

