## Supplementary for "A multivariable Mendelian randomisation study of serum lipids and dementia risk within the UK Biobank"

### Supplemental files:

#### List of Tables:

**Supplementary table 1:** List of excluded SNPs (tri-allelic, palindromic, and pleiotropic SNPs) for each lipid trait. 2

**Supplementary table 2**: List of 173 SNPs used as instrumental variables for LDL-C, and their association with LDL-C and dementia. 3 – 5

**Supplementary table 3:** List of 423 SNPs used as instrumental variables for HDL-C, and their association with HDL-C and dementia. 6 – 12

**Supplementary table 4:** List of 361 SNPs used as instrumental variables for triglycerides, and their association with triglycerides and dementia. 13 – 18

**Supplementary table 5:** List of 338 SNPs used as instrumental variables for ApoA, and their association with ApoA and dementia. 19 – 24

**Supplementary table 6:** List of 193 SNPs used as instrumental variables for ApoB, and their association with ApoB and dementia. 25 – 28

**Supplementary table 7:** Phenotypic correlations between the lipid traits (LDL-C, HDL-C, triglycerides, ApoA and ApoB). 29

#### List of Figures:

**Supplementary figure 1:** Forest plot showing the associations between genetically instrumented lipid traits and dementia risk, using the Kunkle et al. 2019 IGAP GWAS. Estimates are shown for multivariable MR, using the IWVMR method. 30

**Supplementary figure 2:** Forest plot showing the associations between genetically instrumented lipid traits and dementia risk, adjusting for fasting time before blood sample collection or adjusting for history of lipid-lowering medication use. Estimates are shown for univariable and multivariable MR, using the IWVMR method. 31

**Supplementary figure 3:** Forest plot showing the associations between genetically instrumented lipid traits and dementia risk. Model 1 uses SNPs associated with LDL-C, triglycerides and ApoB (while controlling for pathways through HDL-C and ApoA) and model 2 uses SNPs associated with HDL-C and ApoA (while controlling for pathways through LDL-C, triglycerides and ApoB). Estimates are shown for multivariable MR using IVWMR, MR-Egger, weighted median, and MR-Lasso. 32

**Supplementary Figure 4:** Forest plot showing the associations between genetically instrumented lipid traits and dementia risk, including SNPs in *APOE* and neighbouring genes (model 1: rs6857, rs4452060, rs1551891, rs483082, rs12691088; model 2: rs429358, rs2965169). Estimates are shown for multivariable MR using IVWMR, MR-Egger, weighted median, and MR-Lasso. 33

**Supplementary table 1:** List of excluded SNPs (tri-allelic, pleiotropic, and palindromic SNPs) for each lipid trait.

| | **LDL-C SNPs** | **HDL-C SNPs** | **Trg SNPs** | **apoA SNPs** | **apoB SNPs** | **Total SNPs excluded** |
|---|---|---|---|---|---|---|
| **Tri-allelic SNPs** | *rs9832727* | *rs34555420, rs1047964, rs1601934* | *rs4858894, rs9832727, rs9553567* | *rs34555420, rs1047964, rs2069945* | *rs9832727* | 7 |
| **Palindromic SNPs** | *rs990619, rs28849176, rs4782568, rs4808360, rs516316* | *rs11118320, rs2705616, rs74419430, rs10901802, rs2204886, rs62112763, rs12976395, rs407133, rs133015, rs1894544* | *rs1861435, rs6722159, rs11722924, rs2705616, rs6916318, rs4500049, rs10822163* | *rs59038405, rs12044156, rs115660648, rs989075, rs10036789, rs74419430, rs7835108, rs2204886, rs3760230, rs12976395, rs133015* | *rs4782568, rs516316* | 28 |
| **Pleiotropic SNPs** | *rs6857 (NECTIN2), rs4452060 (NECTIN2), rs1551891** | *rs429358 (APOE), rs2965169** | *rs483082 (APOC1)* | *rs429358 (APOE), rs2965169** | *rs12691088 (APOC1)* | 7 |

* Associated with dementia within the IGAP dementia GWAS at a significance threshold of $P<5x10^{-8}$.

**Supplementary table 2:** List of 173 SNPs used as instrumental variables for LDL-C, and their association with LDL-C and dementia.

| CHR | rsID | POS | Effect allele | Other allele | EAF | BETA LDL-C | SE LDL-C | Pval LDL-C | n LDL-C | BETA dementia | SE dementia | Pval dementia | n dementia |
|---|---|---|---|---|---|---|---|---|---|---|---|---|---|
| 1 | rs880315 | 10796866 | T | C | 0.66 | 0.015 | 0.003 | 3.9E-08 | 304433 | 0.008 | 0.017 | 0.63 | 54162 |
| 1 | rs12078100 | 16512586 | G | C | 0.63 | 0.012 | 0.003 | 7.4E-06 | 310568 | 0.012 | 0.016 | 0.45 | 54162 |
| 1 | rs61775180 | 25793663 | C | T | 0.58 | 0.026 | 0.003 | 5.1E-24 | 309739 | 0.012 | 0.018 | 0.49 | 54162 |
| 1 | rs472495 | 55521313 | T | G | 0.65 | 0.042 | 0.003 | 1.6E-56 | 313346 | -0.046 | 0.018 | 0.01 | 54162 |
| 1 | rs11206517 | 55526428 | G | T | 0.03 | 0.065 | 0.007 | 7.6E-20 | 313840 | 0.019 | 0.043 | 0.66 | 54162 |
| 1 | rs11206788 | 56797981 | C | G | 0.60 | 0.012 | 0.003 | 5.7E-06 | 285380 | -0.005 | 0.017 | 0.77 | 54162 |
| 1 | rs1556562 | 93034023 | T | G | 0.79 | 0.019 | 0.003 | 1.1E-10 | 314021 | -0.031 | 0.020 | 0.12 | 54162 |
| 1 | rs6693893 | 109797763 | T | C | 0.97 | 0.078 | 0.007 | 1.4E-29 | 313683 | -0.009 | 0.045 | 0.84 | 54162 |
| 1 | rs4970834 | 109814880 | C | T | 0.82 | 0.107 | 0.003 | 4.6E-238 | 309456 | 0.006 | 0.021 | 0.79 | 54162 |
| 1 | rs115458560 | 110061974 | T | C | 0.98 | 0.046 | 0.009 | 9.5E-07 | 314021 | -0.043 | 0.072 | 0.55 | 54162 |
| 1 | rs6667939 | 198994619 | T | C | 0.72 | 0.018 | 0.003 | 5.2E-10 | 307137 | 0.000 | 0.018 | 0.99 | 54162 |
| 1 | rs2642438 | 220970028 | G | A | 0.70 | 0.024 | 0.003 | 6.6E-19 | 314021 | 0.006 | 0.019 | 0.77 | 54162 |
| 1 | rs10910476 | 234734956 | T | C | 0.56 | 0.013 | 0.003 | 2.9E-07 | 307617 | 0.013 | 0.018 | 0.45 | 54162 |
| 1 | rs556107 | 234853059 | T | C | 0.52 | 0.036 | 0.003 | 3.1E-46 | 310569 | 0.017 | 0.016 | 0.28 | 54162 |
| 1 | rs28631087 | 235109214 | T | C | 0.79 | 0.016 | 0.003 | 1.9E-07 | 313350 | -0.005 | 0.024 | 0.82 | 54162 |
| 2 | rs56236159 | 3636478 | G | T | 0.13 | 0.017 | 0.004 | 4.7E-06 | 313305 | 0.003 | 0.025 | 0.91 | 54162 |
| 2 | rs72774870 | 11512825 | C | T | 0.93 | 0.018 | 0.005 | 1.8E-04 | 313052 | 0.011 | 0.032 | 0.73 | 54162 |
| 2 | rs907866 | 20371380 | G | A | 0.55 | 0.018 | 0.003 | 5.7E-12 | 310594 | 0.024 | 0.016 | 0.13 | 54162 |
| 2 | rs934197 | 21267461 | A | G | 0.34 | 0.083 | 0.003 | 3.8E-214 | 313231 | 0.005 | 0.018 | 0.76 | 54162 |
| 2 | rs1731243 | 26930411 | T | C | 0.61 | 0.012 | 0.003 | 4.6E-06 | 311581 | -0.030 | 0.017 | 0.07 | 54162 |
| 2 | rs1260326 | 27730940 | T | C | 0.39 | 0.036 | 0.003 | 9.0E-44 | 314021 | -0.001 | 0.016 | 0.96 | 54162 |
| 2 | rs4299376 | 44072576 | G | T | 0.32 | 0.051 | 0.003 | 7.6E-82 | 313155 | -0.018 | 0.017 | 0.30 | 54162 |
| 2 | rs6709904 | 44080324 | A | G | 0.89 | 0.039 | 0.004 | 2.5E-22 | 313357 | 0.037 | 0.024 | 0.13 | 54162 |
| 2 | rs7562734 | 63047973 | G | C | 0.68 | 0.019 | 0.003 | 8.6E-12 | 310852 | 0.003 | 0.016 | 0.86 | 54162 |
| 2 | rs12471768 | 64928603 | C | T | 0.71 | 0.016 | 0.003 | 7.0E-09 | 312447 | 0.045 | 0.018 | 0.01 | 54162 |
| 2 | rs2718717 | 109206139 | G | A | 0.14 | 0.020 | 0.004 | 1.2E-08 | 312723 | -0.036 | 0.022 | 0.11 | 54162 |
| 2 | rs150474434 | 118845121 | G | A | 0.90 | 0.036 | 0.004 | 1.2E-17 | 310331 | 0.046 | 0.027 | 0.09 | 54162 |
| 2 | rs17050272 | 121306440 | G | A | 0.59 | 0.021 | 0.003 | 3.1E-16 | 314021 | -0.005 | 0.017 | 0.76 | 54162 |
| 2 | rs4954192 | 135632981 | T | C | 0.36 | 0.015 | 0.003 | 6.5E-09 | 313866 | 0.037 | 0.016 | 0.02 | 54162 |
| 2 | rs6714750 | 136783169 | G | A | 0.18 | 0.014 | 0.003 | 2.5E-05 | 300038 | 0.019 | 0.019 | 0.33 | 54162 |
| 2 | rs2287622 | 169830328 | A | G | 0.40 | 0.020 | 0.003 | 1.0E-14 | 314021 | 0.021 | 0.016 | 0.19 | 54162 |
| 2 | rs7569317 | 203527979 | C | T | 0.53 | 0.019 | 0.003 | 2.8E-14 | 313169 | 0.032 | 0.016 | 0.04 | 54162 |
| 2 | rs1250258 | 216300185 | T | C | 0.74 | 0.013 | 0.003 | 3.5E-06 | 311460 | 0.013 | 0.019 | 0.48 | 54162 |
| 2 | rs11568318 | 234665498 | A | C | 0.07 | 0.024 | 0.005 | 2.8E-06 | 313771 | 0.028 | 0.032 | 0.38 | 54162 |
| 3 | rs13076933 | 12327431 | T | G | 0.74 | 0.019 | 0.003 | 6.1E-11 | 308085 | -0.007 | 0.019 | 0.70 | 54162 |
| 3 | rs9834932 | 32535382 | A | G | 0.91 | 0.034 | 0.004 | 3.0E-14 | 313759 | 0.017 | 0.028 | 0.53 | 54162 |
| 3 | rs71311871 | 58420613 | A | G | 0.92 | 0.031 | 0.005 | 1.1E-11 | 313551 | -0.002 | 0.027 | 0.93 | 54162 |
| 3 | rs55921103 | 69810294 | T | G | 0.65 | 0.015 | 0.003 | 5.2E-08 | 308812 | 0.020 | 0.017 | 0.22 | 54162 |
| 3 | rs3732359 | 119536429 | G | A | 0.22 | 0.019 | 0.003 | 5.4E-10 | 312697 | -0.018 | 0.019 | 0.34 | 54162 |
| 3 | rs9841897 | 122282569 | C | T | 0.16 | 0.020 | 0.003 | 3.9E-09 | 311116 | 0.017 | 0.023 | 0.45 | 54162 |
| 3 | rs113177823 | 132217703 | G | A | 0.95 | 0.036 | 0.006 | 1.3E-10 | 311566 | -0.067 | 0.062 | 0.28 | 54162 |
| 3 | rs3932048 | 136258924 | G | C | 0.32 | 0.010 | 0.003 | 2.0E-04 | 312679 | 0.005 | 0.017 | 0.75 | 54162 |
| 4 | rs13108218 | 3443931 | A | G | 0.38 | 0.019 | 0.003 | 4.2E-13 | 304320 | -0.015 | 0.018 | 0.39 | 54162 |
| 4 | rs1458038 | 81164723 | C | T | 0.71 | 0.019 | 0.003 | 5.0E-12 | 309504 | 0.031 | 0.018 | 0.09 | 54162 |
| 4 | rs1229984 | 100239319 | C | T | 0.98 | 0.054 | 0.008 | 2.4E-10 | 314021 | -0.044 | 0.042 | 0.29 | 54162 |
| 4 | rs13107325 | 103188709 | C | T | 0.93 | 0.018 | 0.005 | 1.9E-04 | 314021 | -0.051 | 0.031 | 0.10 | 54162 |
| 5 | rs9686661 | 55861786 | T | C | 0.20 | 0.014 | 0.003 | 5.8E-06 | 313881 | 0.026 | 0.021 | 0.20 | 54162 |
| 5 | rs2925677 | 71953629 | C | G | 0.79 | 0.016 | 0.003 | 3.2E-07 | 313520 | -0.019 | 0.019 | 0.33 | 54162 |
| 5 | rs12916 | 74656539 | C | T | 0.40 | 0.063 | 0.003 | 1.5E-133 | 314021 | 0.005 | 0.016 | 0.77 | 54162 |
| 5 | rs7734476 | 122848876 | A | G | 0.55 | 0.021 | 0.003 | 2.3E-16 | 313404 | 0.018 | 0.016 | 0.24 | 54162 |
| 5 | rs1016988 | 131744574 | T | C | 0.81 | 0.017 | 0.003 | 1.1E-07 | 314021 | 0.019 | 0.020 | 0.34 | 54162 |
| 5 | rs6874202 | 156391628 | C | T | 0.64 | 0.034 | 0.003 | 3.2E-39 | 313911 | 0.007 | 0.017 | 0.70 | 54162 |
| 6 | rs7746081 | 16126934 | G | A | 0.70 | 0.024 | 0.003 | 7.8E-19 | 312515 | -0.006 | 0.017 | 0.74 | 54162 |
| 6 | rs79220007 | 26098474 | T | C | 0.92 | 0.058 | 0.005 | 3.5E-35 | 313689 | 0.015 | 0.034 | 0.67 | 54162 |
| 6 | rs76079263 | 28019665 | G | C | 0.91 | 0.029 | 0.004 | 4.5E-11 | 308658 | -0.015 | 0.033 | 0.65 | 54162 |
| 6 | rs3179865 | 31324194 | A | G | 0.40 | 0.024 | 0.003 | 2.9E-14 | 217327 | -0.151 | 0.080 | 0.06 | 54162 |

| | | | | | | | | | | | | | |
|---|---|---|---|---|---|---|---|---|---|---|---|---|---|
| 6 | rs76967117 | 34603691 | G | A | 0.88 | 0.029 | 0.004 | 2.9E-13 | 313879 | 0.018 | 0.025 | 0.45 | 54162 |
| 6 | rs913499 | 37038432 | A | G | 0.49 | 0.010 | 0.003 | 4.2E-05 | 312884 | 0.018 | 0.016 | 0.26 | 54162 |
| 6 | rs9496567 | 100602753 | G | A | 0.76 | 0.019 | 0.003 | 3.3E-10 | 311900 | 0.008 | 0.019 | 0.67 | 54162 |
| 6 | rs3822855 | 116316882 | T | G | 0.40 | 0.019 | 0.003 | 3.3E-13 | 313351 | -0.019 | 0.016 | 0.24 | 54162 |
| 6 | rs9491699 | 127471533 | T | C | 0.48 | 0.014 | 0.003 | 2.4E-08 | 313790 | 0.012 | 0.016 | 0.45 | 54162 |
| 6 | rs12197047 | 130389211 | A | G | 0.67 | 0.016 | 0.003 | 4.6E-09 | 305348 | 0.011 | 0.018 | 0.52 | 54162 |
| 6 | rs7776054 | 135418916 | A | G | 0.74 | 0.014 | 0.003 | 2.2E-06 | 313004 | 0.000 | 0.018 | 0.99 | 54162 |
| 6 | rs73025516 | 160520806 | A | G | 0.96 | 0.033 | 0.006 | 1.2E-07 | 313343 | -0.006 | 0.043 | 0.89 | 54162 |
| 6 | rs12208357 | 160543148 | T | C | 0.07 | 0.058 | 0.005 | 2.1E-31 | 312667 | -0.084 | 0.030 | 0.00 | 54162 |
| 6 | rs146534110 | 160578069 | T | G | 0.01 | 0.060 | 0.011 | 5.1E-08 | 314021 | 0.091 | 0.097 | 0.35 | 54162 |
| 6 | rs117733303 | 160922870 | G | A | 0.02 | 0.089 | 0.009 | 1.3E-21 | 314021 | -0.004 | 0.127 | 0.97 | 54162 |
| 6 | rs118039278 | 160985526 | A | G | 0.08 | 0.081 | 0.005 | 1.8E-68 | 311954 | -0.057 | 0.037 | 0.13 | 54162 |
| 7 | rs869412 | 1074134 | T | C | 0.77 | 0.012 | 0.003 | 4.5E-05 | 310947 | 0.014 | 0.020 | 0.46 | 54162 |
| 7 | rs836550 | 6440437 | G | A | 0.41 | 0.015 | 0.003 | 4.8E-09 | 312079 | -0.055 | 0.017 | 8.5E-04 | 54162 |
| 7 | rs28406917 | 21449451 | T | C | 0.43 | 0.011 | 0.003 | 1.1E-05 | 309510 | -0.006 | 0.016 | 0.70 | 54162 |
| 7 | rs56130071 | 21598753 | C | G | 0.22 | 0.031 | 0.003 | 2.4E-24 | 311734 | -0.010 | 0.020 | 0.63 | 54162 |
| 7 | rs4722551 | 25991826 | C | T | 0.16 | 0.026 | 0.003 | 5.7E-14 | 314021 | -0.022 | 0.022 | 0.31 | 54162 |
| 7 | rs67050321 | 36169203 | C | T | 0.30 | 0.015 | 0.003 | 6.0E-08 | 310270 | -0.022 | 0.017 | 0.19 | 54162 |
| 7 | rs2073547 | 44582331 | G | A | 0.18 | 0.037 | 0.003 | 2.4E-30 | 314021 | -0.025 | 0.021 | 0.24 | 54162 |
| 7 | rs4148826 | 87074419 | T | C | 0.82 | 0.014 | 0.003 | 1.5E-05 | 310380 | -0.031 | 0.021 | 0.13 | 54162 |
| 7 | rs112758337 | 97977268 | G | A | 0.81 | 0.014 | 0.003 | 2.4E-05 | 312384 | 0.018 | 0.020 | 0.36 | 54162 |
| 7 | rs111338114 | 100330492 | A | G | 0.96 | 0.037 | 0.006 | 6.6E-09 | 293140 | -0.046 | 0.035 | 0.20 | 54162 |
| 7 | rs10231941 | 100532540 | C | T | 0.18 | 0.021 | 0.003 | 7.4E-11 | 312838 | 0.005 | 0.022 | 0.84 | 54162 |
| 8 | rs1350559 | 9367743 | G | C | 0.40 | 0.015 | 0.003 | 5.2E-09 | 308947 | 0.007 | 0.016 | 0.65 | 54162 |
| 8 | rs1495741 | 18272881 | G | A | 0.22 | 0.020 | 0.003 | 7.6E-11 | 314021 | -0.004 | 0.019 | 0.83 | 54162 |
| 8 | rs59328596 | 21928227 | G | A | 0.85 | 0.019 | 0.004 | 1.4E-07 | 313645 | -0.041 | 0.023 | 0.08 | 54162 |
| 8 | rs9298506 | 55437524 | G | A | 0.21 | 0.022 | 0.003 | 5.5E-13 | 314021 | 0.016 | 0.020 | 0.41 | 54162 |
| 8 | rs4620259 | 109991668 | C | A | 0.19 | 0.015 | 0.003 | 4.7E-06 | 306584 | 0.001 | 0.020 | 0.98 | 54162 |
| 8 | rs2737265 | 116667634 | A | G | 0.72 | 0.021 | 0.003 | 1.8E-13 | 312899 | -0.035 | 0.017 | 0.04 | 54162 |
| 8 | rs28601761 | 126500031 | C | G | 0.58 | 0.060 | 0.003 | 5.3E-117 | 299703 | 0.004 | 0.017 | 0.82 | 54162 |
| 8 | rs11786083 | 145050358 | A | G | 0.37 | 0.015 | 0.003 | 7.3E-09 | 306927 | 0.023 | 0.018 | 0.19 | 54162 |
| 9 | rs3780181 | 2640759 | A | G | 0.93 | 0.029 | 0.005 | 9.4E-09 | 311543 | 0.071 | 0.031 | 0.02 | 54162 |
| 9 | rs6475606 | 22081850 | C | T | 0.52 | 0.021 | 0.003 | 5.4E-17 | 314021 | -0.032 | 0.016 | 0.04 | 54162 |
| 9 | rs6560499 | 78730766 | G | A | 0.42 | 0.014 | 0.003 | 1.3E-07 | 306861 | -0.009 | 0.016 | 0.57 | 54162 |
| 9 | rs2066714 | 107586753 | C | T | 0.13 | 0.025 | 0.004 | 3.6E-11 | 314021 | -0.082 | 0.033 | 0.01 | 54162 |
| 9 | rs11789603 | 107647019 | T | C | 0.11 | 0.024 | 0.004 | 3.6E-09 | 313207 | 0.021 | 0.026 | 0.43 | 54162 |
| 9 | rs2740488 | 107661742 | A | C | 0.74 | 0.022 | 0.003 | 4.5E-15 | 312497 | -0.052 | 0.018 | 0.00 | 54162 |
| 9 | rs13283282 | 131465481 | C | G | 0.85 | 0.023 | 0.004 | 1.9E-10 | 314021 | 0.011 | 0.030 | 0.73 | 54162 |
| 9 | rs10448340 | 139320069 | T | G | 0.68 | 0.015 | 0.003 | 2.0E-08 | 311982 | 0.021 | 0.018 | 0.23 | 54162 |
| 10 | rs11014204 | 18720845 | T | C | 0.28 | 0.015 | 0.003 | 1.3E-07 | 311499 | 0.024 | 0.018 | 0.17 | 54162 |
| 10 | rs79828839 | 52352431 | T | C | 0.20 | 0.015 | 0.003 | 3.2E-06 | 313934 | -0.027 | 0.020 | 0.18 | 54162 |
| 10 | rs7090758 | 65335315 | T | C | 0.53 | 0.011 | 0.003 | 9.4E-06 | 313865 | -0.004 | 0.016 | 0.82 | 54162 |
| 10 | rs17476364 | 71094504 | T | C | 0.89 | 0.023 | 0.004 | 1.2E-08 | 313322 | 0.010 | 0.028 | 0.73 | 54162 |
| 10 | rs2068888 | 94839642 | G | A | 0.55 | 0.020 | 0.003 | 1.3E-15 | 314021 | 0.021 | 0.016 | 0.20 | 54162 |
| 10 | rs2250802 | 113921354 | G | A | 0.27 | 0.019 | 0.003 | 4.3E-11 | 313791 | 0.012 | 0.017 | 0.49 | 54162 |
| 10 | rs12246352 | 124705307 | G | A | 0.10 | 0.028 | 0.004 | 1.6E-11 | 312518 | -0.020 | 0.027 | 0.46 | 54162 |
| 11 | rs7108486 | 5677158 | T | C | 0.98 | 0.041 | 0.008 | 1.3E-06 | 312791 | -0.059 | 0.063 | 0.35 | 54162 |
| 11 | rs11601507 | 5701074 | A | C | 0.07 | 0.032 | 0.005 | 1.0E-10 | 314021 | -0.025 | 0.044 | 0.58 | 54162 |
| 11 | rs10832963 | 18664241 | G | T | 0.75 | 0.016 | 0.003 | 1.3E-08 | 312156 | 0.006 | 0.018 | 0.72 | 54162 |
| 11 | rs174564 | 61588305 | A | G | 0.65 | 0.033 | 0.003 | 5.5E-36 | 313455 | 0.010 | 0.016 | 0.54 | 54162 |
| 11 | rs11227247 | 65422853 | C | A | 0.13 | 0.018 | 0.004 | 1.1E-06 | 313988 | -0.015 | 0.023 | 0.51 | 54162 |
| 11 | rs74869459 | 66296569 | T | C | 0.76 | 0.017 | 0.003 | 4.6E-09 | 314004 | 0.010 | 0.018 | 0.60 | 54162 |
| 11 | rs964184 | 116648917 | G | C | 0.13 | 0.058 | 0.004 | 1.2E-55 | 314021 | 0.021 | 0.023 | 0.36 | 54162 |
| 11 | rs6589939 | 122518525 | G | A | 0.38 | 0.012 | 0.003 | 1.5E-06 | 312369 | 0.030 | 0.016 | 0.06 | 54162 |
| 11 | rs59379014 | 126228000 | T | C | 0.07 | 0.055 | 0.005 | 3.6E-30 | 313827 | 0.007 | 0.032 | 0.84 | 54162 |
| 12 | rs35882350 | 623129 | G | A | 0.26 | 0.017 | 0.003 | 1.9E-09 | 314021 | -0.006 | 0.020 | 0.78 | 54162 |
| 12 | rs1007938 | 26802549 | G | A | 0.41 | 0.013 | 0.003 | 3.2E-07 | 307069 | -0.007 | 0.016 | 0.66 | 54162 |
| 12 | rs2160994 | 50650057 | C | T | 0.65 | 0.019 | 0.003 | 9.4E-13 | 311870 | 0.000 | 0.018 | 0.99 | 54162 |
| 12 | rs597808 | 111973358 | G | A | 0.52 | 0.027 | 0.003 | 1.8E-26 | 311431 | 0.034 | 0.016 | 0.03 | 54162 |
| 12 | rs233721 | 113031543 | A | T | 0.65 | 0.021 | 0.003 | 3.3E-15 | 307366 | 0.028 | 0.017 | 0.09 | 54162 |
| 12 | rs1169294 | 121426594 | A | G | 0.31 | 0.025 | 0.003 | 6.4E-20 | 311706 | -0.015 | 0.017 | 0.38 | 54162 |

| | | | | | | | | | | | | | |
|---|---|---|---|---|---|---|---|---|---|---|---|---|---|
| 12 | rs112403212 | 125303254 | T | C | 0.14 | 0.017 | 0.004 | 7.2E-06 | 309363 | -0.040 | 0.025 | 0.11 | 54162 |
| 13 | rs2238162 | 32959199 | C | T | 0.48 | 0.017 | 0.003 | 1.1E-11 | 313761 | 0.005 | 0.015 | 0.73 | 54162 |
| 13 | rs6602912 | 114546549 | G | T | 0.28 | 0.024 | 0.003 | 3.7E-17 | 312202 | -0.006 | 0.018 | 0.72 | 54162 |
| 14 | rs11621792 | 24871926 | T | C | 0.45 | 0.019 | 0.003 | 4.4E-13 | 305593 | -0.025 | 0.018 | 0.16 | 54162 |
| 14 | rs8008068 | 64233717 | G | A | 0.16 | 0.015 | 0.003 | 1.2E-05 | 313567 | 0.010 | 0.022 | 0.64 | 54162 |
| 14 | rs6573971 | 71011469 | G | A | 0.45 | 0.012 | 0.003 | 4.6E-06 | 300526 | -0.018 | 0.016 | 0.25 | 54162 |
| 14 | rs61988556 | 73439258 | T | C | 0.91 | 0.022 | 0.004 | 1.2E-06 | 314021 | -0.052 | 0.029 | 0.07 | 54162 |
| 14 | rs145730801 | 94768196 | C | T | 0.04 | 0.037 | 0.006 | 4.7E-09 | 309822 | 0.077 | 0.045 | 0.09 | 54162 |
| 15 | rs10851478 | 49829019 | T | C | 0.58 | 0.012 | 0.003 | 3.4E-06 | 313568 | -0.007 | 0.016 | 0.65 | 54162 |
| 15 | rs72733928 | 57512284 | T | A | 0.06 | 0.026 | 0.005 | 9.4E-07 | 312992 | -0.005 | 0.031 | 0.87 | 54162 |
| 15 | rs1532085 | 58683366 | A | G | 0.39 | 0.018 | 0.003 | 5.1E-12 | 314021 | -0.039 | 0.016 | 0.01 | 54162 |
| 15 | rs261334 | 58726744 | G | C | 0.21 | 0.023 | 0.003 | 1.6E-13 | 314021 | -0.011 | 0.019 | 0.55 | 54162 |
| 15 | rs62011285 | 63791063 | C | T | 0.34 | 0.013 | 0.003 | 6.8E-07 | 312380 | 0.028 | 0.016 | 0.09 | 54162 |
| 16 | rs12445804 | 11706100 | A | G | 0.07 | 0.025 | 0.005 | 4.6E-07 | 309828 | 0.059 | 0.037 | 0.11 | 54162 |
| 16 | rs62033400 | 53811788 | A | G | 0.61 | 0.012 | 0.003 | 5.0E-06 | 313537 | -0.007 | 0.016 | 0.65 | 54162 |
| 16 | rs3764261 | 56993324 | C | A | 0.67 | 0.032 | 0.003 | 1.2E-33 | 314021 | -0.003 | 0.017 | 0.85 | 54162 |
| 16 | rs34042070 | 72101525 | G | C | 0.19 | 0.051 | 0.003 | 7.4E-56 | 310247 | -0.009 | 0.020 | 0.64 | 54162 |
| 16 | rs7202323 | 72217113 | T | G | 0.77 | 0.025 | 0.003 | 1.5E-16 | 312880 | 0.014 | 0.018 | 0.45 | 54162 |
| 17 | rs55714927 | 7080316 | C | T | 0.81 | 0.028 | 0.003 | 8.3E-19 | 314021 | -0.006 | 0.028 | 0.84 | 54162 |
| 17 | rs9894946 | 7571080 | A | G | 0.15 | 0.018 | 0.004 | 6.2E-07 | 302717 | 0.021 | 0.024 | 0.37 | 54162 |
| 17 | rs704 | 26694861 | A | G | 0.47 | 0.016 | 0.003 | 4.7E-10 | 314021 | -0.006 | 0.017 | 0.73 | 54162 |
| 17 | rs56208742 | 27884667 | T | C | 0.97 | 0.039 | 0.007 | 7.9E-08 | 314021 | -0.087 | 0.074 | 0.24 | 54162 |
| 17 | rs12603885 | 29466722 | A | G | 0.70 | 0.017 | 0.003 | 3.2E-10 | 313768 | -0.029 | 0.018 | 0.10 | 54162 |
| 17 | rs36043200 | 45629406 | G | A | 0.48 | 0.026 | 0.003 | 7.2E-24 | 309916 | -0.053 | 0.020 | 0.01 | 54162 |
| 17 | rs3110609 | 46753543 | T | C | 0.66 | 0.017 | 0.003 | 1.7E-10 | 310282 | 0.002 | 0.017 | 0.90 | 54162 |
| 17 | rs1801689 | 64210580 | C | A | 0.03 | 0.065 | 0.007 | 1.6E-18 | 314021 | -0.144 | 0.076 | 0.06 | 54162 |
| 17 | rs12936113 | 66401063 | C | T | 0.76 | 0.013 | 0.003 | 2.1E-05 | 310505 | 0.009 | 0.021 | 0.65 | 54162 |
| 17 | rs77542162 | 67081278 | G | A | 0.02 | 0.126 | 0.008 | 3.1E-51 | 314021 | 0.006 | 0.079 | 0.94 | 54162 |
| 17 | rs72631343 | 67191270 | C | G | 0.87 | 0.032 | 0.004 | 6.0E-17 | 314021 | -0.042 | 0.026 | 0.11 | 54162 |
| 17 | rs12948394 | 76382791 | C | T | 0.52 | 0.018 | 0.003 | 1.4E-11 | 295044 | 0.030 | 0.016 | 0.06 | 54162 |
| 18 | rs77960347 | 47109955 | G | A | 0.01 | 0.079 | 0.011 | 3.5E-13 | 314021 | 0.024 | 0.058 | 0.67 | 54162 |
| 18 | rs7241918 | 47160953 | T | G | 0.82 | 0.017 | 0.003 | 1.2E-07 | 314021 | -0.015 | 0.021 | 0.48 | 54162 |
| 19 | rs143020224 | 11187324 | C | G | 0.88 | 0.176 | 0.004 | 0.0E+00 | 313884 | 0.019 | 0.028 | 0.49 | 54162 |
| 19 | rs2738447 | 11227480 | C | A | 0.59 | 0.043 | 0.003 | 2.2E-62 | 313312 | 0.001 | 0.016 | 0.94 | 54162 |
| 19 | rs8101801 | 11335477 | C | A | 0.97 | 0.037 | 0.007 | 9.0E-08 | 313859 | -0.039 | 0.043 | 0.36 | 54162 |
| 19 | rs62120394 | 18338709 | A | G | 0.29 | 0.020 | 0.003 | 3.7E-13 | 311339 | 0.006 | 0.017 | 0.74 | 54162 |
| 19 | rs8107974 | 19388500 | A | T | 0.92 | 0.103 | 0.005 | 1.5E-104 | 313654 | -0.005 | 0.047 | 0.92 | 54162 |
| 19 | rs56113850 | 41353107 | C | T | 0.58 | 0.014 | 0.003 | 1.4E-08 | 311479 | 0.027 | 0.023 | 0.24 | 54162 |
| 19 | rs2021092 | 44068706 | T | C | 0.81 | 0.016 | 0.003 | 8.7E-07 | 312844 | 0.014 | 0.020 | 0.49 | 54162 |
| 19 | rs62116889 | 45022560 | T | C | 0.93 | 0.055 | 0.005 | 2.3E-27 | 313015 | 0.113 | 0.044 | 0.01 | 54162 |
| 19 | rs204469 | 45490285 | G | A | 0.96 | 0.030 | 0.006 | 1.4E-06 | 312239 | 0.111 | 0.046 | 0.02 | 54162 |
| 19 | rs35081008 | 58662235 | C | T | 0.85 | 0.035 | 0.004 | 2.0E-22 | 311538 | -0.002 | 0.028 | 0.95 | 54162 |
| 20 | rs73075609 | 5580789 | T | C | 0.03 | 0.037 | 0.008 | 1.9E-06 | 313162 | 0.097 | 0.061 | 0.11 | 54162 |
| 20 | rs438568 | 12958687 | G | A | 0.61 | 0.011 | 0.003 | 1.8E-05 | 312712 | -0.015 | 0.016 | 0.33 | 54162 |
| 20 | rs61433703 | 17804068 | A | G | 0.16 | 0.019 | 0.003 | 8.7E-08 | 307149 | 0.012 | 0.023 | 0.61 | 54162 |
| 20 | rs2618566 | 17844684 | G | T | 0.34 | 0.024 | 0.003 | 3.1E-19 | 314021 | 0.022 | 0.017 | 0.20 | 54162 |
| 20 | rs6050463 | 25208990 | A | G | 0.49 | 0.013 | 0.003 | 5.6E-07 | 313853 | 0.000 | 0.015 | 0.98 | 54162 |
| 20 | rs224424 | 34147998 | A | G | 0.79 | 0.020 | 0.003 | 8.6E-11 | 313839 | 0.004 | 0.018 | 0.82 | 54162 |
| 20 | rs6093446 | 39780932 | A | G | 0.29 | 0.020 | 0.003 | 5.7E-13 | 313671 | 0.006 | 0.017 | 0.73 | 54162 |
| 20 | rs1800961 | 43042364 | C | T | 0.97 | 0.058 | 0.007 | 7.8E-16 | 314021 | -0.060 | 0.047 | 0.20 | 54162 |
| 20 | rs6073958 | 44551855 | C | T | 0.20 | 0.017 | 0.003 | 1.5E-07 | 313591 | 0.027 | 0.021 | 0.19 | 54162 |
| 20 | rs2256814 | 62373983 | A | G | 0.20 | 0.015 | 0.003 | 1.1E-06 | 311634 | 0.044 | 0.022 | 0.04 | 54162 |
| 20 | rs6090101 | 62909520 | A | G | 0.20 | 0.018 | 0.003 | 8.4E-09 | 307859 | -0.042 | 0.085 | 0.62 | 54162 |
| 21 | rs4818025 | 40709171 | G | A | 0.57 | 0.013 | 0.003 | 2.0E-07 | 311892 | -0.005 | 0.016 | 0.73 | 54162 |
| 22 | rs960596 | 41393520 | T | C | 0.34 | 0.013 | 0.003 | 8.9E-07 | 307460 | 0.002 | 0.017 | 0.91 | 54162 |
| 22 | rs12162782 | 50853626 | G | T | 0.34 | 0.013 | 0.003 | 9.3E-07 | 314021 | -0.010 | 0.017 | 0.57 | 54162 |

**Supplementary table 3:** List of 423 SNPs used as instrumental variables for HDL-C, and their association with HDL-C and dementia.

| CHR | rsID | POS | Effect allele | Other allele | EAF | BETA HDL-C | SE HDL-C | Pval HDL-C | n HDL-C | BETA dementia | SE dementia | Pval dementia | n dementia |
|---|---|---|---|---|---|---|---|---|---|---|---|---|---|
| 1 | rs2298214 | 935222 | C | A | 0.42 | 0.013 | 0.002 | 4.4E-08 | 285966 | -0.042 | 0.030 | 0.15 | 54162 |
| 1 | rs2298632 | 23710475 | T | C | 0.50 | 0.015 | 0.002 | 3.2E-10 | 273566 | 0.002 | 0.016 | 0.93 | 54162 |
| 1 | rs72654647 | 25022314 | G | A | 0.76 | 0.014 | 0.003 | 2.9E-07 | 292386 | -0.002 | 0.018 | 0.89 | 54162 |
| 1 | rs17185038 | 28219658 | G | C | 0.07 | 0.022 | 0.005 | 2.2E-06 | 294473 | -0.008 | 0.049 | 0.87 | 54162 |
| 1 | rs4654395 | 29567412 | C | T | 0.47 | 0.010 | 0.002 | 4.1E-05 | 289617 | -0.013 | 0.016 | 0.42 | 54162 |
| 1 | rs3768321 | 40035928 | G | T | 0.80 | 0.044 | 0.003 | 2.8E-49 | 293556 | -0.023 | 0.021 | 0.26 | 54162 |
| 1 | rs74328314 | 61670759 | G | A | 0.07 | 0.026 | 0.005 | 1.4E-08 | 287852 | -0.011 | 0.033 | 0.73 | 54162 |
| 1 | rs1168114 | 63156043 | G | A | 0.65 | 0.014 | 0.002 | 9.1E-09 | 293301 | -0.041 | 0.017 | 0.01 | 54162 |
| 1 | rs6664374 | 66073952 | T | C | 0.35 | 0.016 | 0.002 | 3.4E-10 | 290069 | 0.020 | 0.017 | 0.23 | 54162 |
| 1 | rs771481 | 93846653 | A | T | 0.18 | 0.030 | 0.003 | 4.4E-23 | 294174 | -0.029 | 0.021 | 0.16 | 54162 |
| 1 | rs12740374 | 109817590 | T | G | 0.22 | 0.030 | 0.003 | 5.8E-26 | 294473 | 0.000 | 0.019 | 0.98 | 54162 |
| 1 | rs12045101 | 110267651 | C | T | 0.76 | 0.016 | 0.003 | 1.1E-08 | 292224 | 0.006 | 0.019 | 0.73 | 54162 |
| 1 | rs267738 | 150940625 | G | T | 0.22 | 0.022 | 0.003 | 3.0E-14 | 294473 | -0.033 | 0.020 | 0.09 | 54162 |
| 1 | rs113261881 | 154997572 | G | A | 0.94 | 0.025 | 0.005 | 5.2E-07 | 291221 | -0.044 | 0.039 | 0.26 | 54162 |
| 1 | rs4233367 | 161163037 | C | T | 0.61 | 0.011 | 0.002 | 2.4E-06 | 294473 | -0.016 | 0.016 | 0.33 | 54162 |
| 1 | rs34720381 | 171455322 | C | T | 0.91 | 0.020 | 0.004 | 6.9E-07 | 293344 | -0.002 | 0.028 | 0.94 | 54162 |
| 1 | rs4650994 | 178515312 | G | A | 0.47 | 0.021 | 0.002 | 2.7E-18 | 294473 | -0.004 | 0.016 | 0.80 | 54162 |
| 1 | rs61805075 | 182136401 | G | A | 0.67 | 0.024 | 0.003 | 4.7E-21 | 292959 | -0.006 | 0.016 | 0.70 | 54162 |
| 1 | rs12119128 | 201771326 | G | A | 0.70 | 0.010 | 0.003 | 9.2E-05 | 277783 | -0.006 | 0.018 | 0.72 | 54162 |
| 1 | rs3903399 | 205041542 | T | C | 0.79 | 0.014 | 0.003 | 1.6E-06 | 293561 | -0.007 | 0.020 | 0.73 | 54162 |
| 1 | rs3747973 | 205677148 | G | A | 0.59 | 0.013 | 0.002 | 6.5E-08 | 291298 | -0.011 | 0.016 | 0.49 | 54162 |
| 1 | rs2642438 | 220970028 | G | A | 0.70 | 0.029 | 0.003 | 9.0E-29 | 294473 | 0.006 | 0.019 | 0.77 | 54162 |
| 1 | rs10916239 | 228088833 | C | T | 0.38 | 0.013 | 0.002 | 6.9E-08 | 290531 | 0.012 | 0.017 | 0.47 | 54162 |
| 1 | rs2281718 | 230297778 | T | A | 0.61 | 0.062 | 0.002 | 1.4E-142 | 293390 | 0.011 | 0.017 | 0.54 | 54162 |
| 1 | rs1043897 | 230416399 | T | G | 0.41 | 0.021 | 0.002 | 4.8E-18 | 290815 | -0.019 | 0.016 | 0.24 | 54162 |
| 1 | rs557933 | 234853268 | C | A | 0.52 | 0.018 | 0.002 | 2.1E-14 | 291456 | 0.020 | 0.016 | 0.20 | 54162 |
| 2 | rs7595075 | 264019 | A | C | 0.35 | 0.014 | 0.002 | 1.3E-08 | 293043 | -0.056 | 0.016 | 5.6E-04 | 54162 |
| 2 | rs907866 | 20371380 | G | A | 0.55 | 0.019 | 0.002 | 5.2E-15 | 291269 | 0.024 | 0.016 | 0.13 | 54162 |
| 2 | rs676210 | 21231524 | A | G | 0.20 | 0.064 | 0.003 | 6.5E-105 | 294473 | -0.052 | 0.019 | 0.01 | 54162 |
| 2 | rs2362541 | 30478453 | T | G | 0.49 | 0.009 | 0.002 | 1.2E-04 | 293051 | -0.018 | 0.016 | 0.27 | 54162 |
| 2 | rs12998038 | 42602387 | T | C | 0.26 | 0.010 | 0.003 | 3.6E-04 | 288820 | 0.053 | 0.017 | 2.2E-03 | 54162 |
| 2 | rs12713007 | 48484467 | C | T | 0.50 | 0.010 | 0.002 | 3.8E-05 | 287932 | 0.019 | 0.015 | 0.23 | 54162 |
| 2 | rs17326656 | 48962291 | G | T | 0.76 | 0.019 | 0.003 | 2.3E-11 | 291305 | -0.004 | 0.019 | 0.85 | 54162 |
| 2 | rs12986742 | 58975143 | T | C | 0.52 | 0.010 | 0.002 | 1.9E-05 | 288187 | -0.029 | 0.016 | 0.06 | 54162 |
| 2 | rs2723065 | 65279414 | G | A | 0.37 | 0.014 | 0.002 | 6.7E-09 | 294473 | -0.017 | 0.016 | 0.27 | 54162 |
| 2 | rs11883967 | 66673862 | C | A | 0.66 | 0.013 | 0.003 | 2.3E-07 | 289497 | 0.019 | 0.016 | 0.24 | 54162 |
| 2 | rs4599108 | 85543222 | T | C | 0.48 | 0.014 | 0.002 | 2.4E-08 | 278731 | -0.004 | 0.016 | 0.79 | 54162 |
| 2 | rs7583067 | 100796850 | T | C | 0.24 | 0.016 | 0.003 | 1.5E-08 | 290015 | -0.031 | 0.019 | 0.09 | 54162 |
| 2 | rs9646934 | 111822002 | G | C | 0.68 | 0.013 | 0.003 | 7.7E-07 | 284578 | 0.011 | 0.017 | 0.52 | 54162 |
| 2 | rs17041868 | 111894720 | T | C | 0.94 | 0.026 | 0.005 | 4.8E-08 | 294473 | -0.010 | 0.034 | 0.77 | 54162 |
| 2 | rs4550673 | 112941372 | A | G | 0.92 | 0.017 | 0.004 | 9.2E-05 | 294473 | -0.013 | 0.027 | 0.64 | 54162 |
| 2 | rs11688682 | 121347612 | C | G | 0.26 | 0.013 | 0.003 | 4.4E-06 | 265017 | 0.005 | 0.022 | 0.83 | 54162 |
| 2 | rs35706812 | 128593977 | A | G | 0.57 | 0.013 | 0.002 | 2.0E-08 | 293986 | 0.009 | 0.016 | 0.57 | 54162 |
| 2 | rs56131490 | 135263081 | A | G | 0.15 | 0.016 | 0.003 | 1.1E-06 | 294473 | -0.025 | 0.021 | 0.24 | 54162 |
| 2 | rs1446585 | 136407479 | G | A | 0.23 | 0.020 | 0.003 | 9.5E-13 | 294473 | 0.028 | 0.018 | 0.11 | 54162 |
| 2 | rs12692596 | 161265910 | C | T | 0.63 | 0.012 | 0.002 | 2.2E-06 | 291885 | -0.032 | 0.017 | 0.06 | 54162 |
| 2 | rs13389219 | 165528876 | T | C | 0.39 | 0.026 | 0.002 | 5.4E-27 | 294357 | -0.027 | 0.016 | 0.09 | 54162 |
| 2 | rs2364723 | 178126546 | C | G | 0.32 | 0.013 | 0.003 | 2.9E-07 | 291830 | 0.005 | 0.017 | 0.77 | 54162 |
| 2 | rs72926946 | 203477868 | C | A | 0.70 | 0.018 | 0.003 | 7.5E-13 | 293750 | 0.029 | 0.017 | 0.09 | 54162 |
| 2 | rs2551980 | 208518317 | A | G | 0.18 | 0.014 | 0.003 | 7.9E-06 | 292668 | -0.006 | 0.020 | 0.76 | 54162 |
| 2 | rs1047891 | 211540507 | C | A | 0.68 | 0.018 | 0.003 | 3.1E-12 | 294473 | 0.015 | 0.020 | 0.43 | 54162 |
| 2 | rs1517500 | 226306993 | T | C | 0.83 | 0.016 | 0.003 | 6.5E-07 | 293158 | -0.016 | 0.022 | 0.46 | 54162 |
| 2 | rs2943645 | 227099180 | C | T | 0.35 | 0.044 | 0.002 | 4.2E-70 | 294473 | -0.011 | 0.016 | 0.52 | 54162 |
| 2 | rs57074291 | 227229344 | G | C | 0.26 | 0.013 | 0.003 | 8.8E-07 | 293149 | 0.004 | 0.018 | 0.81 | 54162 |
| 2 | rs6738438 | 230020220 | C | T | 0.64 | 0.013 | 0.002 | 2.0E-07 | 290537 | 0.007 | 0.017 | 0.69 | 54162 |
| 2 | rs59104589 | 242237902 | T | C | 0.36 | 0.016 | 0.002 | 7.0E-11 | 294274 | 0.001 | 0.017 | 0.98 | 54162 |
| 3 | rs13076933 | 12327431 | T | G | 0.74 | 0.013 | 0.003 | 2.0E-06 | 288879 | -0.007 | 0.019 | 0.70 | 54162 |
| 3 | rs12485478 | 12351223 | A | G | 0.97 | 0.056 | 0.007 | 3.9E-14 | 294153 | -0.024 | 0.046 | 0.61 | 54162 |

| | | | | | | | | | | | | | |
|---|---|---|---|---|---|---|---|---|---|---|---|---|---|
| 3 | rs13323506 | 12737231 | C | A | 0.69 | 0.014 | 0.003 | 1.1E-07 | 293486 | -0.006 | 0.017 | 0.72 | 54162 |
| 3 | rs13097947 | 15846011 | C | T | 0.65 | 0.017 | 0.003 | 4.8E-11 | 272001 | -0.004 | 0.017 | 0.79 | 54162 |
| 3 | rs7622114 | 36960660 | A | C | 0.58 | 0.012 | 0.002 | 5.3E-07 | 286922 | -0.009 | 0.016 | 0.56 | 54162 |
| 3 | rs2100692 | 48193753 | A | G | 0.09 | 0.021 | 0.004 | 1.4E-07 | 290855 | 0.074 | 0.054 | 0.17 | 54162 |
| 3 | rs74735576 | 48357600 | G | A | 0.98 | 0.063 | 0.009 | 2.3E-11 | 294473 | 0.016 | 0.064 | 0.80 | 54162 |
| 3 | rs6765484 | 50041313 | T | C | 0.47 | 0.024 | 0.002 | 5.1E-24 | 292998 | -0.009 | 0.016 | 0.56 | 54162 |
| 3 | rs73082723 | 51926817 | A | C | 0.21 | 0.020 | 0.003 | 1.2E-10 | 267592 | -0.008 | 0.035 | 0.82 | 54162 |
| 3 | rs2159607 | 52501451 | G | T | 0.19 | 0.024 | 0.003 | 4.4E-16 | 294186 | 0.026 | 0.021 | 0.20 | 54162 |
| 3 | rs830620 | 71679148 | T | C | 0.42 | 0.013 | 0.002 | 1.9E-08 | 294473 | 0.006 | 0.016 | 0.73 | 54162 |
| 3 | rs4855582 | 108867705 | T | C | 0.43 | 0.012 | 0.002 | 5.8E-07 | 294424 | -0.004 | 0.016 | 0.78 | 54162 |
| 3 | rs7650845 | 114484372 | C | T | 0.18 | 0.014 | 0.003 | 3.1E-06 | 290041 | 0.015 | 0.020 | 0.46 | 54162 |
| 3 | rs3732356 | 119529113 | G | T | 0.06 | 0.031 | 0.005 | 2.2E-10 | 291995 | 0.031 | 0.039 | 0.43 | 54162 |
| 3 | rs6806529 | 123049938 | C | A | 0.56 | 0.010 | 0.002 | 1.8E-05 | 287749 | 0.008 | 0.016 | 0.64 | 54162 |
| 3 | rs113761591 | 127305355 | C | T | 0.80 | 0.018 | 0.003 | 1.3E-09 | 293632 | -0.018 | 0.020 | 0.37 | 54162 |
| 3 | rs1225053 | 131642852 | T | C | 0.74 | 0.014 | 0.003 | 1.2E-07 | 291332 | 0.018 | 0.018 | 0.32 | 54162 |
| 3 | rs6762415 | 133478557 | T | G | 0.46 | 0.016 | 0.002 | 2.3E-11 | 292266 | -0.004 | 0.016 | 0.82 | 54162 |
| 3 | rs9647335 | 135880410 | T | A | 0.19 | 0.027 | 0.003 | 2.6E-19 | 291760 | 0.036 | 0.020 | 0.07 | 54162 |
| 3 | rs62271373 | 150066540 | T | A | 0.94 | 0.037 | 0.005 | 8.8E-13 | 289129 | 0.037 | 0.035 | 0.30 | 54162 |
| 3 | rs1086056 | 154088411 | T | G | 0.16 | 0.017 | 0.003 | 1.6E-07 | 293445 | -0.025 | 0.023 | 0.26 | 54162 |
| 3 | rs9817452 | 156795414 | T | G | 0.39 | 0.020 | 0.002 | 1.1E-16 | 292562 | 0.016 | 0.017 | 0.33 | 54162 |
| 3 | rs6790951 | 160025287 | C | T | 0.52 | 0.012 | 0.002 | 2.6E-07 | 294224 | -0.024 | 0.015 | 0.11 | 54162 |
| 3 | rs11546878 | 183976103 | T | C | 0.18 | 0.012 | 0.003 | 1.0E-04 | 294473 | -0.038 | 0.024 | 0.11 | 54162 |
| 3 | rs10513801 | 185822353 | T | G | 0.86 | 0.029 | 0.003 | 1.8E-17 | 293330 | 0.035 | 0.024 | 0.14 | 54162 |
| 3 | rs4686739 | 185878419 | G | A | 0.64 | 0.015 | 0.002 | 5.2E-10 | 290923 | 0.003 | 0.016 | 0.87 | 54162 |
| 3 | rs2268840 | 185931174 | C | T | 0.23 | 0.016 | 0.003 | 2.1E-08 | 294473 | -0.011 | 0.021 | 0.60 | 54162 |
| 3 | rs139828053 | 195298892 | T | C | 0.97 | 0.028 | 0.007 | 4.4E-05 | 292379 | 0.062 | 0.092 | 0.50 | 54162 |
| 3 | rs9877304 | 196073072 | G | A | 0.74 | 0.012 | 0.003 | 9.6E-06 | 289530 | 0.003 | 0.020 | 0.90 | 54162 |
| 4 | rs880674 | 2250109 | C | T | 0.14 | 0.016 | 0.003 | 3.4E-06 | 293614 | -0.047 | 0.022 | 0.04 | 54162 |
| 4 | rs11938781 | 17924734 | T | C | 0.84 | 0.015 | 0.003 | 2.9E-06 | 291581 | -0.030 | 0.021 | 0.15 | 54162 |
| 4 | rs1395221 | 24626903 | G | T | 0.60 | 0.011 | 0.002 | 1.1E-05 | 289009 | 0.004 | 0.016 | 0.79 | 54162 |
| 4 | rs73243877 | 26047616 | A | G | 0.83 | 0.026 | 0.003 | 1.9E-16 | 294473 | 0.001 | 0.021 | 0.97 | 54162 |
| 4 | rs1055582 | 39700173 | C | T | 0.51 | 0.013 | 0.002 | 9.7E-08 | 292700 | 0.014 | 0.016 | 0.40 | 54162 |
| 4 | rs2237035 | 55526251 | T | G | 0.39 | 0.015 | 0.002 | 2.3E-10 | 293589 | 0.022 | 0.016 | 0.17 | 54162 |
| 4 | rs1349852 | 69533217 | C | A | 0.47 | 0.012 | 0.002 | 1.0E-06 | 292198 | -0.030 | 0.029 | 0.30 | 54162 |
| 4 | rs2175766 | 76572191 | C | A | 0.54 | 0.013 | 0.002 | 1.2E-07 | 294173 | -0.007 | 0.015 | 0.65 | 54162 |
| 4 | rs13111599 | 83917037 | G | A | 0.74 | 0.013 | 0.003 | 1.4E-06 | 292898 | -0.021 | 0.018 | 0.23 | 54162 |
| 4 | rs3775228 | 87985166 | C | T | 0.60 | 0.020 | 0.002 | 2.8E-16 | 290280 | -0.002 | 0.016 | 0.89 | 54162 |
| 4 | rs6824451 | 89723065 | G | A | 0.54 | 0.021 | 0.002 | 4.3E-19 | 293658 | -0.010 | 0.015 | 0.52 | 54162 |
| 4 | rs12650112 | 99788480 | T | C | 0.35 | 0.016 | 0.003 | 1.8E-10 | 284889 | -0.020 | 0.016 | 0.21 | 54162 |
| 4 | rs13107325 | 103188709 | C | T | 0.93 | 0.083 | 0.004 | 5.7E-77 | 294473 | -0.051 | 0.031 | 0.10 | 54162 |
| 4 | rs62338910 | 104253889 | G | A | 0.23 | 0.016 | 0.003 | 6.4E-09 | 293975 | -0.020 | 0.018 | 0.28 | 54162 |
| 4 | rs9884482 | 106081636 | T | C | 0.63 | 0.012 | 0.002 | 3.3E-07 | 294473 | -0.016 | 0.016 | 0.31 | 54162 |
| 4 | rs78025076 | 110569620 | C | T | 0.98 | 0.051 | 0.008 | 7.7E-10 | 294473 | -0.253 | 0.135 | 0.06 | 54162 |
| 4 | rs28455602 | 120034116 | G | A | 0.18 | 0.016 | 0.003 | 5.1E-07 | 291934 | 0.007 | 0.020 | 0.72 | 54162 |
| 4 | rs13144151 | 146403165 | G | A | 0.86 | 0.017 | 0.003 | 5.0E-07 | 287588 | 0.019 | 0.025 | 0.45 | 54162 |
| 4 | rs4691379 | 157706904 | T | C | 0.31 | 0.011 | 0.003 | 6.1E-06 | 293274 | 0.010 | 0.016 | 0.56 | 54162 |
| 5 | rs7725218 | 1282414 | G | A | 0.66 | 0.014 | 0.002 | 2.6E-08 | 294144 | 0.085 | 0.025 | 8.1E-04 | 54162 |
| 5 | rs2910949 | 39522481 | G | T | 0.35 | 0.010 | 0.002 | 3.0E-05 | 292810 | -0.002 | 0.016 | 0.89 | 54162 |
| 5 | rs11948445 | 52664796 | A | G | 0.59 | 0.011 | 0.002 | 5.8E-06 | 277410 | -0.018 | 0.017 | 0.29 | 54162 |
| 5 | rs116006942 | 53405314 | G | A | 0.94 | 0.026 | 0.005 | 2.7E-07 | 291472 | 0.060 | 0.035 | 0.08 | 54162 |
| 5 | rs16885512 | 55766621 | C | G | 0.92 | 0.021 | 0.004 | 3.4E-06 | 292065 | 0.004 | 0.028 | 0.89 | 54162 |
| 5 | rs3936511 | 55860781 | A | G | 0.81 | 0.031 | 0.003 | 3.5E-25 | 294313 | -0.036 | 0.020 | 0.07 | 54162 |
| 5 | rs12516070 | 59391636 | C | T | 0.52 | 0.013 | 0.002 | 1.8E-07 | 281085 | 0.008 | 0.016 | 0.62 | 54162 |
| 5 | rs4976033 | 67714246 | A | G | 0.60 | 0.014 | 0.002 | 3.3E-08 | 280273 | 0.000 | 0.018 | 0.99 | 54162 |
| 5 | rs2307111 | 75003678 | C | T | 0.39 | 0.019 | 0.002 | 3.4E-15 | 294473 | 0.019 | 0.016 | 0.25 | 54162 |
| 5 | rs3733890 | 78421959 | G | A | 0.71 | 0.014 | 0.003 | 9.6E-08 | 294473 | -0.016 | 0.017 | 0.35 | 54162 |
| 5 | rs115912456 | 82815158 | G | A | 0.04 | 0.031 | 0.006 | 1.4E-07 | 294454 | 0.024 | 0.035 | 0.50 | 54162 |
| 5 | rs254024 | 103944020 | G | T | 0.56 | 0.013 | 0.002 | 6.3E-08 | 294278 | -0.006 | 0.016 | 0.73 | 54162 |
| 5 | rs1862205 | 108656635 | A | G | 0.40 | 0.009 | 0.002 | 2.0E-04 | 293147 | -0.017 | 0.016 | 0.29 | 54162 |
| 5 | rs454968 | 112188456 | C | T | 0.65 | 0.009 | 0.002 | 2.0E-04 | 292782 | 0.009 | 0.016 | 0.56 | 54162 |
| 5 | rs1045241 | 118729286 | T | C | 0.27 | 0.016 | 0.003 | 8.8E-10 | 290336 | 0.007 | 0.019 | 0.70 | 54162 |

| | | | | | | | | | | | | | |
|---|---|---|---|---|---|---|---|---|---|---|---|---|---|
| 5 | rs445841 | 122331222 | G | T | 0.63 | 0.011 | 0.002 | 1.1E-05 | 294014 | -0.004 | 0.016 | 0.78 | 54162 |
| 5 | rs6893139 | 124057584 | G | A | 0.60 | 0.009 | 0.002 | 2.4E-04 | 289348 | 0.009 | 0.016 | 0.58 | 54162 |
| 5 | rs3749748 | 127350549 | T | C | 0.25 | 0.017 | 0.003 | 1.3E-09 | 292220 | 0.009 | 0.019 | 0.65 | 54162 |
| 5 | rs248653 | 130656028 | T | A | 0.96 | 0.026 | 0.006 | 3.5E-05 | 293869 | 0.093 | 0.070 | 0.19 | 54162 |
| 5 | rs72801474 | 132444128 | A | G | 0.09 | 0.021 | 0.004 | 1.6E-07 | 294473 | 0.089 | 0.039 | 0.02 | 54162 |
| 5 | rs254559 | 134444982 | C | A | 0.59 | 0.014 | 0.002 | 1.8E-09 | 293575 | 0.004 | 0.016 | 0.81 | 54162 |
| 5 | rs32578 | 149211868 | A | G | 0.31 | 0.013 | 0.003 | 8.5E-07 | 294309 | 0.005 | 0.017 | 0.76 | 54162 |
| 5 | rs2963468 | 158003020 | A | G | 0.77 | 0.019 | 0.003 | 4.1E-11 | 288262 | -0.019 | 0.018 | 0.29 | 54162 |
| 5 | rs55801554 | 158622532 | A | C | 0.24 | 0.013 | 0.003 | 2.9E-06 | 293001 | 0.009 | 0.017 | 0.59 | 54162 |
| 5 | rs2339234 | 170612546 | G | A | 0.32 | 0.012 | 0.003 | 3.4E-06 | 290248 | -0.001 | 0.017 | 0.97 | 54162 |
| 6 | rs75479205 | 7255610 | G | A | 0.19 | 0.017 | 0.003 | 7.7E-09 | 291434 | -0.025 | 0.021 | 0.22 | 54162 |
| 6 | rs1240820 | 16825137 | A | G | 0.29 | 0.012 | 0.003 | 6.1E-06 | 290362 | -0.008 | 0.017 | 0.63 | 54162 |
| 6 | rs6940493 | 20304563 | T | A | 0.69 | 0.011 | 0.003 | 8.6E-06 | 287700 | 0.010 | 0.018 | 0.59 | 54162 |
| 6 | rs68006638 | 25710571 | G | A | 0.90 | 0.020 | 0.004 | 5.8E-07 | 292496 | -0.014 | 0.034 | 0.68 | 54162 |
| 6 | rs13220570 | 26753800 | T | C | 0.87 | 0.022 | 0.004 | 1.6E-09 | 281019 | -0.013 | 0.042 | 0.76 | 54162 |
| 6 | rs142965311 | 27989252 | C | T | 0.88 | 0.025 | 0.004 | 1.7E-11 | 294277 | 0.011 | 0.032 | 0.72 | 54162 |
| 6 | rs9469583 | 33717770 | C | T | 0.52 | 0.012 | 0.002 | 2.7E-07 | 290666 | -0.015 | 0.016 | 0.34 | 54162 |
| 6 | rs2263329 | 34595543 | T | C | 0.64 | 0.029 | 0.002 | 6.4E-32 | 291493 | 0.010 | 0.017 | 0.53 | 54162 |
| 6 | rs6457807 | 35142899 | C | T | 0.17 | 0.021 | 0.003 | 5.1E-11 | 294030 | 0.004 | 0.020 | 0.86 | 54162 |
| 6 | rs1155347 | 39146230 | C | T | 0.21 | 0.014 | 0.003 | 2.6E-06 | 285395 | 0.026 | 0.019 | 0.17 | 54162 |
| 6 | rs7750688 | 41991740 | T | C | 0.76 | 0.022 | 0.003 | 1.0E-15 | 294258 | 0.026 | 0.018 | 0.15 | 54162 |
| 6 | rs3763236 | 42902508 | C | T | 0.51 | 0.014 | 0.002 | 3.4E-09 | 294473 | 0.014 | 0.016 | 0.36 | 54162 |
| 6 | rs998584 | 43757896 | C | A | 0.52 | 0.032 | 0.002 | 3.8E-42 | 291402 | 0.029 | 0.018 | 0.11 | 54162 |
| 6 | rs968050 | 98574560 | T | C | 0.48 | 0.013 | 0.002 | 1.4E-08 | 294413 | -0.026 | 0.016 | 0.10 | 54162 |
| 6 | rs7757193 | 109510972 | G | A | 0.64 | 0.020 | 0.002 | 1.6E-15 | 290391 | 0.048 | 0.016 | 3.4E-03 | 54162 |
| 6 | rs4947121 | 111834954 | T | C | 0.22 | 0.012 | 0.003 | 3.7E-05 | 293056 | 0.005 | 0.019 | 0.77 | 54162 |
| 6 | rs6934962 | 116322349 | T | C | 0.40 | 0.016 | 0.002 | 3.6E-11 | 294236 | -0.019 | 0.016 | 0.22 | 54162 |
| 6 | rs11961755 | 121766286 | G | A | 0.75 | 0.013 | 0.003 | 4.5E-06 | 290579 | -0.026 | 0.018 | 0.14 | 54162 |
| 6 | rs2781668 | 131897278 | C | T | 0.83 | 0.017 | 0.003 | 8.6E-08 | 293310 | -0.038 | 0.022 | 0.08 | 54162 |
| 6 | rs6924387 | 137082948 | A | G | 0.59 | 0.013 | 0.002 | 5.8E-08 | 286735 | 0.019 | 0.016 | 0.24 | 54162 |
| 6 | rs632057 | 139834012 | G | T | 0.63 | 0.021 | 0.002 | 1.9E-17 | 294473 | -0.001 | 0.016 | 0.94 | 54162 |
| 6 | rs62428831 | 143247161 | C | T | 0.14 | 0.020 | 0.003 | 1.4E-08 | 289228 | -0.007 | 0.023 | 0.78 | 54162 |
| 6 | rs13195251 | 153644070 | T | C | 0.28 | 0.015 | 0.003 | 1.2E-07 | 245049 | -0.025 | 0.022 | 0.26 | 54162 |
| 6 | rs9347737 | 163740322 | A | G | 0.57 | 0.014 | 0.002 | 1.8E-09 | 289696 | 0.027 | 0.016 | 0.10 | 54162 |
| 7 | rs10233430 | 1051664 | T | C | 0.57 | 0.021 | 0.002 | 1.7E-18 | 294327 | 0.005 | 0.016 | 0.74 | 54162 |
| 7 | rs10252234 | 1114381 | T | C | 0.23 | 0.014 | 0.003 | 4.7E-07 | 292737 | 0.041 | 0.019 | 0.03 | 54162 |
| 7 | rs13235365 | 6456091 | T | C | 0.27 | 0.024 | 0.003 | 5.3E-20 | 290960 | -0.031 | 0.019 | 0.11 | 54162 |
| 7 | rs10950390 | 12224708 | C | T | 0.80 | 0.015 | 0.003 | 3.5E-07 | 289681 | 0.026 | 0.019 | 0.18 | 54162 |
| 7 | rs38166 | 15889360 | C | T | 0.75 | 0.012 | 0.003 | 9.7E-06 | 289758 | -0.002 | 0.019 | 0.90 | 54162 |
| 7 | rs4410790 | 17284577 | T | C | 0.37 | 0.012 | 0.002 | 5.6E-07 | 294473 | -0.030 | 0.017 | 0.07 | 54162 |
| 7 | rs17138358 | 17920253 | G | C | 0.60 | 0.028 | 0.002 | 1.4E-30 | 293352 | 0.011 | 0.016 | 0.47 | 54162 |
| 7 | rs1534696 | 26397239 | A | C | 0.54 | 0.021 | 0.002 | 3.3E-18 | 294473 | 0.005 | 0.016 | 0.76 | 54162 |
| 7 | rs66763009 | 36193142 | T | G | 0.56 | 0.014 | 0.002 | 2.6E-09 | 285947 | 0.016 | 0.016 | 0.32 | 54162 |
| 7 | rs35580606 | 36292925 | A | G | 0.48 | 0.010 | 0.002 | 4.4E-05 | 294003 | -0.021 | 0.016 | 0.18 | 54162 |
| 7 | rs2534596 | 38277792 | G | A | 0.38 | 0.012 | 0.002 | 1.9E-06 | 288665 | -0.006 | 0.016 | 0.70 | 54162 |
| 7 | rs55935382 | 50289669 | A | C | 0.33 | 0.017 | 0.003 | 8.7E-12 | 294028 | 0.029 | 0.017 | 0.08 | 54162 |
| 7 | rs35493868 | 73039406 | G | C | 0.20 | 0.038 | 0.003 | 8.5E-38 | 293045 | 0.014 | 0.020 | 0.49 | 54162 |
| 7 | rs150300171 | 74102895 | A | C | 0.95 | 0.030 | 0.006 | 1.2E-07 | 294343 | 0.004 | 0.044 | 0.93 | 54162 |
| 7 | rs201441 | 101737327 | T | G | 0.42 | 0.011 | 0.002 | 1.8E-06 | 293121 | -0.026 | 0.016 | 0.11 | 54162 |
| 7 | rs77605964 | 106962948 | A | G | 0.23 | 0.014 | 0.003 | 6.2E-07 | 293641 | 0.008 | 0.018 | 0.65 | 54162 |
| 7 | rs12705595 | 109103912 | A | G | 0.37 | 0.011 | 0.002 | 7.3E-06 | 285694 | -0.009 | 0.016 | 0.59 | 54162 |
| 7 | rs4731701 | 130430930 | T | C | 0.49 | 0.032 | 0.002 | 1.8E-41 | 291715 | 0.003 | 0.016 | 0.85 | 54162 |
| 7 | rs73151974 | 134669523 | C | T | 0.86 | 0.017 | 0.003 | 7.8E-07 | 289711 | -0.033 | 0.022 | 0.13 | 54162 |
| 7 | rs34940374 | 150331377 | G | A | 0.82 | 0.015 | 0.003 | 1.8E-06 | 292896 | -0.008 | 0.020 | 0.68 | 54162 |
| 8 | rs330042 | 9105172 | A | G | 0.26 | 0.014 | 0.003 | 1.8E-07 | 291996 | -0.016 | 0.018 | 0.38 | 54162 |
| 8 | rs62491176 | 9859913 | C | T | 0.14 | 0.020 | 0.003 | 9.9E-09 | 291636 | -0.043 | 0.023 | 0.07 | 54162 |
| 8 | rs9657541 | 10643164 | C | T | 0.80 | 0.014 | 0.003 | 1.2E-06 | 294473 | 0.058 | 0.020 | 3.6E-03 | 54162 |
| 8 | rs62486442 | 12623463 | G | A | 0.67 | 0.014 | 0.003 | 5.0E-08 | 277726 | 0.022 | 0.019 | 0.25 | 54162 |
| 8 | rs67934334 | 14291955 | A | G | 0.63 | 0.012 | 0.002 | 2.7E-06 | 286008 | 0.012 | 0.017 | 0.48 | 54162 |
| 8 | rs79153732 | 19662937 | C | T | 0.98 | 0.097 | 0.009 | 8.3E-26 | 294259 | 0.001 | 0.056 | 0.98 | 54162 |
| 8 | rs343 | 19810787 | A | C | 0.08 | 0.140 | 0.004 | 2.5E-227 | 291884 | 0.023 | 0.029 | 0.43 | 54162 |

| | | | | | | | | | | | | | |
|---|---|---|---|---|---|---|---|---|---|---|---|---|---|
| 8 | rs308 | 19817476 | G | T | 0.02 | 0.136 | 0.008 | 1.3E-60 | 294197 | 0.091 | 0.085 | 0.28 | 54162 |
| 8 | rs142288236 | 19845612 | C | T | 0.99 | 0.080 | 0.010 | 1.5E-15 | 293852 | 0.043 | 0.071 | 0.54 | 54162 |
| 8 | rs80005209 | 19872339 | T | G | 0.97 | 0.147 | 0.007 | 7.6E-97 | 293531 | 0.035 | 0.049 | 0.48 | 54162 |
| 8 | rs61435086 | 19910292 | C | T | 0.01 | 0.090 | 0.011 | 1.4E-16 | 294390 | 0.049 | 0.108 | 0.65 | 54162 |
| 8 | rs75662196 | 19941656 | C | G | 0.03 | 0.075 | 0.007 | 9.0E-24 | 291758 | -0.046 | 0.088 | 0.60 | 54162 |
| 8 | rs7826177 | 34406540 | C | T | 0.64 | 0.014 | 0.002 | 6.2E-09 | 290408 | 0.017 | 0.018 | 0.34 | 54162 |
| 8 | rs67344323 | 64653461 | T | C | 0.74 | 0.015 | 0.003 | 3.6E-08 | 293142 | 0.007 | 0.018 | 0.69 | 54162 |
| 8 | rs10504477 | 71338185 | T | C | 0.59 | 0.017 | 0.002 | 3.6E-12 | 290695 | -0.014 | 0.016 | 0.37 | 54162 |
| 8 | rs1431659 | 73439070 | G | A | 0.73 | 0.013 | 0.003 | 1.7E-06 | 292704 | 0.003 | 0.018 | 0.88 | 54162 |
| 8 | rs61596977 | 95997165 | C | T | 0.86 | 0.014 | 0.003 | 6.2E-05 | 292546 | 0.063 | 0.023 | 0.01 | 54162 |
| 8 | rs2247355 | 103876780 | T | C | 0.18 | 0.019 | 0.003 | 9.9E-10 | 293784 | -0.001 | 0.020 | 0.96 | 54162 |
| 8 | rs6987377 | 106590684 | G | A | 0.58 | 0.011 | 0.002 | 6.5E-06 | 287925 | 0.026 | 0.016 | 0.10 | 54162 |
| 8 | rs6469605 | 116601894 | T | C | 0.57 | 0.033 | 0.002 | 1.4E-42 | 292953 | 0.026 | 0.016 | 0.11 | 54162 |
| 8 | rs17740942 | 116891360 | A | T | 0.10 | 0.020 | 0.004 | 5.1E-07 | 282571 | -0.039 | 0.051 | 0.44 | 54162 |
| 8 | rs10955991 | 121867780 | T | C | 0.32 | 0.018 | 0.003 | 3.1E-13 | 293422 | -0.015 | 0.017 | 0.35 | 54162 |
| 8 | rs72647336 | 126445055 | G | A | 0.96 | 0.038 | 0.006 | 1.6E-10 | 277001 | 0.009 | 0.079 | 0.91 | 54162 |
| 8 | rs4871603 | 126480367 | T | C | 0.66 | 0.039 | 0.002 | 1.1E-54 | 293735 | 0.010 | 0.017 | 0.57 | 54162 |
| 8 | rs4871624 | 126629328 | T | G | 0.72 | 0.023 | 0.003 | 3.1E-18 | 290208 | -0.014 | 0.018 | 0.42 | 54162 |
| 8 | rs7817574 | 144302570 | C | T | 0.19 | 0.037 | 0.003 | 2.0E-33 | 294473 | 0.021 | 0.025 | 0.41 | 54162 |
| 8 | rs4875043 | 144496772 | A | C | 0.79 | 0.013 | 0.003 | 1.8E-05 | 284379 | -0.002 | 0.023 | 0.93 | 54162 |
| 9 | rs1567353 | 1033773 | C | G | 0.69 | 0.014 | 0.003 | 1.0E-07 | 291279 | 0.013 | 0.017 | 0.43 | 54162 |
| 9 | rs7039168 | 13719203 | G | A | 0.67 | 0.013 | 0.003 | 6.7E-07 | 291062 | -0.017 | 0.017 | 0.29 | 54162 |
| 9 | rs686030 | 15304782 | A | C | 0.86 | 0.048 | 0.003 | 1.8E-46 | 293933 | 0.013 | 0.022 | 0.57 | 54162 |
| 9 | rs1412234 | 28410683 | T | C | 0.67 | 0.010 | 0.003 | 1.0E-04 | 291744 | -0.037 | 0.017 | 0.03 | 54162 |
| 9 | rs7036107 | 92177897 | A | G | 0.49 | 0.011 | 0.002 | 8.8E-06 | 271687 | -0.019 | 0.016 | 0.24 | 54162 |
| 9 | rs76530346 | 94010298 | G | A | 0.91 | 0.023 | 0.004 | 2.3E-08 | 291613 | -0.021 | 0.025 | 0.41 | 54162 |
| 9 | rs12686780 | 95382297 | C | T | 0.83 | 0.017 | 0.003 | 6.4E-08 | 293575 | -0.022 | 0.020 | 0.27 | 54162 |
| 9 | rs62565259 | 102162570 | T | C | 0.17 | 0.014 | 0.003 | 4.9E-06 | 289539 | 0.017 | 0.022 | 0.45 | 54162 |
| 9 | rs2297402 | 107579880 | C | T | 0.98 | 0.066 | 0.008 | 1.5E-15 | 290523 | 0.114 | 0.067 | 0.09 | 54162 |
| 9 | rs2066714 | 107586753 | C | T | 0.13 | 0.046 | 0.004 | 1.4E-38 | 294473 | -0.082 | 0.033 | 0.01 | 54162 |
| 9 | rs11789603 | 107647019 | T | C | 0.11 | 0.068 | 0.004 | 4.4E-72 | 293708 | 0.021 | 0.026 | 0.43 | 54162 |
| 9 | rs2740488 | 107661742 | A | C | 0.74 | 0.069 | 0.003 | 1.4E-147 | 293034 | -0.052 | 0.018 | 2.9E-03 | 54162 |
| 9 | rs4979372 | 117140082 | C | T | 0.49 | 0.014 | 0.002 | 1.5E-09 | 287682 | -0.018 | 0.016 | 0.26 | 54162 |
| 9 | rs2417125 | 131562232 | A | G | 0.72 | 0.014 | 0.003 | 5.0E-08 | 294131 | -0.013 | 0.019 | 0.49 | 54162 |
| 9 | rs532436 | 136149830 | A | G | 0.19 | 0.022 | 0.003 | 2.3E-13 | 294418 | -0.002 | 0.019 | 0.90 | 54162 |
| 9 | rs2520096 | 136919416 | G | A | 0.27 | 0.012 | 0.003 | 3.6E-06 | 291694 | -0.005 | 0.018 | 0.79 | 54162 |
| 10 | rs11254464 | 17265447 | C | T | 0.42 | 0.015 | 0.002 | 1.2E-09 | 292070 | 0.002 | 0.016 | 0.91 | 54162 |
| 10 | rs11009262 | 33448764 | G | T | 0.94 | 0.027 | 0.005 | 7.6E-08 | 294473 | 0.042 | 0.036 | 0.24 | 54162 |
| 10 | rs2804894 | 33647091 | A | G | 0.74 | 0.018 | 0.003 | 5.6E-11 | 288902 | 0.001 | 0.018 | 0.96 | 54162 |
| 10 | rs12411959 | 34015681 | A | T | 0.78 | 0.016 | 0.003 | 5.2E-08 | 292074 | -0.022 | 0.022 | 0.31 | 54162 |
| 10 | rs11239536 | 45978598 | A | T | 0.24 | 0.028 | 0.003 | 1.2E-23 | 292859 | 0.005 | 0.018 | 0.79 | 54162 |
| 10 | rs10826337 | 61409469 | G | A | 0.59 | 0.011 | 0.002 | 3.1E-06 | 293569 | -0.013 | 0.016 | 0.42 | 54162 |
| 10 | rs10761737 | 65052205 | C | T | 0.42 | 0.015 | 0.002 | 3.4E-10 | 293660 | -0.022 | 0.016 | 0.16 | 54162 |
| 10 | rs72805692 | 71099109 | A | G | 0.88 | 0.017 | 0.004 | 2.3E-06 | 293467 | 0.010 | 0.027 | 0.71 | 54162 |
| 10 | rs11000468 | 74711376 | T | C | 0.25 | 0.012 | 0.003 | 3.4E-05 | 280261 | -0.008 | 0.020 | 0.68 | 54162 |
| 10 | rs7903537 | 76847490 | C | T | 0.48 | 0.012 | 0.002 | 9.6E-07 | 291488 | 0.011 | 0.016 | 0.50 | 54162 |
| 10 | rs703966 | 80954251 | A | G | 0.42 | 0.015 | 0.002 | 2.8E-10 | 292776 | -0.021 | 0.016 | 0.18 | 54162 |
| 10 | rs2068888 | 94839642 | A | G | 0.45 | 0.017 | 0.002 | 1.9E-13 | 294473 | -0.021 | 0.016 | 0.20 | 54162 |
| 10 | rs10786114 | 95309022 | T | C | 0.88 | 0.022 | 0.004 | 1.4E-09 | 293920 | -0.015 | 0.026 | 0.56 | 54162 |
| 10 | rs577525 | 99769388 | T | C | 0.44 | 0.015 | 0.002 | 4.8E-10 | 293244 | -0.001 | 0.016 | 0.96 | 54162 |
| 10 | rs2862954 | 101912064 | T | C | 0.50 | 0.014 | 0.002 | 1.3E-09 | 294473 | 0.026 | 0.016 | 0.12 | 54162 |
| 10 | rs2792751 | 113940329 | T | C | 0.27 | 0.036 | 0.003 | 1.6E-41 | 294473 | 0.017 | 0.017 | 0.32 | 54162 |
| 10 | rs12411732 | 113977211 | G | A | 0.86 | 0.033 | 0.003 | 1.5E-21 | 288231 | 0.004 | 0.023 | 0.88 | 54162 |
| 10 | rs1970811 | 126696496 | T | C | 0.54 | 0.012 | 0.002 | 1.9E-07 | 292669 | 0.010 | 0.016 | 0.50 | 54162 |
| 11 | rs16928809 | 2936952 | G | A | 0.91 | 0.029 | 0.004 | 1.9E-12 | 291753 | -0.042 | 0.028 | 0.13 | 54162 |
| 11 | rs11601507 | 5701074 | C | A | 0.93 | 0.024 | 0.005 | 1.3E-07 | 294473 | 0.025 | 0.044 | 0.58 | 54162 |
| 11 | rs2218793 | 10380828 | C | A | 0.72 | 0.018 | 0.003 | 9.7E-12 | 294148 | 0.021 | 0.017 | 0.21 | 54162 |
| 11 | rs61884030 | 14444543 | C | T | 0.14 | 0.020 | 0.003 | 4.2E-09 | 292050 | -0.048 | 0.024 | 0.04 | 54162 |
| 11 | rs79634051 | 14561945 | C | G | 0.03 | 0.042 | 0.007 | 2.8E-09 | 294473 | 0.009 | 0.057 | 0.87 | 54162 |
| 11 | rs17309930 | 27748493 | C | A | 0.79 | 0.022 | 0.003 | 1.8E-14 | 294473 | -0.029 | 0.020 | 0.14 | 54162 |
| 11 | rs567056 | 30435051 | G | T | 0.62 | 0.009 | 0.002 | 3.4E-04 | 289822 | -0.011 | 0.016 | 0.52 | 54162 |

| | | | | | | | | | | | | | |
|---|---|---|---|---|---|---|---|---|---|---|---|---|---|
| 11 | rs4755720 | 43628749 | T | C | 0.61 | 0.012 | 0.002 | 3.6E-07 | 290414 | -0.014 | 0.016 | 0.38 | 54162 |
| 11 | rs77403571 | 45913607 | A | G | 0.06 | 0.036 | 0.005 | 6.2E-13 | 289786 | 0.100 | 0.042 | 0.02 | 54162 |
| 11 | rs75817747 | 46231010 | T | C | 0.03 | 0.040 | 0.007 | 1.7E-08 | 293692 | -0.016 | 0.052 | 0.77 | 54162 |
| 11 | rs112192770 | 46991808 | T | C | 0.03 | 0.031 | 0.007 | 1.0E-05 | 275521 | 0.029 | 0.044 | 0.51 | 54162 |
| 11 | rs1052373 | 47354787 | T | C | 0.32 | 0.044 | 0.003 | 1.1E-69 | 294473 | -0.003 | 0.018 | 0.87 | 54162 |
| 11 | rs145276599 | 48444711 | C | T | 0.11 | 0.050 | 0.004 | 4.9E-40 | 294199 | 0.020 | 0.033 | 0.55 | 54162 |
| 11 | rs73457437 | 49500475 | G | A | 0.97 | 0.034 | 0.007 | 4.0E-07 | 294444 | 0.032 | 0.047 | 0.49 | 54162 |
| 11 | rs12224170 | 49616839 | A | G | 0.13 | 0.042 | 0.003 | 2.0E-33 | 294106 | 0.030 | 0.025 | 0.22 | 54162 |
| 11 | rs12422125 | 50619511 | A | G | 0.11 | 0.046 | 0.004 | 6.3E-35 | 290015 | 0.026 | 0.028 | 0.36 | 54162 |
| 11 | rs11228871 | 54846440 | C | A | 0.13 | 0.041 | 0.003 | 1.5E-32 | 293059 | 0.017 | 0.029 | 0.57 | 54162 |
| 11 | rs11228212 | 56156771 | G | C | 0.13 | 0.041 | 0.004 | 2.0E-30 | 293979 | 0.036 | 0.027 | 0.18 | 54162 |
| 11 | rs77916918 | 56628086 | A | G | 0.03 | 0.034 | 0.006 | 1.7E-07 | 294473 | 0.041 | 0.052 | 0.43 | 54162 |
| 11 | rs10896581 | 56964224 | G | A | 0.88 | 0.019 | 0.004 | 2.3E-07 | 294011 | 0.014 | 0.024 | 0.56 | 54162 |
| 11 | rs12803463 | 58376120 | A | G | 0.08 | 0.023 | 0.004 | 1.4E-07 | 293971 | 0.063 | 0.032 | 0.05 | 54162 |
| 11 | rs174566 | 61592362 | A | G | 0.65 | 0.057 | 0.002 | 1.3E-118 | 293939 | 0.009 | 0.016 | 0.58 | 54162 |
| 11 | rs11231138 | 62316195 | C | G | 0.63 | 0.012 | 0.002 | 3.4E-07 | 293187 | 0.012 | 0.016 | 0.47 | 54162 |
| 11 | rs56271783 | 64004723 | G | C | 0.96 | 0.052 | 0.006 | 8.4E-19 | 292157 | -0.033 | 0.042 | 0.44 | 54162 |
| 11 | rs10750766 | 65473798 | C | A | 0.29 | 0.020 | 0.003 | 2.8E-14 | 292827 | -0.004 | 0.017 | 0.80 | 54162 |
| 11 | rs4930352 | 66066993 | T | G | 0.50 | 0.018 | 0.002 | 8.1E-14 | 272099 | -0.008 | 0.016 | 0.63 | 54162 |
| 11 | rs34695471 | 67076064 | A | G | 0.06 | 0.019 | 0.005 | 1.1E-04 | 288696 | 0.071 | 0.040 | 0.08 | 54162 |
| 11 | rs11605837 | 68597886 | G | T | 0.69 | 0.020 | 0.003 | 7.8E-15 | 292499 | -0.008 | 0.017 | 0.63 | 54162 |
| 11 | rs559355 | 75451281 | A | T | 0.84 | 0.034 | 0.003 | 1.0E-25 | 294254 | -0.008 | 0.021 | 0.68 | 54162 |
| 11 | rs2155220 | 76266172 | C | T | 0.56 | 0.010 | 0.002 | 3.4E-05 | 294473 | -0.018 | 0.016 | 0.26 | 54162 |
| 11 | rs11021232 | 95320808 | T | C | 0.82 | 0.020 | 0.003 | 7.1E-11 | 292030 | -0.024 | 0.021 | 0.25 | 54162 |
| 11 | rs12146566 | 103871404 | A | C | 0.80 | 0.016 | 0.003 | 7.0E-08 | 292633 | 0.001 | 0.020 | 0.96 | 54162 |
| 11 | rs17566828 | 111644401 | A | G | 0.04 | 0.026 | 0.006 | 9.7E-06 | 294303 | 0.021 | 0.045 | 0.64 | 54162 |
| 11 | rs964184 | 116648917 | C | G | 0.87 | 0.108 | 0.003 | 2.2E-213 | 294473 | -0.021 | 0.023 | 0.36 | 54162 |
| 11 | rs514924 | 118346863 | A | G | 0.10 | 0.014 | 0.004 | 3.0E-04 | 293384 | -0.045 | 0.026 | 0.08 | 54162 |
| 11 | rs7925100 | 118941596 | G | A | 0.60 | 0.019 | 0.002 | 9.0E-16 | 294354 | -0.006 | 0.016 | 0.73 | 54162 |
| 11 | rs58473820 | 122514403 | T | C | 0.38 | 0.028 | 0.002 | 1.6E-30 | 292688 | 0.032 | 0.016 | 0.04 | 54162 |
| 12 | rs10774439 | 6731818 | A | G | 0.82 | 0.020 | 0.003 | 5.5E-11 | 286328 | 0.032 | 0.022 | 0.15 | 54162 |
| 12 | rs2302367 | 6861043 | T | C | 0.51 | 0.010 | 0.002 | 4.7E-05 | 286419 | 0.003 | 0.017 | 0.86 | 54162 |
| 12 | rs11045171 | 20470199 | G | A | 0.20 | 0.031 | 0.003 | 8.1E-26 | 291037 | 0.019 | 0.020 | 0.34 | 54162 |
| 12 | rs7488780 | 20579392 | C | G | 0.20 | 0.019 | 0.003 | 2.4E-10 | 292479 | 0.023 | 0.022 | 0.28 | 54162 |
| 12 | rs12814794 | 26440698 | A | G | 0.76 | 0.014 | 0.003 | 8.8E-07 | 292197 | -0.001 | 0.018 | 0.98 | 54162 |
| 12 | rs7316878 | 33715933 | C | T | 0.43 | 0.013 | 0.003 | 1.6E-06 | 238777 | -0.006 | 0.017 | 0.72 | 54162 |
| 12 | rs61926301 | 51157863 | T | G | 0.41 | 0.012 | 0.002 | 2.1E-07 | 293678 | -0.010 | 0.016 | 0.53 | 54162 |
| 12 | rs11170516 | 53752692 | G | A | 0.85 | 0.018 | 0.003 | 2.4E-08 | 293134 | -0.064 | 0.023 | 0.00 | 54162 |
| 12 | rs11171710 | 56368078 | G | A | 0.55 | 0.015 | 0.002 | 2.8E-10 | 286310 | 0.007 | 0.016 | 0.68 | 54162 |
| 12 | rs58298943 | 57391292 | T | C | 0.08 | 0.022 | 0.004 | 2.6E-07 | 294027 | -0.026 | 0.028 | 0.35 | 54162 |
| 12 | rs1252425 | 67651972 | G | C | 0.66 | 0.014 | 0.003 | 6.1E-08 | 289992 | 0.005 | 0.017 | 0.77 | 54162 |
| 12 | rs10879184 | 71106648 | C | T | 0.50 | 0.011 | 0.002 | 2.6E-06 | 285795 | -0.010 | 0.016 | 0.53 | 54162 |
| 12 | rs2645979 | 84017043 | A | G | 0.36 | 0.009 | 0.002 | 2.7E-04 | 294153 | -0.008 | 0.016 | 0.63 | 54162 |
| 12 | rs1012306 | 101888063 | T | C | 0.56 | 0.010 | 0.002 | 3.5E-05 | 292545 | -0.017 | 0.016 | 0.29 | 54162 |
| 12 | rs7308864 | 109871179 | G | A | 0.52 | 0.025 | 0.002 | 1.6E-26 | 292414 | 0.008 | 0.016 | 0.61 | 54162 |
| 12 | rs3184504 | 111884608 | C | T | 0.52 | 0.027 | 0.002 | 1.5E-29 | 294473 | 0.025 | 0.016 | 0.11 | 54162 |
| 12 | rs11066320 | 112906415 | G | A | 0.58 | 0.021 | 0.002 | 1.6E-18 | 292454 | 0.009 | 0.016 | 0.56 | 54162 |
| 12 | rs10774579 | 121405210 | T | C | 0.52 | 0.015 | 0.002 | 7.6E-11 | 293307 | -0.009 | 0.016 | 0.56 | 54162 |
| 12 | rs11043221 | 122301044 | C | A | 0.30 | 0.012 | 0.003 | 3.9E-06 | 285892 | -0.025 | 0.019 | 0.20 | 54162 |
| 12 | rs113740515 | 123199410 | A | G | 0.21 | 0.039 | 0.003 | 3.7E-41 | 294199 | -0.049 | 0.024 | 0.04 | 54162 |
| 12 | rs1054852 | 124496316 | G | A | 0.39 | 0.030 | 0.003 | 2.7E-29 | 243204 | 0.004 | 0.017 | 0.83 | 54162 |
| 12 | rs11057468 | 124637757 | T | C | 0.39 | 0.015 | 0.002 | 4.4E-09 | 282532 | 0.008 | 0.017 | 0.64 | 54162 |
| 12 | rs10846690 | 125086417 | C | T | 0.85 | 0.021 | 0.003 | 6.6E-10 | 286674 | 0.005 | 0.023 | 0.84 | 54162 |
| 12 | rs921919 | 125265201 | G | A | 0.32 | 0.042 | 0.003 | 1.6E-58 | 275331 | 0.004 | 0.018 | 0.83 | 54162 |
| 12 | rs7136506 | 125326153 | T | C | 0.79 | 0.037 | 0.003 | 2.7E-36 | 280966 | 0.008 | 0.021 | 0.71 | 54162 |
| 12 | rs4078216 | 125434716 | A | G | 0.24 | 0.014 | 0.003 | 5.3E-07 | 293774 | 0.038 | 0.019 | 0.05 | 54162 |
| 13 | rs117230571 | 41689067 | A | G | 0.93 | 0.023 | 0.005 | 2.8E-07 | 291330 | 0.011 | 0.032 | 0.73 | 54162 |
| 13 | rs9526023 | 45524626 | A | G | 0.03 | 0.030 | 0.007 | 1.1E-05 | 294092 | 0.071 | 0.049 | 0.15 | 54162 |
| 13 | rs183078 | 49513352 | G | A | 0.40 | 0.014 | 0.002 | 7.4E-09 | 294244 | 0.005 | 0.016 | 0.77 | 54162 |
| 13 | rs549058 | 51201045 | T | G | 0.12 | 0.018 | 0.004 | 6.1E-07 | 294353 | -0.029 | 0.024 | 0.23 | 54162 |
| 14 | rs1760940 | 20938251 | C | A | 0.25 | 0.010 | 0.003 | 1.7E-04 | 294473 | -0.019 | 0.020 | 0.35 | 54162 |

| | | | | | | | | | | | | | |
|---|---|---|---|---|---|---|---|---|---|---|---|---|---|
| 14 | rs1955512 | 33175822 | A | G | 0.58 | 0.012 | 0.002 | 5.9E-07 | 273542 | 0.023 | 0.017 | 0.16 | 54162 |
| 14 | rs35828909 | 52574576 | A | C | 0.45 | 0.012 | 0.002 | 1.9E-06 | 277596 | 0.005 | 0.016 | 0.77 | 54162 |
| 14 | rs72729582 | 69149372 | G | A | 0.07 | 0.028 | 0.005 | 2.0E-09 | 289873 | -0.009 | 0.034 | 0.80 | 54162 |
| 14 | rs13379043 | 74250126 | C | T | 0.27 | 0.019 | 0.003 | 3.6E-12 | 276648 | 0.030 | 0.018 | 0.10 | 54162 |
| 14 | rs8014289 | 75377352 | G | A | 0.56 | 0.015 | 0.002 | 1.3E-10 | 292638 | 0.043 | 0.016 | 0.01 | 54162 |
| 14 | rs9646167 | 81635888 | C | T | 0.48 | 0.014 | 0.002 | 5.0E-09 | 289324 | 0.022 | 0.016 | 0.16 | 54162 |
| 14 | rs17124112 | 88605509 | C | A | 0.92 | 0.020 | 0.004 | 3.4E-06 | 293681 | -0.017 | 0.028 | 0.55 | 54162 |
| 14 | rs3825669 | 89804276 | G | A | 0.81 | 0.016 | 0.003 | 4.4E-08 | 291536 | -0.023 | 0.021 | 0.26 | 54162 |
| 14 | rs11622947 | 98396293 | C | T | 0.46 | 0.010 | 0.002 | 3.2E-05 | 293155 | -0.004 | 0.016 | 0.81 | 54162 |
| 14 | rs61993685 | 100765823 | C | T | 0.08 | 0.021 | 0.004 | 2.1E-06 | 294473 | 0.048 | 0.054 | 0.37 | 54162 |
| 14 | rs7158166 | 103241799 | C | T | 0.60 | 0.014 | 0.002 | 1.8E-08 | 289133 | 0.026 | 0.016 | 0.11 | 54162 |
| 14 | rs2498786 | 105262368 | C | G | 0.38 | 0.026 | 0.002 | 2.6E-26 | 291671 | -0.032 | 0.019 | 0.09 | 54162 |
| 15 | rs1818917 | 23941678 | T | C | 0.51 | 0.012 | 0.002 | 7.4E-07 | 292058 | 0.028 | 0.017 | 0.11 | 54162 |
| 15 | rs28510484 | 31637569 | G | C | 0.83 | 0.018 | 0.003 | 5.0E-09 | 292153 | 0.046 | 0.023 | 0.05 | 54162 |
| 15 | rs4924471 | 40744046 | T | G | 0.08 | 0.021 | 0.004 | 1.7E-06 | 291545 | 0.033 | 0.028 | 0.24 | 54162 |
| 15 | rs7170463 | 41888918 | G | A | 0.31 | 0.022 | 0.003 | 1.3E-17 | 292611 | -0.033 | 0.017 | 0.05 | 54162 |
| 15 | rs147525635 | 43468698 | G | A | 0.44 | 0.012 | 0.002 | 4.4E-07 | 292345 | -0.015 | 0.017 | 0.40 | 54162 |
| 15 | rs150844304 | 43726625 | A | C | 0.98 | 0.095 | 0.008 | 1.3E-35 | 294397 | 0.021 | 0.045 | 0.64 | 54162 |
| 15 | rs4273010 | 44947434 | T | C | 0.98 | 0.073 | 0.008 | 6.0E-21 | 292997 | 0.023 | 0.062 | 0.71 | 54162 |
| 15 | rs2414178 | 53001538 | C | T | 0.22 | 0.011 | 0.003 | 1.1E-04 | 282059 | 0.010 | 0.019 | 0.62 | 54162 |
| 15 | rs10162642 | 58577163 | G | A | 0.79 | 0.048 | 0.003 | 4.0E-60 | 290483 | 0.052 | 0.019 | 0.01 | 54162 |
| 15 | rs1800588 | 58723675 | T | C | 0.22 | 0.121 | 0.003 | 0.0E+00 | 294473 | -0.003 | 0.019 | 0.89 | 54162 |
| 15 | rs2245477 | 61948435 | C | A | 0.63 | 0.009 | 0.002 | 1.4E-04 | 292886 | 0.017 | 0.016 | 0.30 | 54162 |
| 15 | rs3803501 | 63352092 | A | G | 0.57 | 0.019 | 0.002 | 2.9E-15 | 293480 | 0.032 | 0.016 | 0.04 | 54162 |
| 15 | rs10438354 | 67336207 | G | C | 0.35 | 0.012 | 0.003 | 2.5E-06 | 272896 | 0.016 | 0.020 | 0.40 | 54162 |
| 15 | rs28362901 | 74712937 | C | A | 0.91 | 0.027 | 0.004 | 6.5E-11 | 293208 | -0.024 | 0.029 | 0.40 | 54162 |
| 15 | rs2663924 | 81392903 | G | T | 0.76 | 0.012 | 0.003 | 1.9E-05 | 289056 | 0.018 | 0.018 | 0.32 | 54162 |
| 16 | rs12921195 | 4677604 | C | A | 0.87 | 0.021 | 0.004 | 6.6E-09 | 274775 | -0.124 | 0.057 | 0.03 | 54162 |
| 16 | rs12928099 | 15150505 | A | C | 0.30 | 0.024 | 0.003 | 3.0E-20 | 293591 | 0.000 | 0.018 | 0.98 | 54162 |
| 16 | rs7188071 | 28917644 | C | T | 0.64 | 0.009 | 0.002 | 1.4E-04 | 293872 | 0.014 | 0.018 | 0.43 | 54162 |
| 16 | rs3814883 | 29994922 | C | T | 0.52 | 0.017 | 0.002 | 9.6E-13 | 290530 | 0.029 | 0.016 | 0.07 | 54162 |
| 16 | rs57348955 | 31185882 | A | G | 0.39 | 0.012 | 0.003 | 1.4E-06 | 278139 | 0.052 | 0.019 | 0.01 | 54162 |
| 16 | rs11075985 | 53805207 | C | A | 0.58 | 0.015 | 0.002 | 2.6E-10 | 294354 | 0.000 | 0.016 | 0.99 | 54162 |
| 16 | rs75152587 | 56579961 | G | T | 0.99 | 0.103 | 0.011 | 1.2E-21 | 293907 | 0.072 | 0.107 | 0.50 | 54162 |
| 16 | rs142493909 | 56679888 | C | T | 0.02 | 0.072 | 0.009 | 6.3E-16 | 293866 | 0.001 | 0.093 | 0.99 | 54162 |
| 16 | rs79616633 | 56801047 | C | T | 0.98 | 0.038 | 0.008 | 1.6E-06 | 294232 | 0.083 | 0.060 | 0.17 | 54162 |
| 16 | rs79600951 | 56834254 | C | G | 0.91 | 0.119 | 0.004 | 3.7E-188 | 294209 | 0.017 | 0.029 | 0.55 | 54162 |
| 16 | rs9989419 | 56985139 | G | A | 0.61 | 0.154 | 0.002 | 0.0E+00 | 294473 | 0.008 | 0.016 | 0.62 | 54162 |
| 16 | rs11645157 | 57308423 | T | G | 0.50 | 0.022 | 0.002 | 1.2E-20 | 293945 | 0.005 | 0.016 | 0.73 | 54162 |
| 16 | rs138026745 | 57682700 | A | G | 0.02 | 0.038 | 0.008 | 1.5E-06 | 290826 | -0.066 | 0.070 | 0.35 | 54162 |
| 16 | rs34830321 | 58616997 | C | T | 0.99 | 0.045 | 0.011 | 2.9E-05 | 292425 | 0.121 | 0.092 | 0.19 | 54162 |
| 16 | rs6499102 | 66906224 | G | A | 0.94 | 0.039 | 0.005 | 5.4E-15 | 293992 | -0.011 | 0.044 | 0.80 | 54162 |
| 16 | rs77234291 | 66911898 | G | A | 0.02 | 0.054 | 0.009 | 5.5E-09 | 294473 | -0.004 | 0.084 | 0.96 | 54162 |
| 16 | rs55781197 | 67940350 | G | A | 0.11 | 0.061 | 0.004 | 3.5E-62 | 294389 | 0.002 | 0.026 | 0.93 | 54162 |
| 16 | rs77631377 | 69049293 | C | T | 0.96 | 0.040 | 0.006 | 1.5E-11 | 292331 | -0.042 | 0.038 | 0.28 | 54162 |
| 16 | rs4788446 | 71856727 | C | T | 0.75 | 0.014 | 0.003 | 1.2E-07 | 291734 | -0.011 | 0.018 | 0.56 | 54162 |
| 16 | rs2925979 | 81534790 | C | T | 0.70 | 0.035 | 0.003 | 2.5E-43 | 294473 | -0.006 | 0.018 | 0.72 | 54162 |
| 16 | rs79311290 | 85150163 | A | G | 0.91 | 0.022 | 0.004 | 3.7E-07 | 270937 | 0.006 | 0.046 | 0.90 | 54162 |
| 16 | rs12926854 | 85951258 | G | A | 0.27 | 0.013 | 0.003 | 5.6E-07 | 290024 | 0.005 | 0.018 | 0.79 | 54162 |
| 16 | rs11640494 | 88029685 | G | A | 0.54 | 0.015 | 0.002 | 2.8E-10 | 290172 | -0.001 | 0.021 | 0.97 | 54162 |
| 17 | rs141062196 | 495327 | G | A | 0.81 | 0.015 | 0.003 | 2.3E-07 | 293412 | -0.010 | 0.020 | 0.62 | 54162 |
| 17 | rs8081548 | 7438834 | A | T | 0.66 | 0.018 | 0.003 | 9.1E-13 | 290873 | -0.014 | 0.018 | 0.42 | 54162 |
| 17 | rs2585398 | 8054860 | C | A | 0.45 | 0.014 | 0.002 | 1.2E-08 | 288924 | -0.021 | 0.016 | 0.20 | 54162 |
| 17 | rs11658872 | 17425069 | C | T | 0.94 | 0.033 | 0.005 | 7.2E-11 | 294009 | -0.006 | 0.035 | 0.87 | 54162 |
| 17 | rs2071379 | 26695832 | A | G | 0.41 | 0.015 | 0.002 | 6.6E-10 | 293328 | -0.001 | 0.016 | 0.96 | 54162 |
| 17 | rs2011614 | 28781792 | G | A | 0.65 | 0.014 | 0.002 | 2.8E-08 | 293799 | 0.008 | 0.016 | 0.64 | 54162 |
| 17 | rs11658786 | 37815899 | A | G | 0.67 | 0.030 | 0.002 | 1.8E-33 | 294455 | 0.016 | 0.016 | 0.33 | 54162 |
| 17 | rs2314338 | 38344485 | T | C | 0.73 | 0.013 | 0.003 | 6.6E-07 | 294473 | -0.009 | 0.024 | 0.72 | 54162 |
| 17 | rs34138141 | 40781561 | G | T | 0.72 | 0.021 | 0.003 | 6.9E-16 | 293730 | -0.029 | 0.018 | 0.10 | 54162 |
| 17 | rs74456742 | 42191796 | A | G | 0.03 | 0.041 | 0.007 | 1.0E-09 | 289473 | -0.095 | 0.053 | 0.07 | 54162 |
| 17 | rs117499775 | 44078618 | T | C | 0.96 | 0.034 | 0.006 | 1.0E-08 | 294473 | 0.202 | 0.117 | 0.08 | 54162 |

| | | | | | | | | | | | | | |
|---|---|---|---|---|---|---|---|---|---|---|---|---|---|
| 17 | rs16969990 | 46230891 | T | C | 0.07 | 0.026 | 0.005 | 1.5E-08 | 294341 | 0.045 | 0.034 | 0.19 | 54162 |
| 17 | rs595767 | 46957987 | A | G | 0.48 | 0.013 | 0.002 | 1.9E-08 | 291985 | -0.005 | 0.016 | 0.77 | 54162 |
| 17 | rs3794752 | 53382829 | C | T | 0.28 | 0.011 | 0.003 | 5.8E-05 | 285795 | 0.014 | 0.018 | 0.42 | 54162 |
| 17 | rs11653260 | 66464683 | A | G | 0.79 | 0.011 | 0.003 | 1.1E-04 | 290380 | -0.010 | 0.020 | 0.60 | 54162 |
| 17 | rs16975758 | 68419330 | A | G | 0.38 | 0.010 | 0.002 | 3.3E-05 | 290144 | 0.009 | 0.017 | 0.60 | 54162 |
| 17 | rs4969141 | 76391653 | T | C | 0.49 | 0.029 | 0.002 | 2.7E-34 | 293534 | 0.032 | 0.016 | 0.05 | 54162 |
| 17 | rs7218647 | 76769605 | A | G | 0.56 | 0.012 | 0.002 | 8.0E-07 | 291176 | -0.010 | 0.016 | 0.53 | 54162 |
| 18 | rs4800356 | 19658460 | C | T | 0.72 | 0.016 | 0.003 | 6.1E-08 | 249455 | 0.004 | 0.022 | 0.86 | 54162 |
| 18 | rs2435307 | 21127910 | T | C | 0.49 | 0.017 | 0.002 | 3.0E-13 | 292988 | 0.017 | 0.016 | 0.30 | 54162 |
| 18 | rs680321 | 29797958 | C | T | 0.45 | 0.010 | 0.002 | 3.8E-05 | 293877 | -0.001 | 0.016 | 0.95 | 54162 |
| 18 | rs62092069 | 40735531 | A | G | 0.37 | 0.010 | 0.002 | 3.9E-05 | 294473 | 0.010 | 0.016 | 0.54 | 54162 |
| 18 | rs150237291 | 46781014 | C | T | 0.02 | 0.048 | 0.008 | 3.5E-09 | 293975 | -0.061 | 0.082 | 0.46 | 54162 |
| 18 | rs77960347 | 47109955 | G | A | 0.01 | 0.277 | 0.010 | 5.0E-164 | 294473 | 0.024 | 0.058 | 0.67 | 54162 |
| 18 | rs8086351 | 47171888 | G | C | 0.82 | 0.085 | 0.003 | 4.9E-166 | 293914 | -0.014 | 0.021 | 0.52 | 54162 |
| 18 | rs2298624 | 47429022 | T | C | 0.13 | 0.031 | 0.003 | 1.1E-19 | 294473 | 0.013 | 0.023 | 0.57 | 54162 |
| 18 | rs41292412 | 56118358 | C | T | 0.99 | 0.063 | 0.011 | 8.5E-09 | 293529 | -0.092 | 0.087 | 0.29 | 54162 |
| 18 | rs11664369 | 57739072 | C | T | 0.73 | 0.025 | 0.003 | 2.0E-20 | 293801 | 0.027 | 0.017 | 0.12 | 54162 |
| 18 | rs73455693 | 58027496 | A | G | 0.04 | 0.029 | 0.006 | 3.9E-06 | 292306 | 0.001 | 0.053 | 0.99 | 54162 |
| 19 | rs59737437 | 2671100 | T | C | 0.27 | 0.010 | 0.003 | 3.5E-04 | 286727 | -0.047 | 0.020 | 0.02 | 54162 |
| 19 | rs12975319 | 3414088 | G | A | 0.70 | 0.012 | 0.003 | 2.6E-06 | 288459 | 0.012 | 0.018 | 0.50 | 54162 |
| 19 | rs2289863 | 4028783 | C | T | 0.25 | 0.018 | 0.003 | 6.9E-11 | 289336 | -0.006 | 0.019 | 0.77 | 54162 |
| 19 | rs11878235 | 7976698 | A | G | 0.60 | 0.014 | 0.002 | 2.6E-08 | 285315 | 0.026 | 0.020 | 0.20 | 54162 |
| 19 | rs35137994 | 8429066 | T | C | 0.05 | 0.045 | 0.005 | 4.3E-18 | 294049 | -0.040 | 0.034 | 0.25 | 54162 |
| 19 | rs76213248 | 11269893 | T | C | 0.41 | 0.014 | 0.002 | 3.1E-09 | 289686 | -0.032 | 0.017 | 0.06 | 54162 |
| 19 | rs737338 | 11347657 | C | T | 0.97 | 0.089 | 0.006 | 5.6E-43 | 294416 | -0.023 | 0.042 | 0.59 | 54162 |
| 19 | rs7251640 | 18614935 | C | T | 0.19 | 0.014 | 0.003 | 6.6E-06 | 289976 | 0.046 | 0.022 | 0.04 | 54162 |
| 19 | rs62102718 | 33891013 | A | T | 0.72 | 0.023 | 0.003 | 1.0E-18 | 292428 | -0.005 | 0.018 | 0.76 | 54162 |
| 19 | rs34559316 | 38863464 | C | A | 0.81 | 0.017 | 0.003 | 3.0E-08 | 291819 | 0.046 | 0.023 | 0.05 | 54162 |
| 19 | rs15052 | 41813375 | C | T | 0.18 | 0.015 | 0.003 | 6.7E-07 | 293140 | -0.016 | 0.025 | 0.52 | 54162 |
| 19 | rs4760 | 44153100 | A | G | 0.84 | 0.020 | 0.003 | 8.5E-10 | 294473 | 0.057 | 0.037 | 0.12 | 54162 |
| 19 | rs11880219 | 46385997 | T | A | 0.82 | 0.020 | 0.003 | 2.7E-11 | 294473 | 0.014 | 0.022 | 0.52 | 54162 |
| 19 | rs12150914 | 47563418 | T | C | 0.40 | 0.014 | 0.002 | 5.0E-09 | 284252 | 0.003 | 0.019 | 0.89 | 54162 |
| 19 | rs367070 | 54800500 | G | A | 0.23 | 0.042 | 0.003 | 1.5E-49 | 291358 | 0.024 | 0.026 | 0.36 | 54162 |
| 19 | rs36092527 | 56181408 | T | C | 0.88 | 0.018 | 0.004 | 1.2E-06 | 294473 | -0.014 | 0.027 | 0.60 | 54162 |
| 19 | rs8102873 | 57488423 | C | T | 0.42 | 0.010 | 0.002 | 2.6E-05 | 294473 | 0.015 | 0.017 | 0.39 | 54162 |
| 20 | rs144033177 | 571467 | A | C | 0.99 | 0.062 | 0.010 | 2.3E-10 | 292823 | -0.052 | 0.087 | 0.55 | 54162 |
| 20 | rs1132274 | 17596155 | C | A | 0.84 | 0.019 | 0.003 | 1.8E-09 | 294473 | -0.053 | 0.022 | 0.02 | 54162 |
| 20 | rs1884589 | 21885619 | A | C | 0.43 | 0.013 | 0.002 | 1.4E-08 | 294295 | 0.023 | 0.016 | 0.14 | 54162 |
| 20 | rs6059958 | 30143278 | T | C | 0.17 | 0.012 | 0.003 | 1.1E-04 | 289058 | -0.025 | 0.021 | 0.22 | 54162 |
| 20 | rs13041173 | 32542814 | A | G | 0.66 | 0.012 | 0.002 | 2.9E-06 | 292573 | 0.020 | 0.016 | 0.22 | 54162 |
| 20 | rs3736802 | 33604042 | C | T | 0.52 | 0.017 | 0.002 | 1.2E-12 | 294410 | 0.045 | 0.016 | 0.00 | 54162 |
| 20 | rs1800961 | 43042364 | C | T | 0.97 | 0.134 | 0.007 | 9.5E-88 | 294473 | -0.060 | 0.047 | 0.20 | 54162 |
| 20 | rs6073958 | 44551855 | T | C | 0.80 | 0.063 | 0.003 | 2.7E-99 | 294065 | -0.027 | 0.021 | 0.19 | 54162 |
| 20 | rs6131012 | 44728661 | G | A | 0.40 | 0.014 | 0.002 | 4.2E-09 | 285578 | 0.008 | 0.016 | 0.61 | 54162 |
| 20 | rs13042367 | 46219058 | G | A | 0.28 | 0.014 | 0.003 | 6.3E-08 | 294342 | -0.003 | 0.017 | 0.88 | 54162 |
| 20 | rs3859588 | 46476143 | T | A | 0.79 | 0.012 | 0.003 | 1.9E-05 | 287752 | 0.004 | 0.021 | 0.86 | 54162 |
| 20 | rs6021914 | 51031169 | C | T | 0.37 | 0.013 | 0.002 | 1.3E-07 | 293848 | -0.021 | 0.016 | 0.20 | 54162 |
| 20 | rs6123685 | 55836040 | A | G | 0.25 | 0.016 | 0.003 | 4.9E-09 | 294473 | 0.055 | 0.022 | 0.01 | 54162 |
| 20 | rs76602912 | 57459868 | T | C | 0.98 | 0.045 | 0.008 | 8.2E-09 | 294473 | 0.209 | 0.085 | 0.01 | 54162 |
| 20 | rs8126001 | 62711459 | T | C | 0.49 | 0.012 | 0.002 | 1.6E-07 | 291152 | 0.009 | 0.019 | 0.65 | 54162 |
| 21 | rs8127283 | 16403394 | G | A | 0.51 | 0.014 | 0.003 | 1.8E-06 | 192169 | -0.013 | 0.025 | 0.61 | 54162 |
| 21 | rs3746915 | 43718792 | G | A | 0.58 | 0.012 | 0.002 | 3.2E-07 | 292415 | -0.003 | 0.016 | 0.83 | 54162 |
| 21 | rs235314 | 46271452 | C | T | 0.47 | 0.019 | 0.002 | 3.0E-15 | 293779 | -0.025 | 0.017 | 0.13 | 54162 |
| 22 | rs2256609 | 21925017 | A | G | 0.81 | 0.031 | 0.003 | 1.3E-25 | 294167 | 0.079 | 0.021 | 0.00 | 54162 |
| 22 | rs55652051 | 29905277 | A | G | 0.77 | 0.013 | 0.003 | 6.4E-06 | 289558 | 0.039 | 0.019 | 0.04 | 54162 |
| 22 | rs9608972 | 30931307 | T | C | 0.76 | 0.016 | 0.003 | 2.8E-09 | 293215 | 0.009 | 0.019 | 0.62 | 54162 |
| 22 | rs9610329 | 36042986 | C | T | 0.57 | 0.012 | 0.002 | 5.6E-07 | 272509 | -0.006 | 0.018 | 0.73 | 54162 |
| 22 | rs9622830 | 38954703 | C | G | 0.65 | 0.016 | 0.002 | 2.2E-10 | 293842 | 0.026 | 0.016 | 0.11 | 54162 |
| 22 | rs6002946 | 43150523 | T | G | 0.68 | 0.008 | 0.003 | 1.4E-03 | 290791 | 0.017 | 0.018 | 0.35 | 54162 |
| 22 | rs738409 | 44324727 | C | G | 0.78 | 0.014 | 0.003 | 2.3E-06 | 294473 | 0.021 | 0.019 | 0.27 | 54162 |

**Supplementary table 4:** List of 361 SNPs used as instrumental variables for triglycerides, and their association with triglycerides and dementia.

| CHR | rsID | POS | Effect allele | Other allele | EAF | BETA Trg | SE Trg | Pval Trg | n Trg | BETA dementia | SE dementia | Pval dementia | n dementia |
|---|---|---|---|---|---|---|---|---|---|---|---|---|---|
| 1 | rs880315 | 10796866 | T | C | 0.66 | 0.011 | 0.003 | 1.6E-05 | 304735 | 0.008 | 0.017 | 0.63 | 54162 |
| 1 | rs61780049 | 39363294 | G | A | 0.15 | 0.016 | 0.003 | 5.4E-06 | 313211 | -0.017 | 0.023 | 0.45 | 54162 |
| 1 | rs11206374 | 40048009 | A | G | 0.23 | 0.025 | 0.003 | 1.1E-17 | 313888 | 0.017 | 0.019 | 0.39 | 54162 |
| 1 | rs2131311 | 40435999 | A | G | 0.28 | 0.014 | 0.003 | 3.5E-07 | 300594 | 0.054 | 0.021 | 0.01 | 54162 |
| 1 | rs72904737 | 51351846 | G | A | 0.91 | 0.024 | 0.004 | 2.2E-08 | 313081 | 0.036 | 0.027 | 0.18 | 54162 |
| 1 | rs213494 | 54877103 | T | C | 0.65 | 0.016 | 0.003 | 1.0E-09 | 312535 | -0.021 | 0.017 | 0.22 | 54162 |
| 1 | rs9970140 | 61684288 | A | G | 0.92 | 0.025 | 0.005 | 9.4E-08 | 313570 | 0.013 | 0.032 | 0.69 | 54162 |
| 1 | rs9436661 | 62904575 | T | G | 0.65 | 0.080 | 0.003 | 0.00 | 313601 | -0.033 | 0.017 | 0.05 | 54162 |
| 1 | rs1365297 | 93858292 | A | G | 0.82 | 0.019 | 0.003 | 2.3E-09 | 314000 | 0.033 | 0.022 | 0.13 | 54162 |
| 1 | rs1938566 | 98478981 | C | T | 0.17 | 0.021 | 0.003 | 3.2E-10 | 313251 | 0.014 | 0.022 | 0.51 | 54162 |
| 1 | rs320369 | 118143517 | A | G | 0.32 | 0.013 | 0.003 | 2.3E-06 | 307805 | -0.002 | 0.017 | 0.93 | 54162 |
| 1 | rs10797996 | 172353311 | C | T | 0.43 | 0.014 | 0.002 | 3.1E-08 | 313837 | 0.002 | 0.016 | 0.92 | 54162 |
| 1 | rs6700266 | 178508930 | G | A | 0.66 | 0.013 | 0.003 | 1.9E-07 | 312917 | 0.010 | 0.016 | 0.54 | 54162 |
| 1 | rs11240358 | 205070573 | A | G | 0.39 | 0.012 | 0.003 | 2.6E-06 | 313731 | 0.003 | 0.016 | 0.87 | 54162 |
| 1 | rs10863828 | 210592392 | T | G | 0.76 | 0.013 | 0.003 | 3.8E-06 | 313261 | 0.006 | 0.018 | 0.74 | 54162 |
| 1 | rs11118310 | 219637671 | T | A | 0.59 | 0.018 | 0.002 | 1.3E-13 | 313726 | -0.024 | 0.016 | 0.13 | 54162 |
| 1 | rs61830291 | 221001142 | C | A | 0.10 | 0.028 | 0.004 | 8.0E-12 | 314332 | 0.026 | 0.033 | 0.42 | 54162 |
| 1 | rs6690181 | 228056868 | T | C | 0.63 | 0.014 | 0.003 | 1.8E-07 | 293302 | -0.022 | 0.018 | 0.23 | 54162 |
| 1 | rs11122450 | 230301811 | T | G | 0.39 | 0.049 | 0.003 | 3.7E-83 | 313312 | -0.009 | 0.017 | 0.60 | 54162 |
| 1 | rs114052230 | 230410811 | C | T | 0.85 | 0.024 | 0.004 | 1.9E-11 | 291340 | 0.001 | 0.024 | 0.96 | 54162 |
| 2 | rs3820897 | 3642361 | C | T | 0.82 | 0.016 | 0.003 | 1.2E-06 | 308211 | 0.003 | 0.022 | 0.90 | 54162 |
| 2 | rs676210 | 21231524 | G | A | 0.80 | 0.076 | 0.003 | 0.00 | 314332 | 0.052 | 0.019 | 0.01 | 54162 |
| 2 | rs58953077 | 21510295 | T | C | 0.39 | 0.017 | 0.003 | 8.5E-11 | 303081 | -0.010 | 0.017 | 0.56 | 54162 |
| 2 | rs11903847 | 25592918 | T | C | 0.33 | 0.012 | 0.003 | 7.1E-06 | 295931 | -0.012 | 0.018 | 0.49 | 54162 |
| 2 | rs935168 | 26914787 | A | G | 0.66 | 0.013 | 0.003 | 1.2E-06 | 311668 | -0.033 | 0.017 | 0.05 | 54162 |
| 2 | rs4665972 | 27598097 | T | C | 0.39 | 0.100 | 0.003 | 0.0E+00 | 308059 | -0.003 | 0.017 | 0.88 | 54162 |
| 2 | rs75225803 | 28455336 | C | T | 0.92 | 0.022 | 0.005 | 2.3E-06 | 302016 | 0.033 | 0.035 | 0.33 | 54162 |
| 2 | rs10176110 | 28610627 | C | T | 0.12 | 0.029 | 0.004 | 1.1E-14 | 314332 | -0.026 | 0.035 | 0.47 | 54162 |
| 2 | rs58839393 | 43490619 | T | A | 0.16 | 0.016 | 0.003 | 2.1E-06 | 309121 | -0.006 | 0.021 | 0.76 | 54162 |
| 2 | rs17326656 | 48962291 | T | G | 0.24 | 0.016 | 0.003 | 2.4E-08 | 310949 | 0.004 | 0.019 | 0.85 | 54162 |
| 2 | rs10180284 | 50716016 | C | T | 0.52 | 0.013 | 0.002 | 9.6E-08 | 310445 | 0.013 | 0.016 | 0.40 | 54162 |
| 2 | rs7424120 | 59313974 | C | T | 0.40 | 0.013 | 0.003 | 9.6E-08 | 310004 | 0.027 | 0.016 | 0.10 | 54162 |
| 2 | rs1009360 | 65276049 | T | C | 0.58 | 0.020 | 0.002 | 3.8E-16 | 312502 | 0.018 | 0.016 | 0.26 | 54162 |
| 2 | rs10172544 | 85788270 | C | A | 0.59 | 0.012 | 0.002 | 2.1E-06 | 314262 | -0.007 | 0.016 | 0.67 | 54162 |
| 2 | rs6708784 | 111927379 | A | G | 0.50 | 0.014 | 0.002 | 2.3E-08 | 312860 | -0.002 | 0.016 | 0.89 | 54162 |
| 2 | rs77631110 | 119752450 | C | A | 0.02 | 0.057 | 0.010 | 1.6E-08 | 313983 | 0.008 | 0.067 | 0.91 | 54162 |
| 2 | rs954244 | 121309231 | G | C | 0.25 | 0.014 | 0.003 | 5.4E-07 | 314332 | 0.002 | 0.019 | 0.92 | 54162 |
| 2 | rs4662414 | 145818432 | A | G | 0.55 | 0.009 | 0.002 | 1.6E-04 | 313092 | -0.018 | 0.016 | 0.26 | 54162 |
| 2 | rs4128205 | 165467068 | A | C | 0.51 | 0.012 | 0.002 | 1.4E-06 | 307071 | 0.005 | 0.017 | 0.76 | 54162 |
| 2 | rs13389219 | 165528876 | C | T | 0.61 | 0.037 | 0.003 | 1.7E-48 | 314213 | 0.027 | 0.016 | 0.09 | 54162 |
| 2 | rs76172517 | 169514699 | T | C | 0.89 | 0.017 | 0.004 | 2.6E-05 | 309836 | 0.017 | 0.026 | 0.51 | 54162 |
| 2 | rs12472667 | 171629063 | G | C | 0.37 | 0.016 | 0.003 | 2.9E-10 | 309377 | -0.005 | 0.016 | 0.76 | 54162 |
| 2 | rs72917533 | 175238924 | T | C | 0.82 | 0.018 | 0.003 | 2.3E-08 | 311024 | 0.028 | 0.021 | 0.17 | 54162 |
| 2 | rs2110690 | 202185132 | G | A | 0.51 | 0.013 | 0.002 | 3.5E-07 | 311769 | -0.015 | 0.016 | 0.34 | 54162 |
| 2 | rs3731696 | 203431804 | G | A | 0.12 | 0.021 | 0.004 | 4.0E-08 | 314332 | 0.022 | 0.023 | 0.35 | 54162 |
| 2 | rs2382825 | 219184275 | C | T | 0.38 | 0.012 | 0.003 | 3.1E-06 | 311926 | -0.007 | 0.018 | 0.70 | 54162 |
| 2 | rs2943645 | 227099180 | T | C | 0.65 | 0.041 | 0.003 | 1.1E-58 | 314332 | 0.011 | 0.016 | 0.52 | 54162 |
| 2 | rs57074291 | 227229344 | C | G | 0.74 | 0.010 | 0.003 | 3.8E-04 | 312911 | -0.004 | 0.018 | 0.81 | 54162 |
| 2 | rs7596814 | 230128204 | G | T | 0.71 | 0.014 | 0.003 | 1.4E-07 | 312592 | -0.007 | 0.017 | 0.68 | 54162 |
| 2 | rs4675812 | 242395674 | G | A | 0.41 | 0.012 | 0.002 | 6.9E-07 | 313361 | 0.006 | 0.017 | 0.74 | 54162 |
| 3 | rs9812100 | 4763301 | G | A | 0.51 | 0.015 | 0.002 | 3.1E-09 | 312663 | -0.012 | 0.016 | 0.44 | 54162 |
| 3 | rs6798755 | 12139092 | C | T | 0.93 | 0.030 | 0.005 | 1.7E-09 | 313927 | -0.003 | 0.036 | 0.94 | 54162 |
| 3 | rs3103310 | 12473045 | G | A | 0.23 | 0.022 | 0.003 | 1.1E-13 | 295914 | -0.009 | 0.021 | 0.66 | 54162 |
| 3 | rs2455821 | 15681940 | A | C | 0.27 | 0.014 | 0.003 | 3.4E-07 | 311578 | 0.008 | 0.018 | 0.66 | 54162 |
| 3 | rs6792725 | 24520283 | A | G | 0.30 | 0.016 | 0.003 | 1.1E-08 | 280928 | -0.021 | 0.017 | 0.22 | 54162 |
| 3 | rs9831084 | 37025661 | T | C | 0.54 | 0.012 | 0.002 | 1.5E-06 | 311722 | -0.010 | 0.016 | 0.53 | 54162 |
| 3 | rs6800707 | 52516293 | G | C | 0.81 | 0.030 | 0.003 | 5.9E-22 | 312696 | -0.025 | 0.020 | 0.22 | 54162 |
| 3 | rs6805924 | 69879670 | T | G | 0.43 | 0.008 | 0.002 | 7.0E-04 | 313996 | 0.011 | 0.016 | 0.47 | 54162 |

| | | | | | | | | | | | | | |
|---|---|---|---|---|---|---|---|---|---|---|---|---|---|
| 3 | rs79983121 | 127306462 | T | C | 0.20 | 0.016 | 0.003 | 1.0E-07 | 313405 | 0.018 | 0.020 | 0.36 | 54162 |
| 3 | rs684773 | 135956305 | C | A | 0.77 | 0.028 | 0.003 | 1.0E-22 | 314332 | -0.015 | 0.018 | 0.40 | 54162 |
| 3 | rs62271373 | 150066540 | A | T | 0.06 | 0.043 | 0.005 | 9.9E-16 | 308645 | -0.037 | 0.035 | 0.30 | 54162 |
| 3 | rs9817452 | 156795414 | G | T | 0.61 | 0.016 | 0.003 | 7.7E-11 | 312290 | -0.016 | 0.017 | 0.33 | 54162 |
| 3 | rs79287178 | 172294500 | A | G | 0.03 | 0.053 | 0.008 | 1.6E-11 | 307356 | -0.111 | 0.096 | 0.25 | 54162 |
| 3 | rs2194411 | 185548663 | G | A | 0.88 | 0.017 | 0.004 | 8.1E-06 | 307742 | 0.017 | 0.034 | 0.60 | 54162 |
| 3 | rs1152847 | 188421362 | G | A | 0.64 | 0.011 | 0.003 | 2.7E-05 | 312088 | 0.009 | 0.016 | 0.59 | 54162 |
| 3 | rs2342371 | 196187608 | G | A | 0.27 | 0.015 | 0.003 | 1.2E-07 | 310961 | 0.000 | 0.019 | 0.99 | 54162 |
| 4 | rs13101828 | 965720 | A | G | 0.55 | 0.011 | 0.002 | 1.5E-05 | 310417 | -0.040 | 0.016 | 0.01 | 54162 |
| 4 | rs13108218 | 3443931 | A | G | 0.38 | 0.031 | 0.003 | 1.2E-34 | 304621 | -0.015 | 0.018 | 0.39 | 54162 |
| 4 | rs4450871 | 4990298 | A | G | 0.56 | 0.013 | 0.002 | 3.5E-07 | 314332 | -0.008 | 0.027 | 0.78 | 54162 |
| 4 | rs7681288 | 15096523 | A | G | 0.67 | 0.012 | 0.003 | 7.0E-06 | 312311 | -0.004 | 0.016 | 0.81 | 54162 |
| 4 | rs71603401 | 18034463 | G | A | 0.13 | 0.025 | 0.004 | 2.1E-11 | 308015 | 0.031 | 0.024 | 0.19 | 54162 |
| 4 | rs73243877 | 26047616 | G | A | 0.17 | 0.025 | 0.003 | 1.9E-14 | 314332 | -0.001 | 0.021 | 0.97 | 54162 |
| 4 | rs12504746 | 39646631 | C | T | 0.81 | 0.012 | 0.003 | 7.2E-05 | 313358 | 0.002 | 0.020 | 0.90 | 54162 |
| 4 | rs2237029 | 55535336 | G | A | 0.40 | 0.015 | 0.003 | 6.2E-09 | 306765 | -0.022 | 0.016 | 0.18 | 54162 |
| 4 | rs7439032 | 86931091 | T | C | 0.20 | 0.018 | 0.003 | 2.8E-09 | 314332 | -0.047 | 0.025 | 0.06 | 54162 |
| 4 | rs3775228 | 87985166 | T | C | 0.40 | 0.037 | 0.003 | 8.6E-48 | 309844 | 0.002 | 0.016 | 0.89 | 54162 |
| 4 | rs3822072 | 89741269 | A | G | 0.45 | 0.016 | 0.002 | 6.7E-11 | 314332 | 0.010 | 0.016 | 0.54 | 54162 |
| 4 | rs1126673 | 100045616 | T | C | 0.70 | 0.014 | 0.003 | 1.9E-07 | 314332 | 0.050 | 0.017 | 0.00 | 54162 |
| 4 | rs2035816 | 100508556 | A | G | 0.92 | 0.028 | 0.004 | 1.9E-10 | 314015 | -0.015 | 0.030 | 0.61 | 54162 |
| 4 | rs13107325 | 103188709 | T | C | 0.07 | 0.033 | 0.005 | 9.0E-13 | 314332 | 0.051 | 0.031 | 0.10 | 54162 |
| 4 | rs78025076 | 110569620 | T | C | 0.02 | 0.048 | 0.009 | 1.7E-08 | 314332 | 0.253 | 0.135 | 0.06 | 54162 |
| 4 | rs1347188 | 124743259 | G | A | 0.24 | 0.013 | 0.003 | 3.8E-06 | 310954 | 0.020 | 0.018 | 0.26 | 54162 |
| 4 | rs11100083 | 157682598 | T | C | 0.78 | 0.015 | 0.003 | 2.2E-07 | 312739 | 0.003 | 0.018 | 0.85 | 54162 |
| 5 | rs7735249 | 53310139 | G | C | 0.11 | 0.025 | 0.004 | 3.1E-10 | 311583 | -0.010 | 0.025 | 0.70 | 54162 |
| 5 | rs72644085 | 55783832 | T | C | 0.86 | 0.022 | 0.004 | 1.5E-09 | 313681 | -0.001 | 0.025 | 0.97 | 54162 |
| 5 | rs3936511 | 55860781 | G | A | 0.19 | 0.044 | 0.003 | 4.5E-45 | 314162 | 0.036 | 0.020 | 0.07 | 54162 |
| 5 | rs11746801 | 55990342 | G | A | 0.36 | 0.012 | 0.003 | 7.6E-06 | 306696 | 0.000 | 0.017 | 0.99 | 54162 |
| 5 | rs37538 | 57610069 | G | C | 0.40 | 0.017 | 0.003 | 2.9E-11 | 309083 | -0.010 | 0.016 | 0.53 | 54162 |
| 5 | rs4976033 | 67714246 | G | A | 0.40 | 0.021 | 0.003 | 5.0E-16 | 299086 | 0.000 | 0.018 | 0.99 | 54162 |
| 5 | rs1316753 | 78531337 | G | C | 0.61 | 0.013 | 0.003 | 1.1E-07 | 313660 | 0.032 | 0.016 | 0.04 | 54162 |
| 5 | rs34580448 | 82810884 | T | C | 0.96 | 0.033 | 0.006 | 8.7E-08 | 314046 | -0.018 | 0.033 | 0.59 | 54162 |
| 5 | rs7704653 | 90255685 | G | A | 0.73 | 0.014 | 0.003 | 1.4E-06 | 303345 | -0.018 | 0.017 | 0.29 | 54162 |
| 5 | rs325485 | 103995368 | A | G | 0.40 | 0.010 | 0.003 | 8.1E-05 | 306650 | 0.015 | 0.016 | 0.35 | 54162 |
| 5 | rs7714361 | 112490629 | C | A | 0.23 | 0.016 | 0.003 | 1.3E-07 | 307313 | 0.005 | 0.019 | 0.81 | 54162 |
| 5 | rs1045241 | 118729286 | C | T | 0.73 | 0.022 | 0.003 | 6.0E-15 | 309930 | -0.007 | 0.019 | 0.70 | 54162 |
| 5 | rs193735 | 130672958 | A | G | 0.04 | 0.028 | 0.006 | 1.3E-05 | 313622 | -0.059 | 0.046 | 0.20 | 54162 |
| 5 | rs72801474 | 132444128 | G | A | 0.91 | 0.029 | 0.004 | 1.1E-11 | 314332 | -0.089 | 0.039 | 0.02 | 54162 |
| 5 | rs112424890 | 140933792 | T | C | 0.18 | 0.019 | 0.003 | 3.9E-09 | 312242 | -0.034 | 0.023 | 0.13 | 54162 |
| 5 | rs76957426 | 144501147 | T | C | 0.30 | 0.012 | 0.003 | 1.2E-05 | 312288 | -0.028 | 0.017 | 0.10 | 54162 |
| 5 | rs245051 | 149345975 | A | G | 0.59 | 0.012 | 0.002 | 3.4E-06 | 312478 | 0.017 | 0.016 | 0.28 | 54162 |
| 5 | rs7244 | 153800513 | A | G | 0.17 | 0.016 | 0.003 | 8.2E-07 | 314239 | 0.006 | 0.021 | 0.76 | 54162 |
| 5 | rs6882076 | 156390297 | C | T | 0.64 | 0.036 | 0.003 | 1.5E-45 | 314332 | 0.008 | 0.017 | 0.62 | 54162 |
| 5 | rs1030472 | 157994544 | G | A | 0.21 | 0.023 | 0.003 | 2.9E-14 | 307893 | 0.014 | 0.019 | 0.46 | 54162 |
| 5 | rs55646464 | 173324971 | T | G | 0.30 | 0.012 | 0.003 | 5.7E-06 | 312497 | 0.008 | 0.017 | 0.62 | 54162 |
| 5 | rs62397245 | 176750688 | G | C | 0.22 | 0.015 | 0.003 | 3.0E-07 | 309657 | -0.024 | 0.019 | 0.21 | 54162 |
| 6 | rs78588343 | 7249460 | G | A | 0.82 | 0.016 | 0.003 | 1.2E-06 | 313499 | 0.029 | 0.021 | 0.17 | 54162 |
| 6 | rs6924805 | 18747705 | G | T | 0.41 | 0.009 | 0.002 | 1.6E-04 | 311473 | -0.008 | 0.016 | 0.61 | 54162 |
| 6 | rs4134963 | 20486798 | C | T | 0.81 | 0.016 | 0.003 | 3.2E-07 | 309887 | -0.025 | 0.021 | 0.22 | 54162 |
| 6 | rs806973 | 26148326 | A | G | 0.61 | 0.015 | 0.003 | 7.8E-09 | 314332 | 0.006 | 0.016 | 0.71 | 54162 |
| 6 | rs7749305 | 27446566 | T | C | 0.87 | 0.024 | 0.004 | 4.3E-11 | 314332 | -0.007 | 0.030 | 0.81 | 54162 |
| 6 | rs4714001 | 36638175 | A | G | 0.64 | 0.010 | 0.003 | 1.0E-04 | 313429 | -0.010 | 0.016 | 0.53 | 54162 |
| 6 | rs998584 | 43757896 | A | C | 0.48 | 0.039 | 0.002 | 2.7E-57 | 311057 | -0.029 | 0.018 | 0.11 | 54162 |
| 6 | rs729761 | 43804571 | G | T | 0.72 | 0.019 | 0.003 | 6.3E-12 | 303405 | -0.020 | 0.019 | 0.28 | 54162 |
| 6 | rs6458869 | 52630269 | C | A | 0.36 | 0.021 | 0.003 | 5.3E-16 | 313434 | 0.013 | 0.017 | 0.44 | 54162 |
| 6 | rs138191773 | 72522321 | G | A | 0.99 | 0.048 | 0.011 | 4.2E-06 | 311307 | 0.228 | 0.147 | 0.12 | 54162 |
| 6 | rs2499797 | 96848669 | G | A | 0.16 | 0.014 | 0.003 | 1.7E-05 | 314332 | -0.026 | 0.021 | 0.22 | 54162 |
| 6 | rs9496567 | 100602753 | G | A | 0.76 | 0.013 | 0.003 | 9.9E-06 | 312211 | 0.008 | 0.019 | 0.67 | 54162 |
| 6 | rs6913325 | 106378009 | G | T | 0.55 | 0.014 | 0.002 | 3.0E-08 | 312524 | -0.005 | 0.016 | 0.74 | 54162 |
| 6 | rs62427982 | 107437166 | C | T | 0.68 | 0.013 | 0.003 | 1.2E-06 | 309308 | -0.031 | 0.022 | 0.17 | 54162 |

| | | | | | | | | | | | | | |
|---|---|---|---|---|---|---|---|---|---|---|---|---|---|
| 6 | rs9480889 | 109189021 | G | C | 0.78 | 0.019 | 0.003 | 4.5E-10 | 312123 | -0.003 | 0.019 | 0.87 | 54162 |
| 6 | rs4947121 | 111834954 | C | T | 0.78 | 0.015 | 0.003 | 1.5E-07 | 312818 | -0.005 | 0.019 | 0.77 | 54162 |
| 6 | rs9375694 | 130356608 | A | G | 0.70 | 0.017 | 0.003 | 8.3E-11 | 313173 | 0.002 | 0.018 | 0.91 | 54162 |
| 6 | rs2277083 | 133827354 | A | G | 0.44 | 0.016 | 0.002 | 1.8E-10 | 314332 | 0.001 | 0.016 | 0.97 | 54162 |
| 6 | rs9376511 | 140611419 | A | G | 0.80 | 0.014 | 0.003 | 7.6E-06 | 314101 | 0.015 | 0.019 | 0.44 | 54162 |
| 6 | rs1281978 | 153473232 | C | T | 0.47 | 0.012 | 0.002 | 9.5E-07 | 313571 | 0.010 | 0.016 | 0.52 | 54162 |
| 6 | rs73025562 | 160562481 | A | G | 0.24 | 0.015 | 0.003 | 1.3E-07 | 311715 | -0.023 | 0.018 | 0.22 | 54162 |
| 6 | rs77009508 | 161004972 | G | A | 0.08 | 0.045 | 0.005 | 8.0E-22 | 314332 | -0.081 | 0.033 | 0.01 | 54162 |
| 6 | rs1835346 | 161162290 | A | G | 0.98 | 0.040 | 0.008 | 9.0E-07 | 313482 | 0.028 | 0.057 | 0.62 | 54162 |
| 6 | rs4709746 | 164133001 | C | T | 0.87 | 0.022 | 0.004 | 1.2E-09 | 310998 | -0.013 | 0.024 | 0.60 | 54162 |
| 7 | rs71538127 | 1010801 | G | C | 0.12 | 0.016 | 0.004 | 2.1E-05 | 313760 | -0.015 | 0.028 | 0.59 | 54162 |
| 7 | rs852388 | 5574239 | C | G | 0.21 | 0.016 | 0.003 | 2.0E-07 | 304194 | 0.043 | 0.020 | 0.03 | 54162 |
| 7 | rs6968865 | 17287269 | T | A | 0.63 | 0.017 | 0.003 | 1.2E-11 | 314332 | 0.032 | 0.016 | 0.05 | 54162 |
| 7 | rs10242866 | 17920613 | T | C | 0.40 | 0.017 | 0.003 | 4.9E-12 | 313306 | -0.012 | 0.016 | 0.45 | 54162 |
| 7 | rs4722551 | 25991826 | T | C | 0.84 | 0.037 | 0.003 | 1.5E-27 | 314332 | 0.022 | 0.022 | 0.31 | 54162 |
| 7 | rs2699805 | 26394297 | G | A | 0.60 | 0.018 | 0.003 | 5.2E-13 | 307367 | 0.004 | 0.016 | 0.81 | 54162 |
| 7 | rs498475 | 28256240 | G | A | 0.37 | 0.012 | 0.003 | 3.4E-06 | 312976 | -0.046 | 0.016 | 4.0E-03 | 54162 |
| 7 | rs11980456 | 29319249 | A | G | 0.29 | 0.011 | 0.003 | 1.3E-04 | 301018 | 0.024 | 0.017 | 0.16 | 54162 |
| 7 | rs12669911 | 32262377 | A | C | 0.38 | 0.009 | 0.003 | 2.9E-04 | 310401 | 0.027 | 0.017 | 0.11 | 54162 |
| 7 | rs1799831 | 44199142 | T | C | 0.16 | 0.022 | 0.003 | 5.2E-11 | 312717 | -0.031 | 0.022 | 0.16 | 54162 |
| 7 | rs62459095 | 44267538 | C | T | 0.95 | 0.031 | 0.005 | 1.3E-08 | 304556 | -0.152 | 0.062 | 0.01 | 54162 |
| 7 | rs36104871 | 71808047 | G | T | 0.97 | 0.066 | 0.007 | 5.3E-19 | 311433 | 0.013 | 0.061 | 0.82 | 54162 |
| 7 | rs139453187 | 72155834 | C | T | 0.07 | 0.024 | 0.005 | 6.1E-07 | 313975 | -0.064 | 0.059 | 0.27 | 54162 |
| 7 | rs2240466 | 72856269 | G | A | 0.88 | 0.123 | 0.004 | 0.00 | 314332 | -0.024 | 0.024 | 0.32 | 54162 |
| 7 | rs72555385 | 73123473 | G | A | 0.05 | 0.065 | 0.006 | 5.6E-30 | 313738 | 0.048 | 0.038 | 0.21 | 54162 |
| 7 | rs111914893 | 74002323 | T | C | 0.05 | 0.024 | 0.006 | 3.8E-05 | 308347 | -0.023 | 0.054 | 0.67 | 54162 |
| 7 | rs6465120 | 76036364 | A | G | 0.51 | 0.014 | 0.002 | 6.8E-09 | 313717 | -0.040 | 0.016 | 0.01 | 54162 |
| 7 | rs12530679 | 106632113 | A | G | 0.52 | 0.012 | 0.003 | 5.0E-06 | 296018 | 0.016 | 0.016 | 0.31 | 54162 |
| 7 | rs41785 | 116486020 | C | A | 0.58 | 0.014 | 0.002 | 7.1E-09 | 313482 | -0.012 | 0.016 | 0.44 | 54162 |
| 7 | rs4731701 | 130430930 | C | T | 0.50 | 0.034 | 0.002 | 2.3E-44 | 311390 | -0.003 | 0.016 | 0.85 | 54162 |
| 7 | rs62473520 | 130583442 | T | C | 0.93 | 0.022 | 0.005 | 3.5E-06 | 305337 | -0.012 | 0.044 | 0.78 | 54162 |
| 7 | rs56321085 | 150542048 | A | G | 0.08 | 0.019 | 0.004 | 2.1E-05 | 311840 | 0.016 | 0.028 | 0.56 | 54162 |
| 8 | rs2980755 | 8363683 | A | G | 0.54 | 0.025 | 0.002 | 3.5E-24 | 305216 | 0.007 | 0.016 | 0.67 | 54162 |
| 8 | rs7018436 | 10574848 | C | T | 0.31 | 0.016 | 0.003 | 8.3E-10 | 311892 | -0.009 | 0.017 | 0.59 | 54162 |
| 8 | rs34893217 | 16279180 | G | T | 0.90 | 0.020 | 0.004 | 6.3E-07 | 311604 | 0.004 | 0.025 | 0.89 | 54162 |
| 8 | rs383091 | 17884606 | C | T | 0.63 | 0.014 | 0.003 | 6.8E-08 | 313605 | 0.003 | 0.016 | 0.86 | 54162 |
| 8 | rs1495741 | 18272881 | G | A | 0.22 | 0.039 | 0.003 | 1.6E-40 | 314332 | -0.004 | 0.019 | 0.83 | 54162 |
| 8 | rs79153732 | 19662937 | T | C | 0.02 | 0.084 | 0.010 | 1.5E-18 | 314107 | -0.001 | 0.056 | 0.98 | 54162 |
| 8 | rs343 | 19810787 | C | A | 0.92 | 0.148 | 0.005 | 0.00 | 311548 | -0.023 | 0.029 | 0.43 | 54162 |
| 8 | rs308 | 19817476 | T | G | 0.98 | 0.168 | 0.009 | 1.2E-84 | 314035 | -0.091 | 0.085 | 0.28 | 54162 |
| 8 | rs11781692 | 19848117 | A | C | 0.01 | 0.084 | 0.010 | 9.4E-16 | 313718 | -0.043 | 0.071 | 0.54 | 54162 |
| 8 | rs7000494 | 19873502 | C | G | 0.03 | 0.138 | 0.007 | 4.0E-79 | 313164 | -0.035 | 0.049 | 0.48 | 54162 |
| 8 | rs78376313 | 19917471 | C | T | 0.03 | 0.045 | 0.007 | 1.6E-09 | 309010 | -0.021 | 0.047 | 0.65 | 54162 |
| 8 | rs113266765 | 37402248 | C | T | 0.97 | 0.032 | 0.007 | 6.6E-06 | 313797 | -0.035 | 0.077 | 0.65 | 54162 |
| 8 | rs36061954 | 38329650 | T | C | 0.40 | 0.010 | 0.003 | 3.8E-05 | 312944 | 0.016 | 0.016 | 0.32 | 54162 |
| 8 | rs2081687 | 59388565 | T | C | 0.34 | 0.025 | 0.003 | 1.1E-22 | 314332 | 0.001 | 0.016 | 0.96 | 54162 |
| 8 | rs1149470 | 64608119 | T | A | 0.24 | 0.014 | 0.003 | 3.0E-06 | 309816 | -0.003 | 0.018 | 0.87 | 54162 |
| 8 | rs3808477 | 116670347 | C | T | 0.72 | 0.012 | 0.003 | 1.1E-05 | 313396 | -0.038 | 0.018 | 0.03 | 54162 |
| 8 | rs35859536 | 118191475 | C | T | 0.69 | 0.015 | 0.003 | 8.4E-09 | 312463 | 0.033 | 0.017 | 0.06 | 54162 |
| 8 | rs6999569 | 126475770 | A | G | 0.53 | 0.089 | 0.002 | 0.00 | 313606 | 0.018 | 0.016 | 0.26 | 54162 |
| 8 | rs2054067 | 126694377 | A | T | 0.63 | 0.012 | 0.003 | 2.9E-06 | 303457 | -0.023 | 0.016 | 0.16 | 54162 |
| 8 | rs1561928 | 129568061 | G | A | 0.89 | 0.022 | 0.004 | 2.7E-08 | 308850 | -0.026 | 0.024 | 0.28 | 54162 |
| 8 | rs72691637 | 144306970 | G | A | 0.82 | 0.016 | 0.003 | 1.3E-06 | 303763 | -0.024 | 0.025 | 0.35 | 54162 |
| 9 | rs1567353 | 1033773 | G | C | 0.31 | 0.013 | 0.003 | 8.2E-07 | 310919 | -0.013 | 0.017 | 0.43 | 54162 |
| 9 | rs7855395 | 13676484 | A | G | 0.43 | 0.014 | 0.002 | 2.5E-08 | 313148 | 0.017 | 0.016 | 0.29 | 54162 |
| 9 | rs581080 | 15305378 | C | G | 0.82 | 0.015 | 0.003 | 1.8E-06 | 314332 | 0.016 | 0.021 | 0.43 | 54162 |
| 9 | rs10811662 | 22134253 | G | A | 0.83 | 0.016 | 0.003 | 1.2E-06 | 314161 | -0.007 | 0.021 | 0.73 | 54162 |
| 9 | rs696825 | 86583076 | C | T | 0.75 | 0.022 | 0.003 | 2.1E-15 | 314174 | 0.030 | 0.018 | 0.10 | 54162 |
| 9 | rs10797119 | 92202495 | C | T | 0.54 | 0.017 | 0.002 | 1.9E-11 | 308113 | 0.008 | 0.017 | 0.62 | 54162 |
| 9 | rs2131919 | 95283887 | G | A | 0.16 | 0.016 | 0.003 | 2.7E-06 | 314087 | 0.023 | 0.021 | 0.26 | 54162 |
| 9 | rs62565259 | 102162570 | C | T | 0.83 | 0.015 | 0.003 | 3.0E-06 | 309038 | -0.017 | 0.022 | 0.45 | 54162 |

| | | | | | | | | | | | | | |
|---|---|---|---|---|---|---|---|---|---|---|---|---|---|
| 9 | rs1800978 | 107665978 | C | G | 0.88 | 0.025 | 0.004 | 3.0E-11 | 312493 | -0.058 | 0.024 | 0.01 | 54162 |
| 9 | rs77824033 | 112241136 | T | C | 0.96 | 0.022 | 0.007 | 1.1E-03 | 312755 | -0.020 | 0.040 | 0.62 | 54162 |
| 9 | rs2416759 | 123358262 | A | G | 0.70 | 0.012 | 0.003 | 1.6E-05 | 309781 | -0.008 | 0.018 | 0.65 | 54162 |
| 9 | rs4564007 | 134854280 | T | C | 0.32 | 0.012 | 0.003 | 9.5E-06 | 313579 | -0.001 | 0.017 | 0.96 | 54162 |
| 9 | rs2519093 | 136141870 | C | T | 0.81 | 0.021 | 0.003 | 5.0E-11 | 314090 | 0.003 | 0.019 | 0.86 | 54162 |
| 9 | rs4382584 | 139386138 | A | G | 0.27 | 0.016 | 0.003 | 5.6E-09 | 310625 | -0.013 | 0.018 | 0.48 | 54162 |
| 10 | rs140107293 | 5267191 | A | G | 0.84 | 0.024 | 0.003 | 8.8E-13 | 313637 | 0.008 | 0.022 | 0.71 | 54162 |
| 10 | rs3758413 | 17268839 | C | T | 0.41 | 0.010 | 0.002 | 5.1E-05 | 310206 | 0.001 | 0.016 | 0.93 | 54162 |
| 10 | rs973709 | 56639940 | G | A | 0.44 | 0.010 | 0.002 | 7.7E-05 | 312424 | 0.013 | 0.016 | 0.42 | 54162 |
| 10 | rs1171617 | 61467182 | T | G | 0.77 | 0.020 | 0.003 | 1.2E-11 | 312972 | 0.003 | 0.019 | 0.89 | 54162 |
| 10 | rs55767272 | 63702572 | A | C | 0.94 | 0.027 | 0.005 | 3.1E-08 | 313358 | 0.006 | 0.037 | 0.86 | 54162 |
| 10 | rs3829126 | 74714177 | T | G | 0.09 | 0.022 | 0.004 | 1.2E-07 | 313665 | 0.019 | 0.028 | 0.50 | 54162 |
| 10 | rs71473777 | 77217080 | G | A | 0.12 | 0.018 | 0.004 | 1.7E-06 | 312010 | -0.020 | 0.026 | 0.46 | 54162 |
| 10 | rs7077812 | 81096071 | C | T | 0.19 | 0.015 | 0.003 | 2.0E-06 | 314003 | -0.013 | 0.020 | 0.53 | 54162 |
| 10 | rs11187027 | 94259180 | A | G | 0.21 | 0.017 | 0.003 | 2.0E-08 | 313325 | -0.010 | 0.020 | 0.61 | 54162 |
| 10 | rs2068888 | 94839642 | G | A | 0.55 | 0.031 | 0.002 | 1.5E-35 | 314332 | 0.021 | 0.016 | 0.20 | 54162 |
| 10 | rs113344423 | 95316037 | A | G | 0.06 | 0.043 | 0.005 | 3.0E-15 | 307365 | 0.012 | 0.043 | 0.79 | 54162 |
| 10 | rs563296 | 99772404 | A | G | 0.56 | 0.018 | 0.002 | 1.2E-12 | 312583 | 0.000 | 0.016 | 0.99 | 54162 |
| 10 | rs75398587 | 103946480 | C | G | 0.93 | 0.025 | 0.005 | 1.8E-07 | 313129 | -0.004 | 0.033 | 0.91 | 54162 |
| 10 | rs2487294 | 113937941 | T | G | 0.73 | 0.017 | 0.003 | 8.9E-10 | 313879 | -0.012 | 0.017 | 0.47 | 54162 |
| 10 | rs12415159 | 113978850 | G | A | 0.14 | 0.020 | 0.003 | 1.6E-08 | 313512 | -0.002 | 0.022 | 0.93 | 54162 |
| 10 | rs2773469 | 115798895 | A | G | 0.26 | 0.020 | 0.003 | 4.2E-13 | 310422 | -0.005 | 0.018 | 0.77 | 54162 |
| 10 | rs2420477 | 120242187 | T | C | 0.47 | 0.012 | 0.003 | 3.6E-06 | 301691 | -0.020 | 0.016 | 0.23 | 54162 |
| 10 | rs878409 | 122999550 | G | A | 0.45 | 0.013 | 0.002 | 3.5E-07 | 312729 | -0.012 | 0.016 | 0.43 | 54162 |
| 10 | rs1133400 | 134459388 | G | A | 0.22 | 0.016 | 0.003 | 9.4E-08 | 314332 | 0.039 | 0.019 | 0.05 | 54162 |
| 11 | rs4909945 | 10673739 | C | T | 0.69 | 0.014 | 0.003 | 1.3E-07 | 314332 | 0.000 | 0.017 | 1.00 | 54162 |
| 11 | rs6486122 | 13361524 | T | C | 0.69 | 0.022 | 0.003 | 3.8E-16 | 314332 | 0.018 | 0.017 | 0.29 | 54162 |
| 11 | rs79634051 | 14561945 | G | C | 0.97 | 0.042 | 0.007 | 1.2E-08 | 314332 | -0.009 | 0.057 | 0.87 | 54162 |
| 11 | rs75268115 | 18301915 | A | G | 0.92 | 0.019 | 0.004 | 2.1E-05 | 311766 | 0.076 | 0.027 | 0.01 | 54162 |
| 11 | rs11030107 | 27694835 | G | A | 0.26 | 0.016 | 0.003 | 7.4E-09 | 314006 | 0.009 | 0.018 | 0.60 | 54162 |
| 11 | rs499293 | 30504660 | G | A | 0.34 | 0.010 | 0.003 | 1.3E-04 | 314324 | -0.004 | 0.016 | 0.81 | 54162 |
| 11 | rs77756595 | 45875161 | A | G | 0.96 | 0.038 | 0.007 | 4.6E-09 | 314332 | -0.161 | 0.049 | 1.0E-03 | 54162 |
| 11 | rs80078546 | 46195220 | C | G | 0.95 | 0.028 | 0.005 | 1.4E-07 | 314123 | 0.012 | 0.039 | 0.75 | 54162 |
| 11 | rs326222 | 47259668 | C | T | 0.70 | 0.025 | 0.003 | 2.0E-20 | 313626 | 0.009 | 0.018 | 0.61 | 54162 |
| 11 | rs140874911 | 48693639 | T | C | 0.07 | 0.032 | 0.005 | 8.7E-11 | 314016 | 0.013 | 0.037 | 0.73 | 54162 |
| 11 | rs146706984 | 49845321 | T | G | 0.08 | 0.027 | 0.005 | 1.8E-09 | 313291 | 0.058 | 0.033 | 0.08 | 54162 |
| 11 | rs10792091 | 57119156 | C | T | 0.14 | 0.018 | 0.004 | 7.2E-07 | 312899 | -0.038 | 0.023 | 0.11 | 54162 |
| 11 | rs174566 | 61592362 | G | A | 0.35 | 0.050 | 0.003 | 2.5E-84 | 313763 | -0.009 | 0.016 | 0.58 | 54162 |
| 11 | rs3017106 | 62200846 | T | C | 0.70 | 0.010 | 0.003 | 1.3E-04 | 299191 | -0.011 | 0.019 | 0.55 | 54162 |
| 11 | rs11231161 | 62378221 | G | A | 0.37 | 0.016 | 0.003 | 7.4E-10 | 314332 | -0.002 | 0.016 | 0.89 | 54162 |
| 11 | rs56271783 | 64004723 | C | G | 0.04 | 0.062 | 0.006 | 1.8E-24 | 311841 | 0.033 | 0.042 | 0.44 | 54162 |
| 11 | rs10750766 | 65473798 | A | C | 0.71 | 0.020 | 0.003 | 2.4E-13 | 312583 | 0.004 | 0.017 | 0.80 | 54162 |
| 11 | rs490972 | 66079786 | A | G | 0.47 | 0.013 | 0.002 | 2.9E-07 | 311425 | 0.016 | 0.017 | 0.36 | 54162 |
| 11 | rs11228377 | 68603346 | T | C | 0.40 | 0.016 | 0.003 | 1.1E-10 | 314332 | -0.019 | 0.016 | 0.22 | 54162 |
| 11 | rs10899490 | 78105879 | C | T | 0.84 | 0.017 | 0.003 | 3.5E-07 | 313852 | 0.046 | 0.021 | 0.03 | 54162 |
| 11 | rs2850245 | 111760738 | G | T | 0.37 | 0.016 | 0.003 | 2.6E-10 | 313421 | 0.033 | 0.016 | 0.04 | 54162 |
| 11 | rs480823 | 116525730 | C | T | 0.08 | 0.158 | 0.005 | 0.00 | 309691 | -0.013 | 0.030 | 0.67 | 54162 |
| 11 | rs61905078 | 116596309 | C | A | 0.07 | 0.206 | 0.005 | 0.00 | 313432 | 0.000 | 0.031 | 0.99 | 54162 |
| 11 | rs78484485 | 116598065 | G | A | 0.95 | 0.076 | 0.005 | 3.3E-44 | 313551 | -0.031 | 0.033 | 0.35 | 54162 |
| 11 | rs150555490 | 116697081 | C | T | 0.94 | 0.038 | 0.005 | 4.6E-13 | 314332 | -0.005 | 0.060 | 0.94 | 54162 |
| 11 | rs79357714 | 116902639 | A | G | 0.95 | 0.029 | 0.006 | 1.1E-06 | 313456 | 0.034 | 0.041 | 0.41 | 54162 |
| 11 | rs1064939 | 118396331 | A | T | 0.98 | 0.067 | 0.008 | 9.0E-16 | 313232 | 0.037 | 0.068 | 0.59 | 54162 |
| 12 | rs35104374 | 6739497 | T | C | 0.27 | 0.013 | 0.003 | 5.0E-06 | 303994 | -0.008 | 0.022 | 0.73 | 54162 |
| 12 | rs35764600 | 11791628 | C | G | 0.40 | 0.013 | 0.003 | 1.3E-07 | 303203 | 0.015 | 0.017 | 0.38 | 54162 |
| 12 | rs7134375 | 20473758 | C | A | 0.57 | 0.021 | 0.002 | 3.9E-17 | 314332 | 0.002 | 0.016 | 0.92 | 54162 |
| 12 | rs67981690 | 21343886 | G | A | 0.13 | 0.033 | 0.004 | 1.0E-18 | 310685 | 0.021 | 0.023 | 0.36 | 54162 |
| 12 | rs9943778 | 22765864 | A | G | 0.76 | 0.012 | 0.003 | 6.7E-05 | 302158 | -0.014 | 0.019 | 0.47 | 54162 |
| 12 | rs10842703 | 26456188 | T | A | 0.24 | 0.013 | 0.003 | 6.9E-06 | 314307 | 0.001 | 0.018 | 0.96 | 54162 |
| 12 | rs7136223 | 29437861 | A | G | 0.73 | 0.013 | 0.003 | 1.6E-06 | 311186 | -0.028 | 0.027 | 0.30 | 54162 |
| 12 | rs4002684 | 37896288 | A | T | 0.59 | 0.011 | 0.003 | 2.3E-05 | 308695 | -0.007 | 0.022 | 0.74 | 54162 |
| 12 | rs35763453 | 46215895 | C | T | 0.05 | 0.029 | 0.005 | 1.2E-07 | 308141 | -0.010 | 0.039 | 0.79 | 54162 |

| | | | | | | | | | | | | | |
|---|---|---|---|---|---|---|---|---|---|---|---|---|---|
| 12 | rs12422600 | 54429385 | G | A | 0.63 | 0.011 | 0.003 | 3.9E-05 | 308959 | -0.008 | 0.017 | 0.62 | 54162 |
| 12 | rs4760254 | 57766392 | G | C | 0.76 | 0.028 | 0.003 | 5.3E-22 | 314140 | -0.010 | 0.019 | 0.58 | 54162 |
| 12 | rs113439801 | 62838230 | C | T | 0.83 | 0.021 | 0.003 | 3.9E-10 | 310101 | 0.002 | 0.021 | 0.91 | 54162 |
| 12 | rs1351394 | 66351826 | C | T | 0.51 | 0.013 | 0.002 | 7.9E-08 | 314332 | 0.025 | 0.016 | 0.12 | 54162 |
| 12 | rs775633 | 67645247 | T | A | 0.34 | 0.014 | 0.003 | 8.0E-08 | 308804 | -0.008 | 0.017 | 0.64 | 54162 |
| 12 | rs4761234 | 69732105 | T | C | 0.51 | 0.017 | 0.002 | 8.1E-12 | 311332 | 0.009 | 0.016 | 0.56 | 54162 |
| 12 | rs7138037 | 107228676 | C | G | 0.23 | 0.019 | 0.003 | 3.7E-11 | 314134 | 0.038 | 0.019 | 0.04 | 54162 |
| 12 | rs9788220 | 109699616 | C | T | 0.82 | 0.015 | 0.003 | 3.7E-06 | 313788 | -0.003 | 0.020 | 0.89 | 54162 |
| 12 | rs580063 | 123206340 | T | C | 0.79 | 0.024 | 0.003 | 1.1E-15 | 313462 | 0.042 | 0.020 | 0.03 | 54162 |
| 12 | rs4930724 | 124423817 | T | C | 0.67 | 0.026 | 0.003 | 7.5E-23 | 313637 | -0.010 | 0.017 | 0.58 | 54162 |
| 12 | rs863750 | 124505444 | T | C | 0.60 | 0.029 | 0.003 | 2.4E-30 | 311191 | -0.018 | 0.016 | 0.28 | 54162 |
| 12 | rs112403212 | 125303254 | T | C | 0.14 | 0.022 | 0.004 | 3.7E-10 | 309664 | -0.040 | 0.025 | 0.11 | 54162 |
| 13 | rs1340819 | 29145323 | A | C | 0.65 | 0.014 | 0.003 | 1.6E-07 | 313845 | 0.013 | 0.017 | 0.44 | 54162 |
| 13 | rs2812208 | 50707087 | G | C | 0.98 | 0.054 | 0.008 | 1.9E-10 | 314214 | 0.020 | 0.051 | 0.69 | 54162 |
| 13 | rs9561643 | 95253131 | C | A | 0.31 | 0.016 | 0.003 | 2.3E-09 | 313234 | 0.017 | 0.017 | 0.32 | 54162 |
| 13 | rs9584870 | 99245866 | T | C | 0.64 | 0.012 | 0.003 | 3.8E-06 | 296048 | -0.002 | 0.018 | 0.93 | 54162 |
| 13 | rs1556124 | 110991189 | A | G | 0.78 | 0.014 | 0.003 | 2.4E-06 | 314332 | 0.039 | 0.019 | 0.04 | 54162 |
| 13 | rs2774430 | 112245313 | G | A | 0.56 | 0.011 | 0.003 | 9.2E-06 | 300634 | -0.011 | 0.016 | 0.52 | 54162 |
| 13 | rs7400002 | 114524944 | G | A | 0.23 | 0.012 | 0.003 | 6.3E-05 | 312173 | -0.062 | 0.026 | 0.02 | 54162 |
| 13 | rs7140110 | 114544024 | C | T | 0.30 | 0.030 | 0.003 | 2.0E-28 | 313143 | -0.014 | 0.019 | 0.46 | 54162 |
| 13 | rs79192570 | 114625570 | G | A | 0.86 | 0.026 | 0.003 | 6.5E-14 | 313185 | 0.019 | 0.022 | 0.40 | 54162 |
| 14 | rs56902258 | 23733114 | T | A | 0.80 | 0.014 | 0.003 | 1.2E-05 | 310297 | 0.024 | 0.020 | 0.24 | 54162 |
| 14 | rs6572807 | 52480621 | G | A | 0.27 | 0.012 | 0.003 | 1.3E-05 | 313980 | 0.001 | 0.018 | 0.96 | 54162 |
| 14 | rs61975915 | 58665283 | C | T | 0.71 | 0.012 | 0.003 | 1.5E-05 | 303895 | 0.031 | 0.020 | 0.13 | 54162 |
| 14 | rs12880341 | 64236191 | C | T | 0.16 | 0.020 | 0.003 | 1.9E-09 | 311899 | 0.002 | 0.064 | 0.97 | 54162 |
| 14 | rs2240533 | 71541026 | T | C | 0.69 | 0.013 | 0.003 | 9.3E-07 | 312426 | 0.008 | 0.017 | 0.66 | 54162 |
| 14 | rs61993685 | 100765823 | T | C | 0.92 | 0.022 | 0.005 | 1.4E-06 | 314332 | -0.048 | 0.054 | 0.37 | 54162 |
| 15 | rs28624578 | 31637666 | C | T | 0.17 | 0.012 | 0.003 | 1.8E-04 | 311907 | -0.047 | 0.023 | 0.04 | 54162 |
| 15 | rs34245505 | 40397191 | G | C | 0.19 | 0.019 | 0.003 | 2.5E-09 | 306067 | 0.024 | 0.025 | 0.34 | 54162 |
| 15 | rs28364531 | 42701374 | T | C | 0.02 | 0.086 | 0.009 | 2.5E-20 | 313993 | -0.030 | 0.059 | 0.60 | 54162 |
| 15 | rs139974673 | 44027885 | C | T | 0.02 | 0.147 | 0.008 | 4.0E-77 | 314058 | -0.025 | 0.046 | 0.59 | 54162 |
| 15 | rs138751626 | 57154952 | T | A | 0.08 | 0.025 | 0.005 | 4.3E-08 | 313530 | -0.018 | 0.031 | 0.56 | 54162 |
| 15 | rs1532085 | 58683366 | A | G | 0.39 | 0.033 | 0.003 | 2.7E-40 | 314332 | -0.039 | 0.016 | 0.01 | 54162 |
| 15 | rs1077835 | 58723426 | G | A | 0.22 | 0.045 | 0.003 | 8.4E-53 | 312856 | 0.001 | 0.019 | 0.96 | 54162 |
| 15 | rs12591786 | 60902512 | C | T | 0.85 | 0.018 | 0.003 | 2.1E-07 | 304635 | 0.035 | 0.023 | 0.13 | 54162 |
| 15 | rs12440800 | 61960302 | T | A | 0.25 | 0.015 | 0.003 | 8.1E-08 | 311258 | -0.012 | 0.018 | 0.50 | 54162 |
| 15 | rs2652806 | 63374127 | C | T | 0.33 | 0.016 | 0.003 | 2.5E-09 | 305709 | -0.049 | 0.017 | 2.8E-03 | 54162 |
| 15 | rs11635675 | 63793238 | G | T | 0.34 | 0.024 | 0.003 | 2.5E-20 | 312853 | 0.029 | 0.017 | 0.08 | 54162 |
| 15 | rs4776793 | 66872114 | T | C | 0.36 | 0.017 | 0.003 | 4.6E-11 | 314309 | 0.000 | 0.016 | 1.00 | 54162 |
| 15 | rs3784310 | 72103427 | T | C | 0.72 | 0.013 | 0.003 | 1.5E-06 | 309056 | 0.017 | 0.019 | 0.36 | 54162 |
| 15 | rs3826043 | 73618238 | C | T | 0.57 | 0.012 | 0.003 | 2.1E-06 | 305445 | -0.011 | 0.016 | 0.51 | 54162 |
| 15 | rs2017500 | 99196112 | A | G | 0.51 | 0.014 | 0.002 | 1.5E-08 | 309265 | -0.001 | 0.016 | 0.95 | 54162 |
| 15 | rs10152471 | 101890913 | G | A | 0.61 | 0.012 | 0.003 | 9.7E-07 | 312230 | 0.031 | 0.018 | 0.08 | 54162 |
| 15 | rs8025505 | 102067841 | T | C | 0.25 | 0.023 | 0.003 | 1.1E-15 | 311067 | 0.030 | 0.021 | 0.16 | 54162 |
| 16 | rs12600110 | 962154 | T | C | 0.62 | 0.013 | 0.003 | 5.2E-07 | 313217 | -0.019 | 0.016 | 0.26 | 54162 |
| 16 | rs28577186 | 4488191 | G | A | 0.33 | 0.014 | 0.003 | 3.2E-08 | 310223 | 0.009 | 0.018 | 0.61 | 54162 |
| 16 | rs933574 | 11792700 | C | A | 0.48 | 0.015 | 0.002 | 1.1E-09 | 312172 | -0.003 | 0.015 | 0.84 | 54162 |
| 16 | rs12928099 | 15150505 | C | A | 0.70 | 0.030 | 0.003 | 1.0E-29 | 313390 | 0.000 | 0.018 | 0.98 | 54162 |
| 16 | rs3814883 | 29994922 | T | C | 0.48 | 0.016 | 0.002 | 1.2E-10 | 310086 | -0.029 | 0.016 | 0.07 | 54162 |
| 16 | rs2288004 | 31054040 | G | C | 0.62 | 0.015 | 0.003 | 1.4E-09 | 314332 | -0.038 | 0.016 | 0.02 | 54162 |
| 16 | rs12446515 | 56987015 | C | T | 0.68 | 0.036 | 0.003 | 6.8E-42 | 310807 | -0.001 | 0.017 | 0.97 | 54162 |
| 16 | rs62064941 | 58834402 | A | C | 0.96 | 0.027 | 0.006 | 1.7E-05 | 314207 | -0.010 | 0.041 | 0.81 | 54162 |
| 16 | rs7186635 | 69378445 | G | A | 0.32 | 0.010 | 0.003 | 1.3E-04 | 313746 | 0.018 | 0.017 | 0.30 | 54162 |
| 16 | rs2917677 | 69750849 | C | T | 0.59 | 0.019 | 0.002 | 3.4E-14 | 312348 | -0.004 | 0.016 | 0.82 | 54162 |
| 16 | rs3794695 | 72097827 | T | C | 0.19 | 0.029 | 0.003 | 5.3E-20 | 312772 | -0.009 | 0.020 | 0.63 | 54162 |
| 16 | rs7191623 | 72212044 | G | A | 0.79 | 0.015 | 0.003 | 1.5E-06 | 312046 | 0.012 | 0.019 | 0.51 | 54162 |
| 16 | rs2925979 | 81534790 | T | C | 0.30 | 0.031 | 0.003 | 7.2E-32 | 314332 | 0.006 | 0.018 | 0.72 | 54162 |
| 16 | rs7199293 | 81614892 | A | G | 0.53 | 0.010 | 0.002 | 3.5E-05 | 306249 | 0.010 | 0.016 | 0.54 | 54162 |
| 16 | rs79311290 | 85150163 | G | A | 0.09 | 0.026 | 0.004 | 4.1E-09 | 289305 | -0.006 | 0.046 | 0.90 | 54162 |
| 16 | rs1728407 | 86422112 | G | A | 0.45 | 0.012 | 0.002 | 1.0E-06 | 312427 | -0.027 | 0.016 | 0.10 | 54162 |
| 16 | rs12926107 | 88004092 | G | A | 0.46 | 0.012 | 0.002 | 7.5E-07 | 312585 | -0.001 | 0.017 | 0.95 | 54162 |

| | | | | | | | | | | | | | |
|---|---|---|---|---|---|---|---|---|---|---|---|---|---|
| 17 | rs11657201 | 599924 | G | A | 0.24 | 0.012 | 0.003 | 3.8E-05 | 312832 | -0.008 | 0.019 | 0.67 | 54162 |
| 17 | rs11078597 | 1618363 | C | T | 0.19 | 0.021 | 0.003 | 3.0E-11 | 314332 | 0.056 | 0.024 | 0.02 | 54162 |
| 17 | rs7215055 | 17458353 | G | A | 0.06 | 0.041 | 0.005 | 1.1E-15 | 313816 | 0.020 | 0.031 | 0.52 | 54162 |
| 17 | rs704 | 26694861 | G | A | 0.53 | 0.013 | 0.002 | 1.7E-07 | 314332 | 0.006 | 0.017 | 0.73 | 54162 |
| 17 | rs591939 | 40698075 | G | A | 0.25 | 0.021 | 0.003 | 3.5E-14 | 313394 | 0.023 | 0.019 | 0.22 | 54162 |
| 17 | rs116878033 | 42031770 | T | C | 0.04 | 0.044 | 0.007 | 3.1E-11 | 311912 | 0.009 | 0.084 | 0.91 | 54162 |
| 17 | rs112162280 | 43194413 | C | T | 0.71 | 0.013 | 0.003 | 9.2E-07 | 308162 | -0.013 | 0.017 | 0.47 | 54162 |
| 17 | rs10775406 | 46197755 | G | A | 0.76 | 0.021 | 0.003 | 2.6E-13 | 314324 | -0.014 | 0.018 | 0.46 | 54162 |
| 17 | rs595767 | 46957987 | G | A | 0.52 | 0.013 | 0.002 | 4.6E-08 | 311673 | 0.005 | 0.016 | 0.77 | 54162 |
| 17 | rs12185242 | 47407071 | C | A | 0.46 | 0.015 | 0.002 | 9.5E-10 | 313100 | 0.058 | 0.016 | 2.0E-04 | 54162 |
| 17 | rs9890200 | 48624523 | A | C | 0.63 | 0.014 | 0.003 | 6.7E-08 | 312960 | -0.012 | 0.017 | 0.47 | 54162 |
| 17 | rs1292065 | 57906288 | C | G | 0.29 | 0.016 | 0.003 | 4.6E-09 | 314303 | -0.007 | 0.017 | 0.68 | 54162 |
| 17 | rs1801689 | 64210580 | A | C | 0.97 | 0.065 | 0.007 | 3.1E-19 | 314332 | 0.144 | 0.076 | 0.06 | 54162 |
| 17 | rs62084237 | 65854807 | A | G | 0.17 | 0.026 | 0.003 | 2.6E-15 | 304374 | -0.007 | 0.022 | 0.75 | 54162 |
| 17 | rs8066985 | 68453345 | A | G | 0.48 | 0.013 | 0.002 | 1.2E-07 | 313071 | 0.001 | 0.016 | 0.94 | 54162 |
| 17 | rs4559942 | 73309269 | A | G | 0.78 | 0.015 | 0.003 | 4.1E-07 | 310620 | 0.008 | 0.018 | 0.64 | 54162 |
| 17 | rs77244849 | 74281391 | T | C | 0.69 | 0.016 | 0.003 | 9.3E-10 | 308931 | 0.004 | 0.018 | 0.82 | 54162 |
| 17 | rs4969179 | 76391454 | T | G | 0.40 | 0.018 | 0.003 | 2.3E-12 | 313904 | -0.019 | 0.016 | 0.23 | 54162 |
| 18 | rs6506033 | 289209 | C | T | 0.93 | 0.027 | 0.005 | 1.7E-08 | 314071 | -0.003 | 0.030 | 0.92 | 54162 |
| 18 | rs11664106 | 2846812 | A | T | 0.63 | 0.013 | 0.003 | 1.2E-06 | 288602 | 0.031 | 0.019 | 0.09 | 54162 |
| 18 | rs867939 | 19911690 | G | A | 0.42 | 0.014 | 0.002 | 3.6E-08 | 310456 | 0.013 | 0.016 | 0.42 | 54162 |
| 18 | rs7239575 | 21120035 | T | C | 0.51 | 0.017 | 0.002 | 2.7E-12 | 314171 | -0.012 | 0.016 | 0.45 | 54162 |
| 18 | rs41292412 | 56118358 | T | C | 0.01 | 0.058 | 0.011 | 3.9E-07 | 313337 | 0.092 | 0.087 | 0.29 | 54162 |
| 18 | rs921971 | 57861663 | C | T | 0.27 | 0.016 | 0.003 | 4.9E-09 | 312879 | -0.038 | 0.018 | 0.04 | 54162 |
| 18 | rs12454712 | 60845884 | T | C | 0.62 | 0.012 | 0.003 | 1.1E-06 | 314332 | 0.039 | 0.018 | 0.03 | 54162 |
| 18 | rs2187114 | 70517838 | G | A | 0.90 | 0.019 | 0.004 | 2.4E-06 | 314332 | -0.018 | 0.028 | 0.52 | 54162 |
| 19 | rs7260465 | 4139440 | C | T | 0.74 | 0.013 | 0.003 | 2.0E-06 | 308996 | 0.031 | 0.019 | 0.10 | 54162 |
| 19 | rs2860183 | 7189375 | T | C | 0.38 | 0.013 | 0.003 | 4.4E-07 | 309456 | 0.011 | 0.017 | 0.52 | 54162 |
| 19 | rs3890483 | 7220596 | T | G | 0.44 | 0.021 | 0.002 | 1.9E-17 | 310345 | 0.019 | 0.016 | 0.24 | 54162 |
| 19 | rs62117489 | 8442202 | C | A | 0.95 | 0.047 | 0.005 | 3.4E-18 | 313670 | 0.037 | 0.038 | 0.32 | 54162 |
| 19 | rs2278426 | 11350488 | C | T | 0.97 | 0.039 | 0.007 | 8.8E-09 | 314332 | 0.009 | 0.043 | 0.83 | 54162 |
| 19 | rs58542926 | 19379549 | C | T | 0.92 | 0.105 | 0.005 | 0.00 | 314332 | -0.015 | 0.031 | 0.62 | 54162 |
| 19 | rs55737395 | 33751349 | G | A | 0.66 | 0.013 | 0.003 | 1.5E-06 | 307581 | -0.014 | 0.018 | 0.45 | 54162 |
| 19 | rs10422861 | 33894846 | C | T | 0.33 | 0.020 | 0.003 | 1.6E-14 | 313424 | 0.014 | 0.017 | 0.39 | 54162 |
| 19 | rs58895965 | 35551428 | A | C | 0.18 | 0.024 | 0.003 | 3.9E-14 | 314183 | 0.004 | 0.021 | 0.86 | 54162 |
| 19 | rs4802113 | 41740895 | T | C | 0.54 | 0.014 | 0.002 | 5.5E-09 | 309184 | 0.017 | 0.016 | 0.31 | 54162 |
| 19 | rs296360 | 48388658 | T | C | 0.84 | 0.017 | 0.003 | 4.5E-07 | 311399 | -0.015 | 0.021 | 0.48 | 54162 |
| 19 | rs838133 | 49259529 | A | G | 0.45 | 0.023 | 0.003 | 1.8E-18 | 277625 | 0.026 | 0.017 | 0.12 | 54162 |
| 19 | rs12610709 | 56102362 | A | G | 0.17 | 0.024 | 0.003 | 1.2E-13 | 312279 | -0.034 | 0.026 | 0.19 | 54162 |
| 19 | rs8102873 | 57488423 | T | C | 0.58 | 0.012 | 0.002 | 1.3E-06 | 314332 | -0.015 | 0.017 | 0.39 | 54162 |
| 20 | rs151235402 | 569164 | T | C | 0.01 | 0.050 | 0.010 | 6.9E-07 | 312740 | 0.054 | 0.087 | 0.54 | 54162 |
| 20 | rs293561 | 31091206 | G | T | 0.36 | 0.009 | 0.003 | 5.1E-04 | 307459 | -0.025 | 0.017 | 0.14 | 54162 |
| 20 | rs149142833 | 32188142 | T | C | 0.15 | 0.018 | 0.003 | 1.4E-07 | 310311 | 0.026 | 0.024 | 0.29 | 54162 |
| 20 | rs55837381 | 38556466 | G | A | 0.75 | 0.016 | 0.003 | 1.0E-08 | 309931 | -0.008 | 0.018 | 0.65 | 54162 |
| 20 | rs6093446 | 39780932 | A | G | 0.29 | 0.015 | 0.003 | 1.9E-08 | 313980 | 0.006 | 0.017 | 0.73 | 54162 |
| 20 | rs6073958 | 44551855 | C | T | 0.20 | 0.056 | 0.003 | 1.4E-74 | 313901 | 0.027 | 0.021 | 0.19 | 54162 |
| 20 | rs55966194 | 45599090 | C | G | 0.72 | 0.021 | 0.003 | 3.5E-14 | 313795 | 0.028 | 0.018 | 0.12 | 54162 |
| 20 | rs6068280 | 51235613 | G | A | 0.68 | 0.013 | 0.003 | 8.5E-07 | 309851 | 0.011 | 0.017 | 0.50 | 54162 |
| 20 | rs7274718 | 56113783 | A | G | 0.60 | 0.017 | 0.002 | 3.2E-11 | 314124 | 0.010 | 0.016 | 0.52 | 54162 |
| 20 | rs8126001 | 62711459 | C | T | 0.51 | 0.017 | 0.002 | 4.5E-12 | 310788 | -0.009 | 0.019 | 0.65 | 54162 |
| 21 | rs6517522 | 40553845 | T | C | 0.50 | 0.011 | 0.002 | 1.5E-05 | 313496 | 0.007 | 0.016 | 0.68 | 54162 |
| 21 | rs394872 | 46582100 | T | C | 0.54 | 0.011 | 0.002 | 1.9E-05 | 311397 | -0.012 | 0.017 | 0.46 | 54162 |
| 22 | rs140288 | 24335977 | G | A | 0.43 | 0.011 | 0.002 | 1.7E-05 | 313741 | 0.010 | 0.020 | 0.62 | 54162 |
| 22 | rs134551 | 28807625 | C | T | 0.66 | 0.009 | 0.003 | 1.1E-03 | 313028 | -0.021 | 0.017 | 0.21 | 54162 |
| 22 | rs9610329 | 36042986 | T | C | 0.43 | 0.014 | 0.003 | 1.5E-07 | 290890 | 0.006 | 0.018 | 0.73 | 54162 |
| 22 | rs6000553 | 37469192 | G | A | 0.53 | 0.012 | 0.002 | 1.8E-06 | 310467 | -0.006 | 0.016 | 0.70 | 54162 |
| 22 | rs2267373 | 38600542 | T | C | 0.58 | 0.022 | 0.002 | 3.7E-19 | 313801 | 0.011 | 0.017 | 0.49 | 54162 |
| 22 | rs2071887 | 38879010 | A | T | 0.34 | 0.017 | 0.003 | 1.5E-11 | 313255 | -0.028 | 0.016 | 0.08 | 54162 |
| 22 | rs60610697 | 46619870 | G | T | 0.21 | 0.014 | 0.003 | 3.6E-06 | 303409 | -0.004 | 0.020 | 0.86 | 54162 |

**Supplementary table 5:** List of 338 SNPs used as instrumental variables for ApoA, and their association with ApoA and dementia.

| CHR | rsID | POS | Effect allele | Other allele | EAF | BETA apoA | SE apoA | Pval apoA | n apoA | BETA dementia | SE dementia | Pval dementia | n dementia |
|---|---|---|---|---|---|---|---|---|---|---|---|---|---|
| 1 | rs589942 | 20916080 | C | G | 0.66 | 0.017 | 0.003 | 6.1E-11 | 291727 | -0.002 | 0.016 | 0.91 | 54162 |
| 1 | rs4648892 | 23720185 | T | C | 0.72 | 0.016 | 0.003 | 7.2E-10 | 292854 | 0.006 | 0.018 | 0.73 | 54162 |
| 1 | rs6686889 | 25030470 | C | T | 0.75 | 0.015 | 0.003 | 2.6E-08 | 292222 | -0.002 | 0.018 | 0.91 | 54162 |
| 1 | rs34397747 | 27092322 | T | C | 0.92 | 0.042 | 0.004 | 4.4E-22 | 289867 | -0.032 | 0.029 | 0.26 | 54162 |
| 1 | rs4654395 | 29567412 | C | T | 0.47 | 0.010 | 0.002 | 2.8E-05 | 288028 | -0.013 | 0.016 | 0.42 | 54162 |
| 1 | rs3768321 | 40035928 | G | T | 0.80 | 0.041 | 0.003 | 2.2E-41 | 291941 | -0.023 | 0.021 | 0.26 | 54162 |
| 1 | rs3014246 | 46086077 | T | C | 0.70 | 0.014 | 0.003 | 7.2E-08 | 292854 | 0.002 | 0.017 | 0.92 | 54162 |
| 1 | rs12074528 | 63010871 | C | T | 0.65 | 0.042 | 0.003 | 6.3E-62 | 292603 | -0.033 | 0.016 | 0.04 | 54162 |
| 1 | rs7546242 | 66093030 | T | C | 0.37 | 0.017 | 0.002 | 1.3E-11 | 292342 | 0.018 | 0.016 | 0.25 | 54162 |
| 1 | rs698927 | 93836218 | C | A | 0.18 | 0.025 | 0.003 | 3.4E-16 | 292557 | -0.030 | 0.021 | 0.14 | 54162 |
| 1 | rs12740374 | 109817590 | T | G | 0.22 | 0.046 | 0.003 | 5.6E-58 | 292854 | 0.000 | 0.019 | 0.98 | 54162 |
| 1 | rs12045101 | 110267651 | C | T | 0.76 | 0.018 | 0.003 | 1.4E-10 | 290613 | 0.006 | 0.019 | 0.73 | 54162 |
| 1 | rs267738 | 150940625 | G | T | 0.22 | 0.035 | 0.003 | 1.9E-33 | 292854 | -0.033 | 0.020 | 0.09 | 54162 |
| 1 | rs9426827 | 154589965 | C | T | 0.48 | 0.017 | 0.002 | 3.6E-13 | 292794 | 0.003 | 0.016 | 0.86 | 54162 |
| 1 | rs4652192 | 176748136 | A | C | 0.19 | 0.016 | 0.003 | 2.4E-07 | 287164 | 0.012 | 0.020 | 0.57 | 54162 |
| 1 | rs10798615 | 178513895 | T | G | 0.47 | 0.018 | 0.002 | 1.6E-13 | 292572 | -0.007 | 0.015 | 0.65 | 54162 |
| 1 | rs3747973 | 205677148 | G | A | 0.59 | 0.014 | 0.002 | 1.7E-08 | 289692 | -0.011 | 0.016 | 0.49 | 54162 |
| 1 | rs2642438 | 220970028 | G | A | 0.70 | 0.030 | 0.003 | 3.3E-31 | 292854 | 0.006 | 0.019 | 0.77 | 54162 |
| 1 | rs2281718 | 230297778 | T | A | 0.61 | 0.056 | 0.002 | 2.9E-113 | 291779 | 0.011 | 0.017 | 0.54 | 54162 |
| 1 | rs1043897 | 230416399 | T | G | 0.41 | 0.019 | 0.002 | 1.3E-14 | 289220 | -0.019 | 0.016 | 0.24 | 54162 |
| 1 | rs557933 | 234853268 | C | A | 0.52 | 0.022 | 0.002 | 1.0E-19 | 289851 | 0.020 | 0.016 | 0.20 | 54162 |
| 1 | rs16844296 | 235108308 | G | A | 0.79 | 0.015 | 0.003 | 2.2E-07 | 291182 | -0.002 | 0.025 | 0.93 | 54162 |
| 2 | rs1107850 | 20371772 | G | A | 0.53 | 0.025 | 0.002 | 8.8E-25 | 291316 | 0.020 | 0.016 | 0.21 | 54162 |
| 2 | rs74333499 | 20499704 | T | C | 0.95 | 0.020 | 0.005 | 2.3E-04 | 283583 | -0.059 | 0.050 | 0.24 | 54162 |
| 2 | rs676210 | 21231524 | A | G | 0.20 | 0.065 | 0.003 | 1.9E-104 | 292854 | -0.052 | 0.019 | 0.01 | 54162 |
| 2 | rs1260326 | 27730940 | T | C | 0.39 | 0.028 | 0.002 | 1.1E-29 | 292854 | -0.001 | 0.016 | 0.96 | 54162 |
| 2 | rs17321999 | 30479857 | C | A | 0.79 | 0.015 | 0.003 | 5.2E-07 | 292854 | -0.033 | 0.020 | 0.10 | 54162 |
| 2 | rs72796869 | 32381964 | T | C | 0.04 | 0.030 | 0.006 | 2.2E-06 | 290065 | -0.009 | 0.045 | 0.84 | 54162 |
| 2 | rs848638 | 36808350 | C | T | 0.64 | 0.011 | 0.003 | 6.8E-06 | 291563 | -0.003 | 0.017 | 0.88 | 54162 |
| 2 | rs62135193 | 48755477 | C | T | 0.46 | 0.012 | 0.002 | 8.4E-07 | 287973 | 0.003 | 0.017 | 0.84 | 54162 |
| 2 | rs17326656 | 48962291 | G | T | 0.76 | 0.014 | 0.003 | 8.8E-07 | 289697 | -0.004 | 0.019 | 0.85 | 54162 |
| 2 | rs2540951 | 65276736 | G | A | 0.37 | 0.010 | 0.002 | 2.3E-05 | 292442 | -0.016 | 0.016 | 0.32 | 54162 |
| 2 | rs11883967 | 66673862 | C | A | 0.66 | 0.012 | 0.003 | 3.4E-06 | 287905 | 0.019 | 0.016 | 0.24 | 54162 |
| 2 | rs4599108 | 85543222 | T | C | 0.48 | 0.014 | 0.002 | 2.2E-08 | 277195 | -0.004 | 0.016 | 0.79 | 54162 |
| 2 | rs11691486 | 100796182 | C | T | 0.24 | 0.015 | 0.003 | 7.4E-08 | 288496 | -0.033 | 0.019 | 0.08 | 54162 |
| 2 | rs6430954 | 128592495 | G | T | 0.68 | 0.014 | 0.003 | 1.6E-07 | 292580 | 0.000 | 0.017 | 0.98 | 54162 |
| 2 | rs1446585 | 136407479 | G | A | 0.23 | 0.022 | 0.003 | 7.4E-15 | 292854 | 0.028 | 0.018 | 0.11 | 54162 |
| 2 | rs75265117 | 165518799 | G | C | 0.12 | 0.031 | 0.004 | 1.5E-16 | 292406 | 0.003 | 0.025 | 0.91 | 54162 |
| 2 | rs1862069 | 169933741 | A | G | 0.54 | 0.012 | 0.002 | 5.0E-07 | 292854 | 0.001 | 0.016 | 0.93 | 54162 |
| 2 | rs11690597 | 174085741 | G | A | 0.21 | 0.016 | 0.003 | 1.5E-07 | 289471 | 0.004 | 0.019 | 0.84 | 54162 |
| 2 | rs72926946 | 203477868 | C | A | 0.70 | 0.013 | 0.003 | 6.1E-07 | 292137 | 0.029 | 0.017 | 0.09 | 54162 |
| 2 | rs1047891 | 211540507 | C | A | 0.68 | 0.024 | 0.003 | 7.6E-21 | 292854 | 0.015 | 0.020 | 0.43 | 54162 |
| 2 | rs2943645 | 227099180 | C | T | 0.35 | 0.034 | 0.003 | 3.9E-42 | 292854 | -0.011 | 0.016 | 0.52 | 54162 |
| 2 | rs13411625 | 238690424 | A | T | 0.79 | 0.014 | 0.003 | 1.5E-06 | 291818 | -0.045 | 0.019 | 0.02 | 54162 |
| 2 | rs59104589 | 242237902 | T | C | 0.36 | 0.018 | 0.002 | 1.2E-12 | 292655 | 0.001 | 0.017 | 0.98 | 54162 |
| 3 | rs2972166 | 12316339 | A | G | 0.27 | 0.017 | 0.003 | 7.3E-10 | 288351 | -0.026 | 0.019 | 0.17 | 54162 |
| 3 | rs13097947 | 15846011 | C | T | 0.65 | 0.015 | 0.003 | 4.9E-09 | 270517 | -0.004 | 0.017 | 0.79 | 54162 |
| 3 | rs9866679 | 25516821 | T | G | 0.76 | 0.014 | 0.003 | 6.9E-07 | 291770 | -0.027 | 0.020 | 0.17 | 54162 |
| 3 | rs4441609 | 36892717 | C | T | 0.63 | 0.012 | 0.002 | 1.9E-06 | 291640 | 0.002 | 0.017 | 0.92 | 54162 |
| 3 | rs4676609 | 39214256 | T | C | 0.20 | 0.013 | 0.003 | 1.3E-05 | 290530 | -0.042 | 0.020 | 0.04 | 54162 |
| 3 | rs6765484 | 50041313 | T | C | 0.47 | 0.022 | 0.002 | 7.9E-20 | 291390 | -0.009 | 0.016 | 0.56 | 54162 |
| 3 | rs73082723 | 51926817 | A | C | 0.21 | 0.018 | 0.003 | 4.8E-09 | 266126 | -0.008 | 0.035 | 0.82 | 54162 |
| 3 | rs13326165 | 52532118 | A | G | 0.20 | 0.020 | 0.003 | 2.6E-11 | 292854 | 0.031 | 0.020 | 0.11 | 54162 |
| 3 | rs1852922 | 70936712 | A | G | 0.69 | 0.013 | 0.003 | 3.3E-07 | 287943 | 0.004 | 0.017 | 0.82 | 54162 |
| 3 | rs17008972 | 71756272 | A | G | 0.12 | 0.017 | 0.004 | 4.1E-06 | 291482 | 0.048 | 0.024 | 0.05 | 54162 |
| 3 | rs34184910 | 104274356 | G | A | 0.37 | 0.012 | 0.002 | 3.0E-06 | 292133 | -0.026 | 0.016 | 0.11 | 54162 |
| 3 | rs73216700 | 108865198 | A | G | 0.43 | 0.017 | 0.002 | 8.3E-12 | 292284 | -0.004 | 0.016 | 0.81 | 54162 |
| 3 | rs3732356 | 119529113 | G | T | 0.06 | 0.038 | 0.005 | 1.4E-14 | 290385 | 0.031 | 0.039 | 0.43 | 54162 |

| | | | | | | | | | | | | | |
|---|---|---|---|---|---|---|---|---|---|---|---|---|---|
| 3 | rs34642857 | 123051019 | T | C | 0.75 | 0.010 | 0.003 | 1.7E-04 | 286914 | 0.040 | 0.019 | 0.04 | 54162 |
| 3 | rs61789561 | 135917627 | A | G | 0.20 | 0.029 | 0.003 | 2.4E-22 | 292490 | 0.026 | 0.020 | 0.19 | 54162 |
| 3 | rs6807935 | 141114293 | A | G | 0.65 | 0.013 | 0.003 | 1.8E-07 | 292854 | 0.011 | 0.016 | 0.52 | 54162 |
| 3 | rs62271373 | 150066540 | T | A | 0.94 | 0.025 | 0.005 | 1.6E-06 | 287533 | 0.037 | 0.035 | 0.30 | 54162 |
| 3 | rs35650976 | 151990779 | T | C | 0.30 | 0.014 | 0.003 | 1.5E-07 | 283553 | -0.057 | 0.018 | 1.4E-03 | 54162 |
| 3 | rs1086056 | 154088411 | T | G | 0.16 | 0.018 | 0.003 | 2.6E-08 | 291831 | -0.025 | 0.023 | 0.26 | 54162 |
| 3 | rs9817452 | 156795414 | T | G | 0.39 | 0.018 | 0.002 | 7.9E-14 | 290949 | 0.016 | 0.017 | 0.33 | 54162 |
| 3 | rs6785881 | 160153989 | C | T | 0.52 | 0.012 | 0.002 | 9.3E-07 | 292341 | -0.025 | 0.016 | 0.11 | 54162 |
| 3 | rs73052033 | 185828465 | T | C | 0.82 | 0.018 | 0.003 | 1.1E-08 | 290727 | 0.030 | 0.021 | 0.15 | 54162 |
| 3 | rs1400362 | 185883562 | C | T | 0.75 | 0.010 | 0.003 | 6.9E-04 | 282064 | 0.000 | 0.018 | 0.99 | 54162 |
| 3 | rs78359342 | 196160928 | G | T | 0.73 | 0.012 | 0.003 | 1.7E-05 | 287138 | 0.006 | 0.021 | 0.76 | 54162 |
| 4 | rs13108218 | 3443931 | A | G | 0.38 | 0.013 | 0.003 | 2.4E-07 | 283790 | -0.015 | 0.018 | 0.39 | 54162 |
| 4 | rs1395221 | 24626903 | G | T | 0.60 | 0.013 | 0.002 | 3.2E-07 | 287415 | 0.004 | 0.016 | 0.79 | 54162 |
| 4 | rs6448429 | 26066863 | C | T | 0.84 | 0.023 | 0.003 | 8.7E-13 | 287787 | 0.006 | 0.021 | 0.79 | 54162 |
| 4 | rs1055582 | 39700173 | C | T | 0.51 | 0.014 | 0.002 | 2.1E-09 | 291092 | 0.014 | 0.016 | 0.40 | 54162 |
| 4 | rs2159935 | 55521017 | A | G | 0.49 | 0.014 | 0.002 | 1.1E-08 | 291741 | 0.014 | 0.015 | 0.38 | 54162 |
| 4 | rs1718859 | 57948224 | C | T | 0.58 | 0.012 | 0.002 | 6.7E-07 | 290546 | 0.007 | 0.016 | 0.69 | 54162 |
| 4 | rs13111599 | 83917037 | G | A | 0.74 | 0.013 | 0.003 | 3.1E-06 | 291286 | -0.021 | 0.018 | 0.23 | 54162 |
| 4 | rs10489044 | 87976387 | A | G | 0.80 | 0.018 | 0.003 | 1.7E-09 | 290838 | -0.024 | 0.019 | 0.21 | 54162 |
| 4 | rs28824216 | 92599431 | T | C | 0.02 | 0.037 | 0.008 | 1.1E-05 | 292389 | -0.040 | 0.055 | 0.47 | 54162 |
| 4 | rs1583974 | 100287812 | C | G | 0.58 | 0.020 | 0.002 | 2.5E-15 | 283704 | -0.008 | 0.017 | 0.62 | 54162 |
| 4 | rs13107325 | 103188709 | C | T | 0.93 | 0.075 | 0.005 | 2.0E-61 | 292854 | -0.051 | 0.031 | 0.10 | 54162 |
| 4 | rs13144764 | 104235669 | A | G | 0.80 | 0.016 | 0.003 | 2.0E-07 | 291323 | 0.001 | 0.021 | 0.96 | 54162 |
| 4 | rs9884482 | 106081636 | T | C | 0.63 | 0.016 | 0.002 | 2.0E-10 | 292854 | -0.016 | 0.016 | 0.31 | 54162 |
| 4 | rs17039171 | 109559063 | A | G | 0.97 | 0.031 | 0.007 | 1.5E-05 | 292098 | -0.025 | 0.048 | 0.60 | 54162 |
| 4 | rs78025076 | 110569620 | C | T | 0.98 | 0.065 | 0.008 | 7.7E-15 | 292854 | -0.253 | 0.135 | 0.06 | 54162 |
| 4 | rs72729623 | 154208278 | C | T | 0.85 | 0.018 | 0.003 | 1.5E-07 | 292361 | -0.040 | 0.023 | 0.08 | 54162 |
| 4 | rs10023962 | 177959511 | G | T | 0.82 | 0.016 | 0.003 | 2.1E-07 | 290499 | 0.001 | 0.023 | 0.97 | 54162 |
| 5 | rs7700617 | 39546706 | C | A | 0.50 | 0.010 | 0.002 | 1.8E-05 | 290235 | -0.003 | 0.016 | 0.85 | 54162 |
| 5 | rs62369484 | 43791149 | T | C | 0.94 | 0.034 | 0.005 | 6.9E-11 | 292398 | -0.086 | 0.041 | 0.04 | 54162 |
| 5 | rs116006942 | 53405314 | G | A | 0.94 | 0.024 | 0.005 | 3.2E-06 | 289872 | 0.060 | 0.035 | 0.08 | 54162 |
| 5 | rs3936511 | 55860781 | A | G | 0.81 | 0.022 | 0.003 | 5.1E-13 | 292694 | -0.036 | 0.020 | 0.07 | 54162 |
| 5 | rs1862205 | 108656635 | A | G | 0.40 | 0.009 | 0.002 | 2.4E-04 | 291538 | -0.017 | 0.016 | 0.29 | 54162 |
| 5 | rs1540687 | 111246489 | T | A | 0.68 | 0.016 | 0.003 | 3.0E-10 | 290136 | 0.005 | 0.016 | 0.74 | 54162 |
| 5 | rs1045241 | 118729286 | T | C | 0.27 | 0.013 | 0.003 | 2.0E-06 | 288740 | 0.007 | 0.019 | 0.70 | 54162 |
| 5 | rs3749748 | 127350549 | T | C | 0.25 | 0.022 | 0.003 | 1.1E-14 | 290614 | 0.009 | 0.019 | 0.65 | 54162 |
| 5 | rs248653 | 130656028 | T | A | 0.96 | 0.024 | 0.006 | 1.9E-04 | 292252 | 0.093 | 0.070 | 0.19 | 54162 |
| 5 | rs254559 | 134444982 | C | A | 0.59 | 0.020 | 0.002 | 6.6E-16 | 291966 | 0.004 | 0.016 | 0.81 | 54162 |
| 5 | rs32578 | 149211868 | A | G | 0.31 | 0.012 | 0.003 | 8.4E-06 | 292691 | 0.005 | 0.017 | 0.76 | 54162 |
| 5 | rs286965 | 153360230 | T | C | 0.37 | 0.018 | 0.002 | 7.7E-13 | 292794 | 0.005 | 0.016 | 0.74 | 54162 |
| 5 | rs2963472 | 157999022 | G | A | 0.79 | 0.017 | 0.003 | 1.8E-08 | 286797 | -0.013 | 0.019 | 0.49 | 54162 |
| 5 | rs55801554 | 158622532 | A | C | 0.24 | 0.014 | 0.003 | 1.2E-06 | 291391 | 0.009 | 0.017 | 0.59 | 54162 |
| 5 | rs351862 | 176541370 | T | C | 0.11 | 0.019 | 0.004 | 3.5E-07 | 292144 | 0.000 | 0.024 | 0.99 | 54162 |
| 5 | rs62405458 | 180225919 | C | T | 0.82 | 0.016 | 0.003 | 4.2E-07 | 290421 | 0.018 | 0.025 | 0.47 | 54162 |
| 6 | rs66757203 | 26454956 | C | T | 0.88 | 0.025 | 0.004 | 7.9E-12 | 292652 | 0.020 | 0.030 | 0.49 | 54162 |
| 6 | rs35715914 | 27592003 | C | T | 0.89 | 0.028 | 0.004 | 2.2E-13 | 290193 | -0.006 | 0.031 | 0.85 | 54162 |
| 6 | rs59137082 | 33732365 | T | C | 0.24 | 0.014 | 0.003 | 7.6E-07 | 292678 | 0.009 | 0.018 | 0.63 | 54162 |
| 6 | rs114760566 | 34192036 | C | A | 0.96 | 0.049 | 0.006 | 7.4E-17 | 292072 | 0.094 | 0.046 | 0.04 | 54162 |
| 6 | rs3800461 | 34616322 | G | C | 0.88 | 0.036 | 0.004 | 1.3E-21 | 292854 | 0.040 | 0.032 | 0.21 | 54162 |
| 6 | rs2395617 | 35285720 | A | C | 0.12 | 0.026 | 0.004 | 1.6E-12 | 292854 | -0.029 | 0.024 | 0.23 | 54162 |
| 6 | rs10947786 | 39156410 | A | G | 0.21 | 0.015 | 0.003 | 3.3E-07 | 289558 | 0.028 | 0.019 | 0.15 | 54162 |
| 6 | rs9471972 | 42915021 | A | G | 0.53 | 0.027 | 0.002 | 6.0E-29 | 290959 | 0.016 | 0.016 | 0.32 | 54162 |
| 6 | rs12204488 | 43764359 | T | C | 0.24 | 0.013 | 0.003 | 3.3E-06 | 288125 | 0.029 | 0.019 | 0.13 | 54162 |
| 6 | rs6458867 | 52629980 | G | A | 0.64 | 0.011 | 0.003 | 9.0E-06 | 291623 | -0.010 | 0.017 | 0.54 | 54162 |
| 6 | rs7757193 | 109510972 | G | A | 0.64 | 0.013 | 0.003 | 1.5E-07 | 288795 | 0.048 | 0.016 | 0.00 | 54162 |
| 6 | rs3798233 | 116317092 | C | A | 0.40 | 0.017 | 0.002 | 3.6E-12 | 292469 | -0.020 | 0.016 | 0.20 | 54162 |
| 6 | rs1406982 | 117236853 | T | A | 0.68 | 0.014 | 0.003 | 6.6E-08 | 291065 | -0.011 | 0.017 | 0.49 | 54162 |
| 6 | rs2246012 | 131898208 | T | C | 0.83 | 0.015 | 0.003 | 1.4E-06 | 292854 | -0.040 | 0.022 | 0.06 | 54162 |
| 6 | rs2982521 | 139835329 | T | A | 0.63 | 0.016 | 0.002 | 1.8E-10 | 292810 | -0.003 | 0.016 | 0.84 | 54162 |
| 6 | rs41272086 | 161008646 | G | A | 0.89 | 0.036 | 0.004 | 7.7E-20 | 292242 | 0.052 | 0.028 | 0.06 | 54162 |
| 6 | rs9347737 | 163740322 | A | G | 0.57 | 0.013 | 0.002 | 9.0E-08 | 288112 | 0.027 | 0.016 | 0.10 | 54162 |

| | | | | | | | | | | | | | |
|---|---|---|---|---|---|---|---|---|---|---|---|---|---|
| 7 | rs6969773 | 980566 | T | C | 0.54 | 0.012 | 0.002 | 1.3E-06 | 289564 | 0.034 | 0.026 | 0.18 | 54162 |
| 7 | rs9769088 | 1029585 | T | C | 0.61 | 0.022 | 0.002 | 3.8E-18 | 286509 | 0.008 | 0.017 | 0.64 | 54162 |
| 7 | rs7784748 | 4784652 | C | T | 0.57 | 0.011 | 0.002 | 4.0E-06 | 292854 | -0.020 | 0.019 | 0.29 | 54162 |
| 7 | rs10950390 | 12224708 | C | T | 0.80 | 0.013 | 0.003 | 1.7E-05 | 288087 | 0.026 | 0.019 | 0.18 | 54162 |
| 7 | rs17138358 | 17920253 | G | C | 0.60 | 0.026 | 0.002 | 1.6E-25 | 291736 | 0.011 | 0.016 | 0.47 | 54162 |
| 7 | rs76456334 | 25827967 | T | C | 0.96 | 0.036 | 0.006 | 7.9E-09 | 290206 | 0.023 | 0.051 | 0.65 | 54162 |
| 7 | rs2700892 | 36171620 | A | C | 0.55 | 0.013 | 0.002 | 7.2E-08 | 290420 | 0.018 | 0.015 | 0.25 | 54162 |
| 7 | rs112942650 | 44819240 | A | T | 0.92 | 0.023 | 0.005 | 4.7E-07 | 292022 | -0.020 | 0.032 | 0.53 | 54162 |
| 7 | rs34767118 | 50271064 | G | A | 0.33 | 0.017 | 0.003 | 4.4E-11 | 290432 | 0.029 | 0.017 | 0.08 | 54162 |
| 7 | rs55710224 | 100698648 | G | A | 0.50 | 0.012 | 0.002 | 4.0E-07 | 291364 | 0.001 | 0.016 | 0.97 | 54162 |
| 7 | rs1364422 | 130445981 | C | T | 0.72 | 0.029 | 0.003 | 2.7E-27 | 290771 | 0.001 | 0.018 | 0.96 | 54162 |
| 7 | rs11973318 | 134681306 | T | C | 0.86 | 0.016 | 0.003 | 3.8E-06 | 289130 | -0.029 | 0.022 | 0.18 | 54162 |
| 7 | rs6467595 | 135247752 | T | C | 0.77 | 0.017 | 0.003 | 8.2E-09 | 289623 | 0.027 | 0.020 | 0.17 | 54162 |
| 7 | rs6977416 | 150542711 | G | A | 0.67 | 0.017 | 0.003 | 8.1E-11 | 283431 | -0.008 | 0.017 | 0.63 | 54162 |
| 8 | rs49675 | 9151688 | A | G | 0.10 | 0.025 | 0.004 | 4.2E-10 | 292189 | -0.026 | 0.027 | 0.35 | 54162 |
| 8 | rs1986868 | 10598344 | G | A | 0.28 | 0.016 | 0.003 | 4.1E-09 | 287654 | 0.024 | 0.018 | 0.18 | 54162 |
| 8 | rs804267 | 11629241 | G | A | 0.32 | 0.019 | 0.003 | 1.3E-13 | 288735 | 0.006 | 0.017 | 0.73 | 54162 |
| 8 | rs4474021 | 12632903 | G | T | 0.68 | 0.013 | 0.003 | 3.7E-07 | 292854 | 0.006 | 0.019 | 0.75 | 54162 |
| 8 | rs11780610 | 18259876 | C | T | 0.29 | 0.011 | 0.003 | 6.8E-05 | 292049 | 0.003 | 0.017 | 0.86 | 54162 |
| 8 | rs59347135 | 19750044 | C | G | 0.96 | 0.080 | 0.006 | 1.1E-39 | 287707 | 0.034 | 0.080 | 0.67 | 54162 |
| 8 | rs139915535 | 19766233 | A | G | 0.98 | 0.155 | 0.009 | 3.8E-65 | 292752 | 0.052 | 0.092 | 0.58 | 54162 |
| 8 | rs331 | 19820405 | A | G | 0.26 | 0.091 | 0.003 | 9.4E-243 | 292364 | 0.011 | 0.017 | 0.53 | 54162 |
| 8 | rs74444445 | 19852491 | T | C | 0.98 | 0.063 | 0.008 | 9.7E-14 | 288000 | -0.019 | 0.101 | 0.85 | 54162 |
| 8 | rs56090699 | 19998949 | C | T | 0.30 | 0.015 | 0.003 | 1.3E-08 | 287266 | 0.003 | 0.018 | 0.89 | 54162 |
| 8 | rs10504477 | 71338185 | T | C | 0.59 | 0.014 | 0.002 | 2.2E-08 | 289092 | -0.014 | 0.016 | 0.37 | 54162 |
| 8 | rs36096231 | 72399623 | C | T | 0.93 | 0.019 | 0.005 | 2.9E-05 | 292279 | -0.003 | 0.033 | 0.92 | 54162 |
| 8 | rs61596977 | 95997165 | C | T | 0.86 | 0.012 | 0.003 | 7.5E-04 | 290946 | 0.063 | 0.023 | 0.01 | 54162 |
| 8 | rs2247355 | 103876780 | T | C | 0.18 | 0.015 | 0.003 | 6.4E-07 | 292169 | -0.001 | 0.020 | 0.96 | 54162 |
| 8 | rs6469605 | 116601894 | T | C | 0.57 | 0.036 | 0.002 | 3.3E-49 | 291341 | 0.026 | 0.016 | 0.11 | 54162 |
| 8 | rs17740942 | 116891360 | A | T | 0.10 | 0.017 | 0.004 | 1.4E-05 | 281013 | -0.039 | 0.051 | 0.44 | 54162 |
| 8 | rs10955991 | 121867780 | T | C | 0.32 | 0.020 | 0.003 | 1.0E-14 | 291812 | -0.015 | 0.017 | 0.35 | 54162 |
| 8 | rs72647336 | 126445055 | G | A | 0.96 | 0.018 | 0.006 | 2.7E-03 | 275476 | 0.009 | 0.079 | 0.91 | 54162 |
| 8 | rs4871603 | 126480367 | T | C | 0.66 | 0.018 | 0.003 | 2.3E-12 | 292125 | 0.010 | 0.017 | 0.57 | 54162 |
| 8 | rs12546096 | 126628841 | A | G | 0.75 | 0.016 | 0.003 | 3.4E-08 | 282156 | -0.023 | 0.019 | 0.23 | 54162 |
| 8 | rs7817574 | 144302570 | C | T | 0.18 | 0.039 | 0.003 | 7.0E-36 | 292854 | 0.021 | 0.025 | 0.41 | 54162 |
| 8 | rs4875043 | 144496772 | A | C | 0.79 | 0.011 | 0.003 | 1.7E-04 | 282814 | -0.002 | 0.023 | 0.93 | 54162 |
| 9 | rs686030 | 15304782 | A | C | 0.86 | 0.055 | 0.003 | 2.3E-57 | 292318 | 0.013 | 0.022 | 0.57 | 54162 |
| 9 | rs7036107 | 92177897 | A | G | 0.49 | 0.009 | 0.002 | 2.2E-04 | 270183 | -0.019 | 0.016 | 0.24 | 54162 |
| 9 | rs2297402 | 107579880 | C | T | 0.98 | 0.069 | 0.008 | 4.0E-16 | 288922 | 0.114 | 0.067 | 0.09 | 54162 |
| 9 | rs2066714 | 107586753 | C | T | 0.13 | 0.054 | 0.004 | 8.8E-51 | 292854 | -0.082 | 0.033 | 0.01 | 54162 |
| 9 | rs11789603 | 107647019 | T | C | 0.11 | 0.073 | 0.004 | 1.4E-79 | 292099 | 0.021 | 0.026 | 0.43 | 54162 |
| 9 | rs2740488 | 107661742 | A | C | 0.74 | 0.082 | 0.003 | 4.7E-197 | 291420 | -0.052 | 0.018 | 2.9E-03 | 54162 |
| 9 | rs10978335 | 108661116 | G | A | 0.51 | 0.013 | 0.002 | 8.1E-08 | 283302 | -0.009 | 0.016 | 0.57 | 54162 |
| 9 | rs74913239 | 114298364 | G | A | 0.84 | 0.015 | 0.003 | 3.3E-06 | 292641 | 0.014 | 0.022 | 0.52 | 54162 |
| 9 | rs4979372 | 117140082 | C | T | 0.49 | 0.015 | 0.002 | 2.3E-10 | 286090 | -0.018 | 0.016 | 0.26 | 54162 |
| 9 | rs635634 | 136155000 | T | C | 0.19 | 0.030 | 0.003 | 2.3E-22 | 292854 | -0.006 | 0.020 | 0.78 | 54162 |
| 9 | rs2520096 | 136919416 | G | A | 0.27 | 0.013 | 0.003 | 2.5E-06 | 290088 | -0.005 | 0.018 | 0.79 | 54162 |
| 10 | rs75406471 | 5257647 | G | A | 0.84 | 0.025 | 0.003 | 8.4E-14 | 292854 | 0.009 | 0.022 | 0.68 | 54162 |
| 10 | rs2804894 | 33647091 | A | G | 0.74 | 0.017 | 0.003 | 1.7E-09 | 287309 | 0.001 | 0.018 | 0.96 | 54162 |
| 10 | rs12411959 | 34015681 | A | T | 0.78 | 0.016 | 0.003 | 2.9E-08 | 290459 | -0.022 | 0.022 | 0.31 | 54162 |
| 10 | rs3802548 | 45952745 | A | T | 0.24 | 0.031 | 0.003 | 4.3E-29 | 291721 | 0.003 | 0.018 | 0.85 | 54162 |
| 10 | rs78937603 | 51580467 | G | C | 0.14 | 0.016 | 0.003 | 6.4E-06 | 292746 | 0.001 | 0.023 | 0.96 | 54162 |
| 10 | rs3915932 | 80941936 | C | G | 0.42 | 0.015 | 0.002 | 3.0E-10 | 292261 | -0.016 | 0.016 | 0.30 | 54162 |
| 10 | rs10458643 | 81152268 | G | A | 0.53 | 0.014 | 0.002 | 2.7E-08 | 283159 | 0.009 | 0.017 | 0.61 | 54162 |
| 10 | rs11202154 | 88511303 | C | T | 0.73 | 0.017 | 0.003 | 3.1E-10 | 289267 | -0.012 | 0.018 | 0.49 | 54162 |
| 10 | rs12263369 | 94823343 | C | T | 0.41 | 0.015 | 0.002 | 9.4E-10 | 292096 | 0.007 | 0.017 | 0.67 | 54162 |
| 10 | rs12357890 | 99762693 | A | G | 0.44 | 0.013 | 0.002 | 1.2E-07 | 284812 | -0.001 | 0.016 | 0.97 | 54162 |
| 10 | rs10883451 | 101924418 | T | C | 0.50 | 0.023 | 0.002 | 2.5E-21 | 292619 | 0.026 | 0.016 | 0.11 | 54162 |
| 10 | rs2792751 | 113940329 | T | C | 0.27 | 0.038 | 0.003 | 2.4E-44 | 292854 | 0.017 | 0.017 | 0.32 | 54162 |
| 10 | rs2419605 | 113978097 | A | G | 0.85 | 0.034 | 0.003 | 4.5E-23 | 290538 | 0.002 | 0.022 | 0.93 | 54162 |
| 10 | rs72823014 | 115786236 | A | G | 0.13 | 0.023 | 0.004 | 1.5E-10 | 291423 | 0.007 | 0.069 | 0.92 | 54162 |

| | | | | | | | | | | | | | |
|---|---|---|---|---|---|---|---|---|---|---|---|---|---|
| 10 | rs4450131 | 126383363 | C | T | 0.53 | 0.013 | 0.002 | 2.8E-08 | 289326 | 0.031 | 0.016 | 0.05 | 54162 |
| 10 | rs11245482 | 126733546 | T | C | 0.61 | 0.012 | 0.002 | 8.8E-07 | 291542 | 0.006 | 0.017 | 0.72 | 54162 |
| 11 | rs450244 | 2940492 | C | T | 0.91 | 0.033 | 0.004 | 1.3E-14 | 291636 | -0.044 | 0.029 | 0.12 | 54162 |
| 11 | rs11601507 | 5701074 | C | A | 0.93 | 0.031 | 0.005 | 2.2E-11 | 292854 | 0.025 | 0.044 | 0.58 | 54162 |
| 11 | rs9704692 | 10366029 | T | C | 0.46 | 0.011 | 0.002 | 7.2E-06 | 278704 | -0.024 | 0.016 | 0.14 | 54162 |
| 11 | rs2923078 | 10381766 | A | T | 0.72 | 0.022 | 0.003 | 4.5E-16 | 292527 | 0.021 | 0.017 | 0.22 | 54162 |
| 11 | rs1919309 | 14029854 | C | T | 0.51 | 0.014 | 0.002 | 1.0E-08 | 288925 | 0.016 | 0.016 | 0.30 | 54162 |
| 11 | rs12273363 | 27744859 | T | C | 0.79 | 0.021 | 0.003 | 1.1E-12 | 291768 | -0.026 | 0.020 | 0.19 | 54162 |
| 11 | rs12292135 | 32470304 | G | A | 0.69 | 0.010 | 0.003 | 1.7E-04 | 291679 | -0.097 | 0.065 | 0.13 | 54162 |
| 11 | rs7943699 | 46320022 | G | A | 0.52 | 0.016 | 0.002 | 1.2E-11 | 285719 | 0.032 | 0.016 | 0.05 | 54162 |
| 11 | rs2269434 | 47360412 | C | T | 0.35 | 0.035 | 0.003 | 2.0E-43 | 292854 | 0.002 | 0.017 | 0.93 | 54162 |
| 11 | rs3740688 | 47380340 | T | G | 0.54 | 0.013 | 0.002 | 1.2E-07 | 290205 | 0.072 | 0.016 | 0.00 | 54162 |
| 11 | rs145276599 | 48444711 | C | T | 0.11 | 0.036 | 0.004 | 1.8E-21 | 292582 | 0.020 | 0.033 | 0.55 | 54162 |
| 11 | rs73457437 | 49500475 | G | A | 0.97 | 0.034 | 0.007 | 6.2E-07 | 292825 | 0.032 | 0.047 | 0.49 | 54162 |
| 11 | rs12224170 | 49616839 | A | G | 0.13 | 0.031 | 0.004 | 1.5E-18 | 292491 | 0.030 | 0.025 | 0.22 | 54162 |
| 11 | rs12422125 | 50619511 | A | G | 0.11 | 0.034 | 0.004 | 4.4E-19 | 288416 | 0.026 | 0.028 | 0.36 | 54162 |
| 11 | rs11228871 | 54846440 | C | A | 0.13 | 0.031 | 0.004 | 5.0E-18 | 291454 | 0.017 | 0.029 | 0.57 | 54162 |
| 11 | rs11227582 | 55995736 | A | C | 0.13 | 0.031 | 0.004 | 2.9E-17 | 292285 | 0.041 | 0.035 | 0.25 | 54162 |
| 11 | rs36090449 | 57053998 | A | G | 0.05 | 0.030 | 0.006 | 1.4E-07 | 292854 | 0.016 | 0.038 | 0.67 | 54162 |
| 11 | rs174566 | 61592362 | A | G | 0.65 | 0.036 | 0.003 | 7.7E-46 | 292323 | 0.009 | 0.016 | 0.58 | 54162 |
| 11 | rs56271783 | 64004723 | G | C | 0.96 | 0.038 | 0.006 | 2.3E-10 | 290549 | -0.033 | 0.042 | 0.44 | 54162 |
| 11 | rs557675 | 65566719 | G | T | 0.47 | 0.013 | 0.002 | 2.6E-08 | 290494 | 0.002 | 0.016 | 0.88 | 54162 |
| 11 | rs4930352 | 66066993 | T | G | 0.50 | 0.018 | 0.002 | 1.0E-12 | 270592 | -0.008 | 0.016 | 0.63 | 54162 |
| 11 | rs36089024 | 67244644 | T | C | 0.43 | 0.014 | 0.002 | 2.0E-08 | 292704 | 0.016 | 0.016 | 0.30 | 54162 |
| 11 | rs11605837 | 68597886 | G | T | 0.69 | 0.018 | 0.003 | 3.3E-12 | 290890 | -0.008 | 0.017 | 0.63 | 54162 |
| 11 | rs7119167 | 73228685 | C | T | 0.21 | 0.015 | 0.003 | 2.9E-07 | 291907 | -0.028 | 0.019 | 0.14 | 54162 |
| 11 | rs559355 | 75451281 | A | T | 0.84 | 0.045 | 0.003 | 2.2E-42 | 292636 | -0.008 | 0.021 | 0.68 | 54162 |
| 11 | rs11021232 | 95320808 | T | C | 0.82 | 0.020 | 0.003 | 3.9E-10 | 290421 | -0.024 | 0.021 | 0.25 | 54162 |
| 11 | rs142958146 | 116660008 | A | G | 0.99 | 0.099 | 0.015 | 8.5E-11 | 292046 | -0.142 | 0.097 | 0.15 | 54162 |
| 11 | rs17520254 | 116665553 | G | C | 0.06 | 0.059 | 0.005 | 7.9E-32 | 292854 | 0.046 | 0.037 | 0.22 | 54162 |
| 11 | rs613808 | 116710968 | A | G | 0.28 | 0.084 | 0.003 | 2.1E-211 | 287178 | 0.030 | 0.018 | 0.09 | 54162 |
| 11 | rs515756 | 117201165 | G | T | 0.28 | 0.014 | 0.003 | 4.4E-07 | 290933 | 0.018 | 0.017 | 0.31 | 54162 |
| 11 | rs514924 | 118346863 | A | G | 0.10 | 0.017 | 0.004 | 1.6E-05 | 291766 | -0.045 | 0.026 | 0.08 | 54162 |
| 11 | rs58473820 | 122514403 | T | C | 0.38 | 0.022 | 0.002 | 2.1E-19 | 291074 | 0.032 | 0.016 | 0.04 | 54162 |
| 12 | rs7134035 | 6733889 | T | C | 0.82 | 0.017 | 0.003 | 1.1E-07 | 284691 | 0.032 | 0.022 | 0.15 | 54162 |
| 12 | rs4762756 | 20597577 | C | T | 0.74 | 0.015 | 0.003 | 4.1E-08 | 283041 | 0.000 | 0.018 | 1.00 | 54162 |
| 12 | rs11170517 | 53753909 | C | T | 0.86 | 0.020 | 0.004 | 9.7E-09 | 291877 | -0.074 | 0.024 | 0.00 | 54162 |
| 12 | rs73119306 | 57826982 | G | A | 0.24 | 0.031 | 0.003 | 4.8E-29 | 292068 | 0.009 | 0.019 | 0.62 | 54162 |
| 12 | rs7304603 | 71114400 | T | C | 0.46 | 0.014 | 0.002 | 2.1E-08 | 288588 | -0.013 | 0.016 | 0.41 | 54162 |
| 12 | rs10748165 | 71372390 | T | C | 0.50 | 0.012 | 0.002 | 2.6E-07 | 291740 | 0.019 | 0.015 | 0.21 | 54162 |
| 12 | rs10745954 | 103483094 | A | G | 0.52 | 0.015 | 0.002 | 2.3E-10 | 292854 | 0.005 | 0.016 | 0.74 | 54162 |
| 12 | rs11067231 | 109993603 | A | C | 0.52 | 0.024 | 0.002 | 2.2E-23 | 292820 | 0.022 | 0.015 | 0.15 | 54162 |
| 12 | rs3184504 | 111884608 | C | T | 0.52 | 0.017 | 0.002 | 2.3E-12 | 292854 | 0.025 | 0.016 | 0.11 | 54162 |
| 12 | rs11066320 | 112906415 | G | A | 0.58 | 0.014 | 0.002 | 1.2E-08 | 290840 | 0.009 | 0.016 | 0.56 | 54162 |
| 12 | rs7304462 | 115894190 | A | G | 0.56 | 0.015 | 0.002 | 8.0E-10 | 291675 | 0.001 | 0.016 | 0.96 | 54162 |
| 12 | rs7953249 | 121403724 | G | A | 0.41 | 0.017 | 0.002 | 1.5E-12 | 292854 | -0.022 | 0.016 | 0.16 | 54162 |
| 12 | rs56263064 | 122631964 | G | C | 0.28 | 0.014 | 0.003 | 1.5E-07 | 278735 | 0.010 | 0.018 | 0.57 | 54162 |
| 12 | rs10846497 | 123761686 | G | A | 0.09 | 0.048 | 0.004 | 1.6E-30 | 292626 | -0.020 | 0.032 | 0.53 | 54162 |
| 12 | rs11057390 | 124383813 | G | T | 0.29 | 0.018 | 0.003 | 2.3E-11 | 289620 | -0.005 | 0.017 | 0.79 | 54162 |
| 12 | rs1716393 | 124560456 | G | T | 0.40 | 0.025 | 0.002 | 2.4E-24 | 291859 | 0.007 | 0.016 | 0.66 | 54162 |
| 12 | rs921919 | 125265201 | G | A | 0.32 | 0.037 | 0.003 | 4.9E-44 | 273804 | 0.004 | 0.018 | 0.83 | 54162 |
| 12 | rs7136506 | 125326153 | T | C | 0.79 | 0.033 | 0.003 | 1.4E-28 | 279438 | 0.008 | 0.021 | 0.71 | 54162 |
| 12 | rs12229372 | 125339990 | C | T | 0.10 | 0.028 | 0.004 | 7.9E-12 | 283310 | -0.069 | 0.035 | 0.05 | 54162 |
| 12 | rs79009737 | 133124753 | A | G | 0.14 | 0.020 | 0.003 | 1.2E-08 | 290034 | 0.011 | 0.025 | 0.67 | 54162 |
| 13 | rs12867032 | 30963923 | T | A | 0.95 | 0.030 | 0.005 | 3.0E-08 | 292619 | 0.040 | 0.036 | 0.26 | 54162 |
| 13 | rs117230571 | 41689067 | A | G | 0.93 | 0.021 | 0.005 | 7.0E-06 | 289727 | 0.011 | 0.032 | 0.73 | 54162 |
| 13 | rs183078 | 49513352 | G | A | 0.40 | 0.015 | 0.002 | 2.3E-10 | 292626 | 0.005 | 0.016 | 0.77 | 54162 |
| 13 | rs3897473 | 95253920 | T | G | 0.38 | 0.009 | 0.002 | 1.3E-04 | 292854 | 0.008 | 0.016 | 0.63 | 54162 |
| 13 | rs79506257 | 103553567 | A | G | 0.05 | 0.027 | 0.006 | 2.5E-06 | 290819 | -0.026 | 0.046 | 0.57 | 54162 |
| 13 | rs9559901 | 111529619 | C | A | 0.34 | 0.011 | 0.003 | 3.3E-05 | 288504 | 0.025 | 0.017 | 0.15 | 54162 |
| 14 | rs12436555 | 24634825 | G | A | 0.83 | 0.014 | 0.003 | 1.8E-05 | 291473 | -0.024 | 0.022 | 0.27 | 54162 |

| | | | | | | | | | | | | | |
|---|---|---|---|---|---|---|---|---|---|---|---|---|---|
| 14 | rs111849006 | 52581893 | A | G | 0.18 | 0.017 | 0.003 | 1.2E-07 | 279442 | 0.009 | 0.021 | 0.66 | 54162 |
| 14 | rs13379043 | 74250126 | C | T | 0.27 | 0.021 | 0.003 | 1.4E-13 | 275130 | 0.030 | 0.018 | 0.10 | 54162 |
| 14 | rs2111705 | 75261641 | G | A | 0.46 | 0.015 | 0.002 | 1.8E-10 | 292678 | 0.003 | 0.015 | 0.86 | 54162 |
| 14 | rs11159261 | 77921120 | C | T | 0.53 | 0.010 | 0.002 | 1.4E-05 | 292433 | 0.021 | 0.016 | 0.19 | 54162 |
| 14 | rs1864167 | 81637074 | G | A | 0.46 | 0.016 | 0.002 | 1.3E-10 | 287996 | 0.023 | 0.015 | 0.13 | 54162 |
| 14 | rs71421262 | 103243408 | G | A | 0.59 | 0.011 | 0.002 | 1.0E-05 | 288163 | 0.025 | 0.016 | 0.12 | 54162 |
| 14 | rs2494748 | 105258892 | C | T | 0.38 | 0.033 | 0.002 | 9.1E-41 | 290374 | -0.031 | 0.019 | 0.11 | 54162 |
| 15 | rs7170463 | 41888918 | G | A | 0.31 | 0.019 | 0.003 | 1.2E-13 | 291002 | -0.033 | 0.017 | 0.05 | 54162 |
| 15 | rs147525635 | 43468698 | G | A | 0.44 | 0.012 | 0.002 | 1.1E-06 | 290735 | -0.015 | 0.017 | 0.40 | 54162 |
| 15 | rs150844304 | 43726625 | A | C | 0.98 | 0.079 | 0.008 | 1.6E-24 | 292779 | 0.021 | 0.045 | 0.64 | 54162 |
| 15 | rs4273010 | 44947434 | T | C | 0.98 | 0.061 | 0.008 | 8.4E-15 | 291388 | 0.023 | 0.062 | 0.71 | 54162 |
| 15 | rs1318175 | 58586129 | C | T | 0.84 | 0.078 | 0.003 | 1.6E-123 | 290529 | 0.045 | 0.021 | 0.03 | 54162 |
| 15 | rs4775033 | 58644087 | A | T | 0.88 | 0.053 | 0.004 | 1.5E-45 | 290521 | -0.043 | 0.024 | 0.07 | 54162 |
| 15 | rs1601933 | 58671559 | C | T | 0.53 | 0.088 | 0.002 | 1.7E-273 | 275774 | -0.047 | 0.018 | 0.01 | 54162 |
| 15 | rs11632618 | 58724706 | A | G | 0.07 | 0.111 | 0.005 | 2.7E-123 | 292854 | -0.020 | 0.043 | 0.65 | 54162 |
| 15 | rs80236739 | 58972699 | A | G | 0.95 | 0.028 | 0.005 | 2.1E-07 | 292854 | -0.025 | 0.040 | 0.54 | 54162 |
| 15 | rs117399007 | 59264999 | C | T | 0.96 | 0.031 | 0.006 | 2.7E-07 | 291321 | 0.067 | 0.046 | 0.15 | 54162 |
| 15 | rs17191491 | 60844847 | G | A | 0.43 | 0.011 | 0.002 | 4.3E-06 | 292854 | 0.021 | 0.016 | 0.19 | 54162 |
| 15 | rs16953475 | 63348087 | G | A | 0.53 | 0.021 | 0.002 | 2.9E-18 | 292854 | 0.034 | 0.016 | 0.03 | 54162 |
| 15 | rs28362901 | 74712937 | C | A | 0.91 | 0.027 | 0.004 | 2.8E-10 | 291596 | -0.024 | 0.029 | 0.40 | 54162 |
| 15 | rs34180494 | 75370012 | A | C | 0.73 | 0.014 | 0.003 | 3.1E-07 | 290024 | 0.016 | 0.017 | 0.35 | 54162 |
| 15 | rs8039305 | 91422543 | C | T | 0.48 | 0.013 | 0.002 | 1.9E-07 | 289099 | 0.022 | 0.017 | 0.21 | 54162 |
| 15 | rs1037117 | 102068658 | A | G | 0.25 | 0.011 | 0.003 | 3.7E-05 | 290366 | 0.017 | 0.021 | 0.41 | 54162 |
| 16 | rs34955778 | 15139594 | T | C | 0.58 | 0.012 | 0.002 | 5.5E-07 | 291995 | -0.005 | 0.017 | 0.76 | 54162 |
| 16 | rs33042 | 28139077 | A | G | 0.24 | 0.018 | 0.003 | 2.7E-10 | 286571 | -0.020 | 0.018 | 0.27 | 54162 |
| 16 | rs3814883 | 29994922 | C | T | 0.52 | 0.015 | 0.002 | 1.5E-09 | 288927 | 0.029 | 0.016 | 0.07 | 54162 |
| 16 | rs12598179 | 53375226 | C | G | 0.29 | 0.012 | 0.003 | 3.2E-06 | 284082 | -0.035 | 0.019 | 0.07 | 54162 |
| 16 | rs75152587 | 56579961 | G | T | 0.99 | 0.080 | 0.011 | 2.0E-13 | 292292 | 0.072 | 0.107 | 0.50 | 54162 |
| 16 | rs4784709 | 56711375 | T | A | 0.04 | 0.067 | 0.006 | 1.6E-27 | 292228 | -0.049 | 0.047 | 0.29 | 54162 |
| 16 | rs79984435 | 56834234 | G | A | 0.91 | 0.094 | 0.004 | 1.0E-112 | 292584 | 0.017 | 0.029 | 0.55 | 54162 |
| 16 | rs9989419 | 56985139 | G | A | 0.61 | 0.121 | 0.002 | 0.0E+00 | 292854 | 0.008 | 0.016 | 0.62 | 54162 |
| 16 | rs116857878 | 57052583 | T | C | 0.02 | 0.056 | 0.009 | 8.8E-10 | 288742 | 0.237 | 0.112 | 0.03 | 54162 |
| 16 | rs7186799 | 57220016 | A | C | 0.56 | 0.021 | 0.002 | 7.4E-19 | 291999 | 0.002 | 0.016 | 0.92 | 54162 |
| 16 | rs140164052 | 57542242 | G | A | 0.97 | 0.039 | 0.007 | 2.0E-08 | 291797 | 0.007 | 0.049 | 0.89 | 54162 |
| 16 | rs34830321 | 58616997 | C | T | 0.99 | 0.045 | 0.011 | 3.8E-05 | 290813 | 0.121 | 0.092 | 0.19 | 54162 |
| 16 | rs6499102 | 66906224 | G | A | 0.94 | 0.043 | 0.005 | 1.1E-17 | 292373 | -0.011 | 0.044 | 0.80 | 54162 |
| 16 | rs77234291 | 66911898 | G | A | 0.02 | 0.053 | 0.009 | 2.2E-08 | 292854 | -0.004 | 0.084 | 0.96 | 54162 |
| 16 | rs55781197 | 67940350 | G | A | 0.11 | 0.058 | 0.004 | 1.5E-54 | 292772 | 0.002 | 0.026 | 0.93 | 54162 |
| 16 | rs77631377 | 69049293 | C | T | 0.96 | 0.041 | 0.006 | 6.7E-12 | 290720 | -0.042 | 0.038 | 0.28 | 54162 |
| 16 | rs79940707 | 72181553 | T | C | 0.16 | 0.019 | 0.003 | 7.7E-09 | 291223 | -0.010 | 0.021 | 0.63 | 54162 |
| 16 | rs2925979 | 81534790 | C | T | 0.70 | 0.027 | 0.003 | 2.9E-24 | 292854 | -0.006 | 0.018 | 0.72 | 54162 |
| 16 | rs11641548 | 88039447 | A | C | 0.56 | 0.012 | 0.002 | 6.6E-07 | 288100 | -0.007 | 0.022 | 0.76 | 54162 |
| 17 | rs71355297 | 568515 | C | T | 0.77 | 0.012 | 0.003 | 2.3E-05 | 292115 | 0.007 | 0.020 | 0.71 | 54162 |
| 17 | rs12449758 | 3884026 | A | G | 0.69 | 0.012 | 0.003 | 3.0E-06 | 287977 | 0.032 | 0.017 | 0.06 | 54162 |
| 17 | rs78608249 | 7455707 | C | T | 0.71 | 0.013 | 0.003 | 1.2E-06 | 290814 | -0.019 | 0.018 | 0.29 | 54162 |
| 17 | rs7503353 | 8107979 | G | T | 0.47 | 0.015 | 0.002 | 5.0E-10 | 288831 | -0.027 | 0.016 | 0.09 | 54162 |
| 17 | rs12938061 | 16846155 | T | C | 0.56 | 0.012 | 0.002 | 3.1E-07 | 287470 | 0.024 | 0.016 | 0.14 | 54162 |
| 17 | rs8075019 | 17455192 | G | A | 0.88 | 0.022 | 0.004 | 5.5E-09 | 290573 | -0.004 | 0.024 | 0.87 | 54162 |
| 17 | rs2071379 | 26695832 | A | G | 0.41 | 0.023 | 0.002 | 1.1E-20 | 291717 | -0.001 | 0.016 | 0.96 | 54162 |
| 17 | rs9916613 | 28682453 | T | A | 0.64 | 0.015 | 0.003 | 4.3E-09 | 292178 | 0.020 | 0.016 | 0.21 | 54162 |
| 17 | rs731758 | 29723000 | G | C | 0.62 | 0.014 | 0.002 | 1.3E-08 | 288310 | -0.029 | 0.016 | 0.07 | 54162 |
| 17 | rs1877031 | 37814080 | A | G | 0.67 | 0.026 | 0.003 | 6.4E-24 | 292854 | 0.014 | 0.016 | 0.39 | 54162 |
| 17 | rs34138141 | 40781561 | G | T | 0.72 | 0.020 | 0.003 | 3.1E-14 | 292113 | -0.029 | 0.018 | 0.10 | 54162 |
| 17 | rs74456742 | 42191796 | A | G | 0.03 | 0.037 | 0.007 | 4.4E-08 | 287886 | -0.095 | 0.053 | 0.07 | 54162 |
| 17 | rs9989466 | 45635239 | C | T | 0.58 | 0.009 | 0.002 | 2.5E-04 | 283720 | -0.050 | 0.023 | 0.03 | 54162 |
| 17 | rs77542162 | 67081278 | A | G | 0.98 | 0.058 | 0.008 | 3.5E-13 | 292854 | -0.006 | 0.079 | 0.94 | 54162 |
| 17 | rs75988962 | 73824786 | T | C | 0.12 | 0.017 | 0.004 | 4.1E-06 | 290609 | 0.002 | 0.025 | 0.94 | 54162 |
| 18 | rs681869 | 2981398 | C | T | 0.30 | 0.017 | 0.003 | 3.8E-11 | 292273 | -0.015 | 0.017 | 0.39 | 54162 |
| 18 | rs4632228 | 19641009 | G | T | 0.79 | 0.018 | 0.003 | 4.4E-10 | 291980 | -0.003 | 0.020 | 0.89 | 54162 |
| 18 | rs6507716 | 21115060 | G | A | 0.49 | 0.013 | 0.002 | 2.9E-08 | 291302 | 0.011 | 0.016 | 0.49 | 54162 |
| 18 | rs1834144 | 40744790 | A | C | 0.37 | 0.008 | 0.002 | 8.7E-04 | 291299 | 0.008 | 0.016 | 0.62 | 54162 |

| | | | | | | | | | | | | | |
|---|---|---|---|---|---|---|---|---|---|---|---|---|---|
| 18 | rs150237291 | 46781014 | C | T | 0.02 | 0.053 | 0.008 | 8.7E-11 | 292360 | -0.061 | 0.082 | 0.46 | 54162 |
| 18 | rs77960347 | 47109955 | G | A | 0.01 | 0.328 | 0.010 | 1.1E-220 | 292854 | 0.024 | 0.058 | 0.67 | 54162 |
| 18 | rs8086351 | 47171888 | G | C | 0.82 | 0.099 | 0.003 | 7.4E-217 | 292296 | -0.014 | 0.021 | 0.52 | 54162 |
| 18 | rs2298624 | 47429022 | T | C | 0.13 | 0.036 | 0.004 | 7.5E-24 | 292854 | 0.013 | 0.023 | 0.57 | 54162 |
| 18 | rs41292412 | 56118358 | C | T | 0.99 | 0.053 | 0.011 | 1.9E-06 | 291916 | -0.092 | 0.087 | 0.29 | 54162 |
| 18 | rs7238484 | 57735552 | G | T | 0.73 | 0.023 | 0.003 | 7.0E-18 | 292771 | 0.031 | 0.018 | 0.08 | 54162 |
| 19 | rs12975319 | 3414088 | G | A | 0.70 | 0.012 | 0.003 | 1.4E-05 | 286874 | 0.012 | 0.018 | 0.50 | 54162 |
| 19 | rs352126 | 4149566 | T | C | 0.33 | 0.015 | 0.003 | 3.4E-08 | 268386 | -0.005 | 0.026 | 0.86 | 54162 |
| 19 | rs35137994 | 8429066 | T | C | 0.05 | 0.034 | 0.005 | 1.1E-10 | 292433 | -0.040 | 0.034 | 0.25 | 54162 |
| 19 | rs76213248 | 11269893 | T | C | 0.41 | 0.023 | 0.002 | 3.8E-21 | 288099 | -0.032 | 0.017 | 0.06 | 54162 |
| 19 | rs737338 | 11347657 | C | T | 0.97 | 0.107 | 0.007 | 1.7E-59 | 292797 | -0.023 | 0.042 | 0.59 | 54162 |
| 19 | rs7251640 | 18614935 | C | T | 0.19 | 0.015 | 0.003 | 4.9E-07 | 288374 | 0.046 | 0.022 | 0.04 | 54162 |
| 19 | rs12984021 | 33778783 | A | C | 0.90 | 0.015 | 0.004 | 1.5E-04 | 292245 | 0.015 | 0.026 | 0.55 | 54162 |
| 19 | rs8103728 | 33900257 | G | C | 0.67 | 0.022 | 0.003 | 1.3E-17 | 290999 | -0.014 | 0.017 | 0.41 | 54162 |
| 19 | rs15052 | 41813375 | C | T | 0.18 | 0.015 | 0.003 | 9.3E-07 | 291528 | -0.016 | 0.025 | 0.52 | 54162 |
| 19 | rs34942359 | 46385795 | C | G | 0.82 | 0.020 | 0.003 | 3.1E-10 | 292796 | 0.014 | 0.022 | 0.51 | 54162 |
| 19 | rs7259070 | 47562509 | T | C | 0.40 | 0.014 | 0.002 | 4.5E-08 | 281990 | 0.003 | 0.019 | 0.86 | 54162 |
| 19 | rs10419198 | 50038017 | T | C | 0.25 | 0.020 | 0.003 | 6.1E-13 | 292854 | -0.015 | 0.019 | 0.43 | 54162 |
| 19 | rs367070 | 54800500 | G | A | 0.23 | 0.038 | 0.003 | 9.8E-41 | 289761 | 0.024 | 0.026 | 0.36 | 54162 |
| 20 | rs144033177 | 571467 | A | C | 0.99 | 0.059 | 0.010 | 2.5E-09 | 291217 | -0.052 | 0.087 | 0.55 | 54162 |
| 20 | rs1132274 | 17596155 | C | A | 0.84 | 0.026 | 0.003 | 1.2E-15 | 292854 | -0.053 | 0.022 | 0.02 | 54162 |
| 20 | rs804553 | 22144775 | C | T | 0.50 | 0.011 | 0.002 | 1.0E-05 | 290543 | -0.023 | 0.016 | 0.15 | 54162 |
| 20 | rs6059958 | 30143278 | T | C | 0.17 | 0.015 | 0.003 | 3.0E-06 | 287479 | -0.025 | 0.021 | 0.22 | 54162 |
| 20 | rs1800961 | 43042364 | C | T | 0.97 | 0.145 | 0.007 | 2.9E-99 | 292854 | -0.060 | 0.047 | 0.20 | 54162 |
| 20 | rs3859588 | 46476143 | T | A | 0.79 | 0.015 | 0.003 | 8.3E-07 | 286168 | 0.004 | 0.021 | 0.86 | 54162 |
| 20 | rs74963256 | 51588847 | T | C | 0.91 | 0.022 | 0.004 | 9.3E-08 | 289449 | -0.026 | 0.032 | 0.41 | 54162 |
| 20 | rs6123685 | 55836040 | A | G | 0.25 | 0.016 | 0.003 | 2.5E-09 | 292854 | 0.055 | 0.022 | 0.01 | 54162 |
| 20 | rs6062510 | 62372148 | G | C | 0.33 | 0.018 | 0.003 | 3.0E-12 | 291875 | 0.015 | 0.018 | 0.40 | 54162 |
| 21 | rs235314 | 46271452 | C | T | 0.47 | 0.022 | 0.002 | 3.6E-19 | 292164 | -0.025 | 0.017 | 0.13 | 54162 |
| 22 | rs11089620 | 21922456 | C | G | 0.81 | 0.031 | 0.003 | 7.2E-24 | 292695 | 0.076 | 0.021 | 2.4E-04 | 54162 |
| 22 | rs55652051 | 29905277 | A | G | 0.77 | 0.011 | 0.003 | 1.7E-04 | 287971 | 0.039 | 0.019 | 0.04 | 54162 |
| 22 | rs9608972 | 30931307 | T | C | 0.76 | 0.018 | 0.003 | 3.8E-10 | 291604 | 0.009 | 0.019 | 0.62 | 54162 |
| 22 | rs8135417 | 31559255 | A | G | 0.64 | 0.009 | 0.003 | 2.0E-04 | 292083 | 0.011 | 0.016 | 0.51 | 54162 |
| 22 | rs4820346 | 39125074 | G | C | 0.69 | 0.015 | 0.003 | 6.7E-09 | 290575 | 0.025 | 0.017 | 0.15 | 54162 |
| 22 | rs141478056 | 42939927 | A | G | 0.90 | 0.018 | 0.004 | 1.5E-05 | 286644 | 0.059 | 0.045 | 0.20 | 54162 |
| 22 | rs738409 | 44324727 | C | G | 0.78 | 0.021 | 0.003 | 5.9E-13 | 292854 | 0.021 | 0.019 | 0.27 | 54162 |

**Supplementary table 6:** List of 193 SNPs used as instrumental variables for ApoB, and their association with ApoB and dementia.

| CHR | rsID | POS | Effect allele | Other allele | EAF | BETA apoB | SE apoB | Pval apoB | n apoB | BETA dementia | SE dementia | Pval dementia | n dementia |
|---|---|---|---|---|---|---|---|---|---|---|---|---|---|
| 1 | rs12046278 | 10799577 | T | C | 0.65 | 0.015 | 0.003 | 4.1E-08 | 313060 | 0.001 | 0.017 | 0.97 | 54162 |
| 1 | rs12078100 | 16512586 | G | C | 0.63 | 0.015 | 0.003 | 2.8E-08 | 309610 | 0.012 | 0.016 | 0.45 | 54162 |
| 1 | rs61775180 | 25793663 | C | T | 0.58 | 0.028 | 0.003 | 5.8E-28 | 308785 | 0.012 | 0.018 | 0.49 | 54162 |
| 1 | rs472495 | 55521313 | T | G | 0.65 | 0.040 | 0.003 | 5.6E-51 | 312386 | -0.046 | 0.018 | 0.01 | 54162 |
| 1 | rs11206517 | 55526428 | G | T | 0.03 | 0.066 | 0.007 | 1.2E-20 | 312879 | 0.019 | 0.043 | 0.66 | 54162 |
| 1 | rs17457613 | 56715853 | G | C | 0.98 | 0.034 | 0.009 | 6.4E-05 | 311515 | -0.124 | 0.071 | 0.08 | 54162 |
| 1 | rs1556562 | 93034023 | T | G | 0.79 | 0.016 | 0.003 | 8.5E-08 | 313060 | -0.031 | 0.020 | 0.12 | 54162 |
| 1 | rs6657811 | 109807283 | A | T | 0.87 | 0.129 | 0.004 | 4.7E-259 | 312000 | 0.011 | 0.024 | 0.66 | 54162 |
| 1 | rs6689611 | 109831268 | G | A | 0.99 | 0.133 | 0.012 | 3.2E-28 | 312209 | -0.104 | 0.100 | 0.30 | 54162 |
| 1 | rs55739424 | 110170640 | T | G | 0.03 | 0.036 | 0.007 | 1.6E-06 | 313060 | 0.113 | 0.064 | 0.08 | 54162 |
| 1 | rs17447211 | 182140185 | G | C | 0.33 | 0.015 | 0.003 | 2.0E-08 | 311656 | 0.004 | 0.016 | 0.80 | 54162 |
| 1 | rs6667939 | 198994619 | T | C | 0.72 | 0.016 | 0.003 | 3.0E-08 | 306194 | 0.000 | 0.018 | 0.99 | 54162 |
| 1 | rs2807854 | 221029841 | C | T | 0.67 | 0.015 | 0.003 | 1.2E-08 | 312533 | 0.038 | 0.018 | 0.04 | 54162 |
| 1 | rs10127775 | 230295789 | A | T | 0.39 | 0.022 | 0.003 | 5.6E-18 | 313060 | -0.010 | 0.017 | 0.56 | 54162 |
| 1 | rs656521 | 234799148 | A | G | 0.51 | 0.014 | 0.003 | 5.0E-08 | 308730 | 0.018 | 0.016 | 0.26 | 54162 |
| 1 | rs556107 | 234853059 | T | C | 0.52 | 0.037 | 0.003 | 8.1E-47 | 309620 | 0.017 | 0.016 | 0.28 | 54162 |
| 1 | rs6426328 | 246891260 | T | G | 0.49 | 0.013 | 0.003 | 1.3E-07 | 308633 | -0.003 | 0.017 | 0.86 | 54162 |
| 2 | rs719148 | 20521647 | G | A | 0.22 | 0.016 | 0.003 | 6.5E-08 | 313060 | -0.015 | 0.018 | 0.42 | 54162 |
| 2 | rs115704890 | 20999520 | A | T | 0.08 | 0.035 | 0.005 | 4.0E-14 | 312402 | -0.011 | 0.032 | 0.72 | 54162 |
| 2 | rs115692156 | 21077208 | A | G | 0.99 | 0.110 | 0.015 | 4.8E-13 | 309897 | 0.048 | 0.072 | 0.50 | 54162 |
| 2 | rs62122481 | 21216815 | A | C | 0.38 | 0.083 | 0.003 | 5.6E-221 | 306439 | 0.005 | 0.017 | 0.78 | 54162 |
| 2 | rs60403635 | 21272778 | T | C | 0.96 | 0.110 | 0.006 | 1.3E-69 | 312182 | 0.009 | 0.050 | 0.85 | 54162 |
| 2 | rs11693526 | 22347500 | C | T | 0.06 | 0.022 | 0.006 | 8.9E-05 | 294071 | 0.011 | 0.038 | 0.77 | 54162 |
| 2 | rs142787485 | 26358156 | A | G | 0.97 | 0.027 | 0.007 | 1.4E-04 | 306050 | -0.086 | 0.051 | 0.09 | 54162 |
| 2 | rs13394970 | 26929282 | G | T | 0.61 | 0.012 | 0.003 | 5.6E-06 | 310508 | -0.032 | 0.016 | 0.04 | 54162 |
| 2 | rs1260326 | 27730940 | T | C | 0.39 | 0.050 | 0.003 | 9.1E-84 | 313060 | -0.001 | 0.016 | 0.96 | 54162 |
| 2 | rs4299376 | 44072576 | G | T | 0.32 | 0.045 | 0.003 | 4.4E-63 | 312197 | -0.018 | 0.017 | 0.30 | 54162 |
| 2 | rs6709904 | 44080324 | A | G | 0.89 | 0.037 | 0.004 | 2.3E-20 | 312398 | 0.037 | 0.024 | 0.13 | 54162 |
| 2 | rs4671050 | 62988169 | G | T | 0.69 | 0.018 | 0.003 | 1.6E-11 | 309874 | 0.009 | 0.017 | 0.60 | 54162 |
| 2 | rs12471768 | 64928603 | C | T | 0.71 | 0.017 | 0.003 | 1.6E-09 | 311492 | 0.045 | 0.018 | 0.01 | 54162 |
| 2 | rs7601412 | 109168508 | A | G | 0.14 | 0.019 | 0.004 | 7.3E-08 | 312900 | -0.036 | 0.022 | 0.10 | 54162 |
| 2 | rs150474434 | 118845121 | G | A | 0.90 | 0.035 | 0.004 | 3.3E-16 | 309381 | 0.046 | 0.027 | 0.09 | 54162 |
| 2 | rs17050272 | 121306440 | G | A | 0.59 | 0.026 | 0.003 | 1.0E-23 | 313060 | -0.005 | 0.017 | 0.76 | 54162 |
| 2 | rs6714750 | 136783169 | G | A | 0.18 | 0.013 | 0.003 | 7.1E-05 | 299118 | 0.019 | 0.019 | 0.33 | 54162 |
| 2 | rs7601153 | 158447571 | C | G | 0.60 | 0.012 | 0.003 | 1.7E-06 | 312466 | -0.005 | 0.016 | 0.73 | 54162 |
| 2 | rs13389219 | 165528876 | C | T | 0.61 | 0.017 | 0.003 | 2.0E-11 | 312942 | 0.027 | 0.016 | 0.09 | 54162 |
| 2 | rs7569317 | 203527979 | C | T | 0.53 | 0.019 | 0.003 | 1.1E-13 | 312212 | 0.032 | 0.016 | 0.04 | 54162 |
| 2 | rs7603427 | 204317553 | T | C | 0.53 | 0.014 | 0.003 | 9.1E-08 | 309997 | 0.001 | 0.016 | 0.95 | 54162 |
| 2 | rs1250258 | 216300185 | T | C | 0.74 | 0.014 | 0.003 | 1.4E-06 | 310513 | 0.013 | 0.019 | 0.48 | 54162 |
| 2 | rs11568318 | 234665498 | A | C | 0.07 | 0.027 | 0.005 | 1.1E-07 | 312811 | 0.028 | 0.032 | 0.38 | 54162 |
| 2 | rs59104589 | 242237902 | C | T | 0.64 | 0.014 | 0.003 | 3.6E-08 | 312853 | -0.001 | 0.017 | 0.98 | 54162 |
| 3 | rs13076933 | 12327431 | T | G | 0.74 | 0.016 | 0.003 | 9.7E-08 | 307147 | -0.007 | 0.019 | 0.70 | 54162 |
| 3 | rs9834932 | 32535382 | A | G | 0.91 | 0.033 | 0.004 | 6.3E-14 | 312798 | 0.017 | 0.028 | 0.53 | 54162 |
| 3 | rs71311871 | 58420613 | A | G | 0.92 | 0.029 | 0.005 | 1.7E-10 | 312590 | -0.002 | 0.027 | 0.93 | 54162 |
| 3 | rs55921103 | 69810294 | T | G | 0.65 | 0.014 | 0.003 | 1.5E-07 | 307862 | 0.020 | 0.017 | 0.22 | 54162 |
| 3 | rs12054451 | 122064369 | G | T | 0.26 | 0.016 | 0.003 | 3.1E-08 | 308393 | -0.018 | 0.017 | 0.31 | 54162 |
| 3 | rs113177823 | 132217703 | G | A | 0.95 | 0.038 | 0.006 | 2.0E-11 | 310615 | -0.067 | 0.062 | 0.28 | 54162 |
| 3 | rs3932048 | 136258924 | G | C | 0.32 | 0.013 | 0.003 | 2.2E-06 | 311723 | 0.005 | 0.017 | 0.75 | 54162 |
| 4 | rs13108218 | 3443931 | A | G | 0.38 | 0.024 | 0.003 | 2.8E-20 | 303388 | -0.015 | 0.018 | 0.39 | 54162 |
| 4 | rs2137234 | 26080549 | C | T | 0.19 | 0.012 | 0.003 | 9.7E-05 | 312262 | -0.015 | 0.020 | 0.46 | 54162 |
| 4 | rs278981 | 40428010 | C | T | 0.76 | 0.016 | 0.003 | 3.5E-08 | 313060 | -0.017 | 0.021 | 0.42 | 54162 |
| 4 | rs72663045 | 74177397 | G | T | 0.02 | 0.041 | 0.009 | 7.9E-06 | 312341 | 0.009 | 0.053 | 0.87 | 54162 |
| 4 | rs1458038 | 81164723 | C | T | 0.71 | 0.016 | 0.003 | 5.6E-09 | 308559 | 0.031 | 0.018 | 0.09 | 54162 |
| 4 | rs2705619 | 87836652 | A | G | 0.71 | 0.017 | 0.003 | 1.7E-09 | 311596 | -0.016 | 0.017 | 0.35 | 54162 |
| 4 | rs1229984 | 100239319 | C | T | 0.98 | 0.041 | 0.009 | 1.4E-06 | 313060 | -0.044 | 0.042 | 0.29 | 54162 |
| 4 | rs58148580 | 124758773 | T | C | 0.11 | 0.019 | 0.004 | 3.7E-06 | 313060 | 0.010 | 0.024 | 0.67 | 54162 |
| 5 | rs3936511 | 55860781 | G | A | 0.19 | 0.022 | 0.003 | 7.9E-12 | 312890 | 0.036 | 0.020 | 0.07 | 54162 |
| 5 | rs2925677 | 71953629 | C | G | 0.79 | 0.017 | 0.003 | 2.1E-08 | 312560 | -0.019 | 0.019 | 0.33 | 54162 |

| | | | | | | | | | | | | | |
|---|---|---|---|---|---|---|---|---|---|---|---|---|---|
| 5 | rs12916 | 74656539 | C | T | 0.40 | 0.056 | 0.003 | 1.7E-106 | 313060 | 0.005 | 0.016 | 0.77 | 54162 |
| 5 | rs7734476 | 122848876 | A | G | 0.55 | 0.022 | 0.003 | 1.2E-17 | 312443 | 0.018 | 0.016 | 0.24 | 54162 |
| 5 | rs1003533 | 131755651 | C | T | 0.81 | 0.021 | 0.003 | 1.9E-10 | 311733 | 0.023 | 0.020 | 0.25 | 54162 |
| 5 | rs6874202 | 156391628 | C | T | 0.64 | 0.034 | 0.003 | 6.1E-38 | 312950 | 0.007 | 0.017 | 0.70 | 54162 |
| 6 | rs147539187 | 11839042 | C | G | 0.93 | 0.017 | 0.005 | 4.3E-04 | 311839 | -0.008 | 0.030 | 0.79 | 54162 |
| 6 | rs7746081 | 16126934 | G | A | 0.70 | 0.024 | 0.003 | 2.2E-18 | 311562 | -0.006 | 0.017 | 0.74 | 54162 |
| 6 | rs79220007 | 26098474 | T | C | 0.92 | 0.053 | 0.005 | 2.2E-29 | 312729 | 0.015 | 0.034 | 0.67 | 54162 |
| 6 | rs76079263 | 28019665 | G | C | 0.91 | 0.027 | 0.004 | 1.5E-09 | 307714 | -0.015 | 0.033 | 0.65 | 54162 |
| 6 | rs6907508 | 34592090 | A | G | 0.88 | 0.029 | 0.004 | 1.5E-13 | 312483 | 0.020 | 0.024 | 0.39 | 54162 |
| 6 | rs913499 | 37038432 | A | G | 0.49 | 0.011 | 0.003 | 2.1E-05 | 311928 | 0.018 | 0.016 | 0.26 | 54162 |
| 6 | rs55804343 | 39234907 | T | C | 0.30 | 0.017 | 0.003 | 4.9E-10 | 309419 | -0.007 | 0.017 | 0.69 | 54162 |
| 6 | rs1358980 | 43764551 | T | C | 0.48 | 0.014 | 0.003 | 9.6E-08 | 302025 | -0.027 | 0.017 | 0.11 | 54162 |
| 6 | rs2063643 | 52478364 | A | G | 0.82 | 0.016 | 0.003 | 8.7E-07 | 313060 | -0.022 | 0.021 | 0.30 | 54162 |
| 6 | rs9496567 | 100602753 | G | A | 0.76 | 0.021 | 0.003 | 2.8E-12 | 310943 | 0.008 | 0.019 | 0.67 | 54162 |
| 6 | rs56264193 | 110028076 | G | C | 0.64 | 0.010 | 0.003 | 6.8E-05 | 311807 | 0.030 | 0.016 | 0.07 | 54162 |
| 6 | rs3822855 | 116316882 | T | G | 0.40 | 0.017 | 0.003 | 1.3E-10 | 312394 | -0.019 | 0.016 | 0.24 | 54162 |
| 6 | rs9491697 | 127456122 | G | A | 0.47 | 0.019 | 0.003 | 9.9E-14 | 310987 | 0.023 | 0.016 | 0.16 | 54162 |
| 6 | rs12197047 | 130389211 | A | G | 0.67 | 0.016 | 0.003 | 6.5E-09 | 304409 | 0.011 | 0.018 | 0.52 | 54162 |
| 6 | rs7776054 | 135418916 | A | G | 0.74 | 0.011 | 0.003 | 2.0E-04 | 312048 | 0.000 | 0.018 | 0.99 | 54162 |
| 6 | rs73025516 | 160520806 | A | G | 0.96 | 0.032 | 0.006 | 1.7E-07 | 312383 | -0.006 | 0.043 | 0.89 | 54162 |
| 6 | rs12208357 | 160543148 | T | C | 0.07 | 0.065 | 0.005 | 4.3E-39 | 311710 | -0.084 | 0.030 | 4.6E-03 | 54162 |
| 6 | rs146534110 | 160578069 | T | G | 0.01 | 0.057 | 0.011 | 3.0E-07 | 313060 | 0.091 | 0.097 | 0.35 | 54162 |
| 6 | rs117733303 | 160922870 | G | A | 0.02 | 0.093 | 0.009 | 2.7E-23 | 313060 | -0.004 | 0.127 | 0.97 | 54162 |
| 6 | rs118039278 | 160985526 | A | G | 0.08 | 0.083 | 0.005 | 3.3E-71 | 310997 | -0.057 | 0.037 | 0.13 | 54162 |
| 7 | rs28406917 | 21449451 | T | C | 0.43 | 0.013 | 0.003 | 6.5E-07 | 308562 | -0.006 | 0.016 | 0.70 | 54162 |
| 7 | rs4470903 | 21604916 | G | C | 0.22 | 0.032 | 0.003 | 1.2E-25 | 312465 | -0.010 | 0.020 | 0.63 | 54162 |
| 7 | rs4722551 | 25991826 | C | T | 0.16 | 0.027 | 0.003 | 1.4E-14 | 313060 | -0.022 | 0.022 | 0.31 | 54162 |
| 7 | rs2073547 | 44582331 | G | A | 0.18 | 0.033 | 0.003 | 2.2E-24 | 313060 | -0.025 | 0.021 | 0.24 | 54162 |
| 7 | rs13247874 | 73010442 | C | T | 0.80 | 0.022 | 0.003 | 1.1E-11 | 312410 | -0.015 | 0.020 | 0.46 | 54162 |
| 7 | rs45537841 | 87110640 | C | T | 0.82 | 0.014 | 0.003 | 1.1E-05 | 312709 | -0.034 | 0.020 | 0.09 | 54162 |
| 7 | rs112758337 | 97977268 | G | A | 0.81 | 0.018 | 0.003 | 1.5E-08 | 311430 | 0.018 | 0.020 | 0.36 | 54162 |
| 7 | rs10953298 | 100216773 | C | T | 0.77 | 0.018 | 0.003 | 2.3E-09 | 306202 | 0.004 | 0.020 | 0.86 | 54162 |
| 7 | rs13230111 | 130437124 | A | G | 0.51 | 0.014 | 0.003 | 5.0E-08 | 311856 | -0.006 | 0.016 | 0.69 | 54162 |
| 8 | rs7012637 | 9173209 | A | G | 0.47 | 0.021 | 0.003 | 3.1E-16 | 305715 | -0.006 | 0.017 | 0.72 | 54162 |
| 8 | rs1495741 | 18272881 | G | A | 0.22 | 0.019 | 0.003 | 1.1E-09 | 313060 | -0.004 | 0.019 | 0.83 | 54162 |
| 8 | rs139915535 | 19766233 | G | A | 0.02 | 0.044 | 0.010 | 4.2E-06 | 312951 | -0.052 | 0.092 | 0.58 | 54162 |
| 8 | rs13702 | 19824492 | T | C | 0.71 | 0.032 | 0.003 | 8.8E-30 | 313060 | -0.007 | 0.017 | 0.68 | 54162 |
| 8 | rs59328596 | 21928227 | G | A | 0.85 | 0.023 | 0.004 | 2.6E-10 | 312685 | -0.041 | 0.023 | 0.08 | 54162 |
| 8 | rs9298506 | 55437524 | G | A | 0.21 | 0.021 | 0.003 | 1.2E-11 | 313060 | 0.016 | 0.020 | 0.41 | 54162 |
| 8 | rs9297994 | 59392324 | G | A | 0.34 | 0.027 | 0.003 | 2.6E-24 | 311064 | 0.003 | 0.016 | 0.83 | 54162 |
| 8 | rs2737263 | 116667539 | G | T | 0.72 | 0.019 | 0.003 | 1.1E-11 | 312003 | -0.034 | 0.017 | 0.05 | 54162 |
| 8 | rs28601761 | 126500031 | C | G | 0.58 | 0.071 | 0.003 | 1.8E-163 | 298770 | 0.004 | 0.017 | 0.82 | 54162 |
| 8 | rs2124034 | 126644200 | G | T | 0.28 | 0.015 | 0.003 | 8.7E-08 | 311862 | 0.015 | 0.018 | 0.39 | 54162 |
| 8 | rs55831924 | 145031968 | T | C | 0.36 | 0.019 | 0.003 | 2.0E-12 | 305042 | 0.022 | 0.018 | 0.23 | 54162 |
| 9 | rs3780181 | 2640759 | A | G | 0.93 | 0.027 | 0.005 | 1.1E-07 | 310592 | 0.071 | 0.031 | 0.02 | 54162 |
| 9 | rs581080 | 15305378 | C | G | 0.82 | 0.017 | 0.003 | 3.8E-07 | 313060 | 0.016 | 0.021 | 0.43 | 54162 |
| 9 | rs6475606 | 22081850 | C | T | 0.52 | 0.018 | 0.003 | 4.2E-12 | 313060 | -0.032 | 0.016 | 0.04 | 54162 |
| 9 | rs6560499 | 78730766 | G | A | 0.42 | 0.013 | 0.003 | 4.2E-07 | 305917 | -0.009 | 0.016 | 0.57 | 54162 |
| 9 | rs13283282 | 131465481 | C | G | 0.85 | 0.019 | 0.004 | 9.3E-08 | 313060 | 0.011 | 0.030 | 0.73 | 54162 |
| 9 | rs10448340 | 139320069 | T | G | 0.68 | 0.017 | 0.003 | 9.0E-10 | 311028 | 0.021 | 0.018 | 0.23 | 54162 |
| 10 | rs11014154 | 18706320 | A | G | 0.28 | 0.014 | 0.003 | 5.0E-07 | 306513 | 0.029 | 0.018 | 0.11 | 54162 |
| 10 | rs17476364 | 71094504 | T | C | 0.89 | 0.021 | 0.004 | 1.0E-07 | 312364 | 0.010 | 0.028 | 0.73 | 54162 |
| 10 | rs2068888 | 94839642 | G | A | 0.55 | 0.025 | 0.003 | 8.7E-23 | 313060 | 0.021 | 0.016 | 0.20 | 54162 |
| 10 | rs79931565 | 104175649 | G | A | 0.07 | 0.025 | 0.005 | 4.7E-07 | 312620 | 0.001 | 0.033 | 0.98 | 54162 |
| 10 | rs12246352 | 124705307 | G | A | 0.10 | 0.027 | 0.004 | 1.6E-10 | 311561 | -0.020 | 0.027 | 0.46 | 54162 |
| 11 | rs7108486 | 5677158 | T | C | 0.98 | 0.046 | 0.008 | 5.0E-08 | 311837 | -0.059 | 0.063 | 0.35 | 54162 |
| 11 | rs11601507 | 5701074 | A | C | 0.07 | 0.041 | 0.005 | 1.2E-16 | 313060 | -0.025 | 0.044 | 0.58 | 54162 |
| 11 | rs10832963 | 18664241 | G | T | 0.75 | 0.021 | 0.003 | 1.4E-12 | 311201 | 0.006 | 0.018 | 0.72 | 54162 |
| 11 | rs546240 | 30533622 | C | T | 0.38 | 0.010 | 0.003 | 1.1E-04 | 310550 | 0.016 | 0.017 | 0.34 | 54162 |
| 11 | rs190104 | 32112532 | A | G | 0.14 | 0.016 | 0.004 | 7.0E-06 | 311486 | -0.010 | 0.023 | 0.66 | 54162 |
| 11 | rs174564 | 61588305 | A | G | 0.65 | 0.046 | 0.003 | 7.4E-69 | 312495 | 0.010 | 0.016 | 0.54 | 54162 |

| | | | | | | | | | | | | | |
|---|---|---|---|---|---|---|---|---|---|---|---|---|---|
| 11 | rs10896125 | 66288733 | G | C | 0.76 | 0.018 | 0.003 | 2.3E-09 | 312974 | 0.010 | 0.018 | 0.59 | 54162 |
| 11 | rs964184 | 116648917 | G | C | 0.13 | 0.076 | 0.004 | 2.3E-93 | 313060 | 0.021 | 0.023 | 0.36 | 54162 |
| 11 | rs12970 | 117074109 | G | A | 0.94 | 0.021 | 0.005 | 8.0E-05 | 312904 | -0.050 | 0.034 | 0.14 | 54162 |
| 12 | rs35882350 | 623129 | G | A | 0.26 | 0.017 | 0.003 | 2.0E-09 | 313060 | -0.006 | 0.020 | 0.78 | 54162 |
| 12 | rs2160994 | 50650057 | C | T | 0.65 | 0.018 | 0.003 | 1.4E-11 | 310911 | 0.000 | 0.018 | 0.99 | 54162 |
| 12 | rs10876450 | 53811034 | C | T | 0.18 | 0.020 | 0.003 | 1.5E-09 | 312140 | 0.061 | 0.020 | 0.00 | 54162 |
| 12 | rs2122982 | 57781893 | G | A | 0.76 | 0.020 | 0.003 | 8.1E-12 | 312727 | -0.011 | 0.019 | 0.57 | 54162 |
| 12 | rs597808 | 111973358 | G | A | 0.52 | 0.021 | 0.003 | 4.0E-16 | 310480 | 0.034 | 0.016 | 0.03 | 54162 |
| 12 | rs233721 | 113031543 | A | T | 0.65 | 0.017 | 0.003 | 6.9E-11 | 306431 | 0.028 | 0.017 | 0.09 | 54162 |
| 12 | rs1169292 | 121426478 | T | C | 0.31 | 0.022 | 0.003 | 2.3E-15 | 310331 | -0.015 | 0.017 | 0.39 | 54162 |
| 12 | rs11057397 | 124419728 | C | T | 0.67 | 0.017 | 0.003 | 1.2E-10 | 311236 | -0.007 | 0.017 | 0.68 | 54162 |
| 12 | rs112403212 | 125303254 | T | C | 0.14 | 0.021 | 0.004 | 1.2E-08 | 308411 | -0.040 | 0.025 | 0.11 | 54162 |
| 13 | rs2238162 | 32959199 | C | T | 0.48 | 0.024 | 0.003 | 7.7E-22 | 312801 | 0.005 | 0.015 | 0.73 | 54162 |
| 13 | rs4771674 | 111039070 | G | A | 0.63 | 0.015 | 0.003 | 7.1E-09 | 307741 | 0.018 | 0.016 | 0.27 | 54162 |
| 13 | rs6602909 | 114551993 | C | T | 0.33 | 0.023 | 0.003 | 3.9E-18 | 311594 | -0.013 | 0.017 | 0.47 | 54162 |
| 14 | rs11621792 | 24871926 | T | C | 0.45 | 0.021 | 0.003 | 8.7E-16 | 304649 | -0.025 | 0.018 | 0.16 | 54162 |
| 14 | rs34767236 | 70810569 | A | G | 0.38 | 0.017 | 0.003 | 1.3E-10 | 310388 | -0.006 | 0.016 | 0.72 | 54162 |
| 14 | rs10151436 | 73616095 | A | T | 0.89 | 0.020 | 0.004 | 6.8E-07 | 312496 | -0.043 | 0.025 | 0.09 | 54162 |
| 14 | rs13379043 | 74250126 | T | C | 0.73 | 0.013 | 0.003 | 4.4E-06 | 294103 | -0.030 | 0.018 | 0.10 | 54162 |
| 14 | rs145730801 | 94768196 | C | T | 0.04 | 0.038 | 0.006 | 2.1E-09 | 308872 | 0.077 | 0.045 | 0.09 | 54162 |
| 15 | rs10851478 | 49829019 | T | C | 0.58 | 0.013 | 0.003 | 4.4E-07 | 312610 | -0.007 | 0.016 | 0.65 | 54162 |
| 15 | rs72733928 | 57512284 | T | A | 0.06 | 0.027 | 0.005 | 3.4E-07 | 312031 | -0.005 | 0.031 | 0.87 | 54162 |
| 15 | rs261290 | 58678720 | T | C | 0.35 | 0.022 | 0.003 | 7.2E-17 | 312140 | -0.033 | 0.017 | 0.05 | 54162 |
| 15 | rs473224 | 58737341 | T | G | 0.14 | 0.022 | 0.004 | 3.7E-09 | 304771 | -0.005 | 0.022 | 0.83 | 54162 |
| 15 | rs41434449 | 64448460 | T | A | 0.13 | 0.020 | 0.004 | 1.4E-07 | 313060 | -0.016 | 0.032 | 0.61 | 54162 |
| 15 | rs56402930 | 91072962 | A | G | 0.09 | 0.019 | 0.004 | 9.2E-06 | 312034 | 0.028 | 0.027 | 0.29 | 54162 |
| 15 | rs4965894 | 102069043 | T | C | 0.60 | 0.009 | 0.003 | 2.9E-04 | 310839 | -0.018 | 0.020 | 0.36 | 54162 |
| 16 | rs1561139 | 56852822 | G | T | 0.58 | 0.012 | 0.003 | 1.9E-06 | 312516 | -0.014 | 0.016 | 0.37 | 54162 |
| 16 | rs3764261 | 56993324 | C | A | 0.67 | 0.048 | 0.003 | 9.4E-70 | 313060 | -0.003 | 0.017 | 0.85 | 54162 |
| 16 | rs62049427 | 70334172 | A | G | 0.06 | 0.024 | 0.005 | 8.4E-06 | 308106 | -0.107 | 0.066 | 0.11 | 54162 |
| 16 | rs34042070 | 72101525 | G | C | 0.19 | 0.052 | 0.003 | 1.4E-57 | 309296 | -0.009 | 0.020 | 0.64 | 54162 |
| 16 | rs1862719 | 79504057 | A | G | 0.24 | 0.015 | 0.003 | 1.2E-06 | 294956 | 0.051 | 0.019 | 0.01 | 54162 |
| 16 | rs12443634 | 81524274 | A | C | 0.28 | 0.012 | 0.003 | 1.5E-05 | 305263 | 0.005 | 0.018 | 0.78 | 54162 |
| 17 | rs12948283 | 1622850 | C | G | 0.29 | 0.015 | 0.003 | 1.3E-06 | 260685 | 0.050 | 0.020 | 0.01 | 54162 |
| 17 | rs55714927 | 7080316 | C | T | 0.81 | 0.034 | 0.003 | 3.8E-26 | 313060 | -0.006 | 0.028 | 0.84 | 54162 |
| 17 | rs74454529 | 17432220 | A | G | 0.06 | 0.025 | 0.005 | 2.5E-06 | 312113 | 0.004 | 0.031 | 0.89 | 54162 |
| 17 | rs704 | 26694861 | A | G | 0.47 | 0.019 | 0.003 | 1.4E-13 | 313060 | -0.006 | 0.017 | 0.73 | 54162 |
| 17 | rs2058122 | 27663848 | C | T | 0.86 | 0.020 | 0.004 | 1.0E-07 | 305831 | -0.002 | 0.024 | 0.95 | 54162 |
| 17 | rs12603885 | 29466722 | A | G | 0.70 | 0.019 | 0.003 | 1.6E-11 | 312809 | -0.029 | 0.018 | 0.10 | 54162 |
| 17 | rs112220485 | 40522713 | C | T | 0.08 | 0.022 | 0.005 | 2.3E-06 | 311093 | 0.034 | 0.030 | 0.26 | 54162 |
| 17 | rs36043200 | 45629406 | G | A | 0.48 | 0.029 | 0.003 | 1.9E-30 | 308968 | -0.053 | 0.020 | 0.01 | 54162 |
| 17 | rs3096644 | 46757575 | G | T | 0.68 | 0.019 | 0.003 | 1.1E-11 | 309505 | -0.003 | 0.017 | 0.88 | 54162 |
| 17 | rs1801689 | 64210580 | C | A | 0.03 | 0.070 | 0.007 | 4.9E-21 | 313060 | -0.144 | 0.076 | 0.06 | 54162 |
| 17 | rs60856912 | 65892343 | T | G | 0.16 | 0.015 | 0.003 | 1.4E-05 | 309356 | -0.004 | 0.021 | 0.85 | 54162 |
| 17 | rs77542162 | 67081278 | G | A | 0.02 | 0.113 | 0.008 | 1.3E-40 | 313060 | 0.006 | 0.079 | 0.94 | 54162 |
| 17 | rs72631343 | 67191270 | C | G | 0.87 | 0.030 | 0.004 | 2.2E-15 | 313060 | -0.042 | 0.026 | 0.11 | 54162 |
| 17 | rs4485425 | 73767437 | G | A | 0.72 | 0.013 | 0.003 | 3.3E-06 | 310639 | 0.010 | 0.017 | 0.55 | 54162 |
| 17 | rs12948394 | 76382791 | C | T | 0.52 | 0.013 | 0.003 | 1.3E-06 | 294149 | 0.030 | 0.016 | 0.06 | 54162 |
| 19 | rs143020224 | 11187324 | C | G | 0.88 | 0.173 | 0.004 | 0.0E+00 | 312924 | 0.019 | 0.028 | 0.49 | 54162 |
| 19 | rs2738447 | 11227480 | C | A | 0.59 | 0.045 | 0.003 | 1.3E-69 | 312356 | 0.001 | 0.016 | 0.94 | 54162 |
| 19 | rs7249565 | 11302807 | A | G | 0.42 | 0.015 | 0.003 | 1.2E-08 | 313060 | 0.014 | 0.016 | 0.38 | 54162 |
| 19 | rs62120394 | 18338709 | A | G | 0.29 | 0.018 | 0.003 | 4.0E-11 | 310388 | 0.006 | 0.017 | 0.74 | 54162 |
| 19 | rs8107974 | 19388500 | A | T | 0.92 | 0.091 | 0.005 | 8.4E-82 | 312696 | -0.005 | 0.047 | 0.92 | 54162 |
| 19 | rs56113850 | 41353107 | C | T | 0.58 | 0.014 | 0.003 | 6.3E-08 | 310532 | 0.027 | 0.023 | 0.24 | 54162 |
| 19 | rs2021092 | 44068706 | T | C | 0.81 | 0.019 | 0.003 | 1.1E-08 | 311884 | 0.014 | 0.020 | 0.49 | 54162 |
| 19 | rs62119267 | 45134682 | A | C | 0.98 | 0.342 | 0.009 | 0.0E+00 | 311428 | 0.256 | 0.132 | 0.05 | 54162 |
| 19 | rs62120573 | 45214556 | C | T | 0.07 | 0.028 | 0.005 | 1.7E-08 | 312900 | -0.053 | 0.035 | 0.13 | 54162 |
| 19 | rs73045960 | 46295223 | A | G | 0.98 | 0.141 | 0.010 | 8.3E-45 | 313060 | 0.277 | 0.086 | 1.3E-03 | 54162 |
| 19 | rs145725232 | 46752541 | G | A | 0.99 | 0.060 | 0.012 | 2.6E-07 | 308064 | 0.046 | 0.114 | 0.69 | 54162 |
| 19 | rs399970 | 57063916 | G | T | 0.25 | 0.013 | 0.003 | 1.9E-05 | 310009 | 0.012 | 0.020 | 0.55 | 54162 |
| 19 | rs35081008 | 58662235 | C | T | 0.85 | 0.034 | 0.004 | 1.8E-21 | 310579 | -0.002 | 0.028 | 0.95 | 54162 |

| | | | | | | | | | | | | | |
|---|---|---|---|---|---|---|---|---|---|---|---|---|---|
| 20 | rs73075609 | 5580789 | T | C | 0.03 | 0.050 | 0.008 | 3.1E-10 | 312199 | 0.097 | 0.061 | 0.11 | 54162 |
| 20 | rs969075 | 17792323 | C | T | 0.67 | 0.015 | 0.003 | 6.4E-08 | 306476 | 0.016 | 0.017 | 0.35 | 54162 |
| 20 | rs2618566 | 17844684 | G | T | 0.34 | 0.029 | 0.003 | 3.4E-28 | 313060 | 0.022 | 0.017 | 0.20 | 54162 |
| 20 | rs6050464 | 25209299 | A | C | 0.49 | 0.014 | 0.003 | 1.2E-08 | 312820 | -0.002 | 0.015 | 0.89 | 54162 |
| 20 | rs224424 | 34147998 | A | G | 0.79 | 0.019 | 0.003 | 6.1E-10 | 312879 | 0.004 | 0.018 | 0.82 | 54162 |
| 20 | rs6093446 | 39780932 | A | G | 0.29 | 0.020 | 0.003 | 4.3E-13 | 312712 | 0.006 | 0.017 | 0.73 | 54162 |
| 20 | rs1800961 | 43042364 | C | T | 0.97 | 0.039 | 0.007 | 7.5E-08 | 313060 | -0.060 | 0.047 | 0.20 | 54162 |
| 20 | rs6073958 | 44551855 | C | T | 0.20 | 0.041 | 0.003 | 1.5E-38 | 312629 | 0.027 | 0.021 | 0.19 | 54162 |
| 20 | rs2256814 | 62373983 | A | G | 0.20 | 0.015 | 0.003 | 3.0E-06 | 310674 | 0.044 | 0.022 | 0.04 | 54162 |
| 20 | rs6090101 | 62909520 | A | G | 0.20 | 0.020 | 0.003 | 2.4E-10 | 306925 | -0.042 | 0.085 | 0.62 | 54162 |
| 21 | rs67038483 | 33096103 | T | C | 0.05 | 0.023 | 0.006 | 2.1E-04 | 312991 | 0.090 | 0.037 | 0.01 | 54162 |
| 22 | rs138354 | 41272143 | T | C | 0.47 | 0.013 | 0.003 | 4.5E-07 | 313060 | -0.011 | 0.016 | 0.49 | 54162 |
| 22 | rs9616822 | 50840573 | A | G | 0.35 | 0.015 | 0.003 | 7.0E-09 | 310999 | -0.013 | 0.017 | 0.47 | 54162 |

**Supplementary table 7:** Phenotypic correlations between the lipid traits (LDL-C, HDL-C, triglycerides, ApoA and ApoB).

| | **LDL-C** | **HDL-C** | **Triglycerides** | **ApoA** | **ApoB** |
|---|---|---|---|---|---|
| **LDL-C** | 1.0000 | 0.1016 | 0.2280 | 0.0870 | 0.9586 |
| **HDL-C** | | 1.0000 | -0.4475 | 0.9188 | -0.0320 |
| **Triglycerides** | | | 1.0000 | -0.2858 | 0.2987 |
| **ApoA** | | | | 1.0000 | -0.0305 |
| **ApoB** | | | | | 1.0000 |

**Supplementary figure 1:** Forest plot showing the associations between genetically instrumented lipid traits and dementia risk, using the Kunkle et al. 2019 IGAP GWAS. Estimates are shown for univariable and multivariable MR, using the IWVMR method.

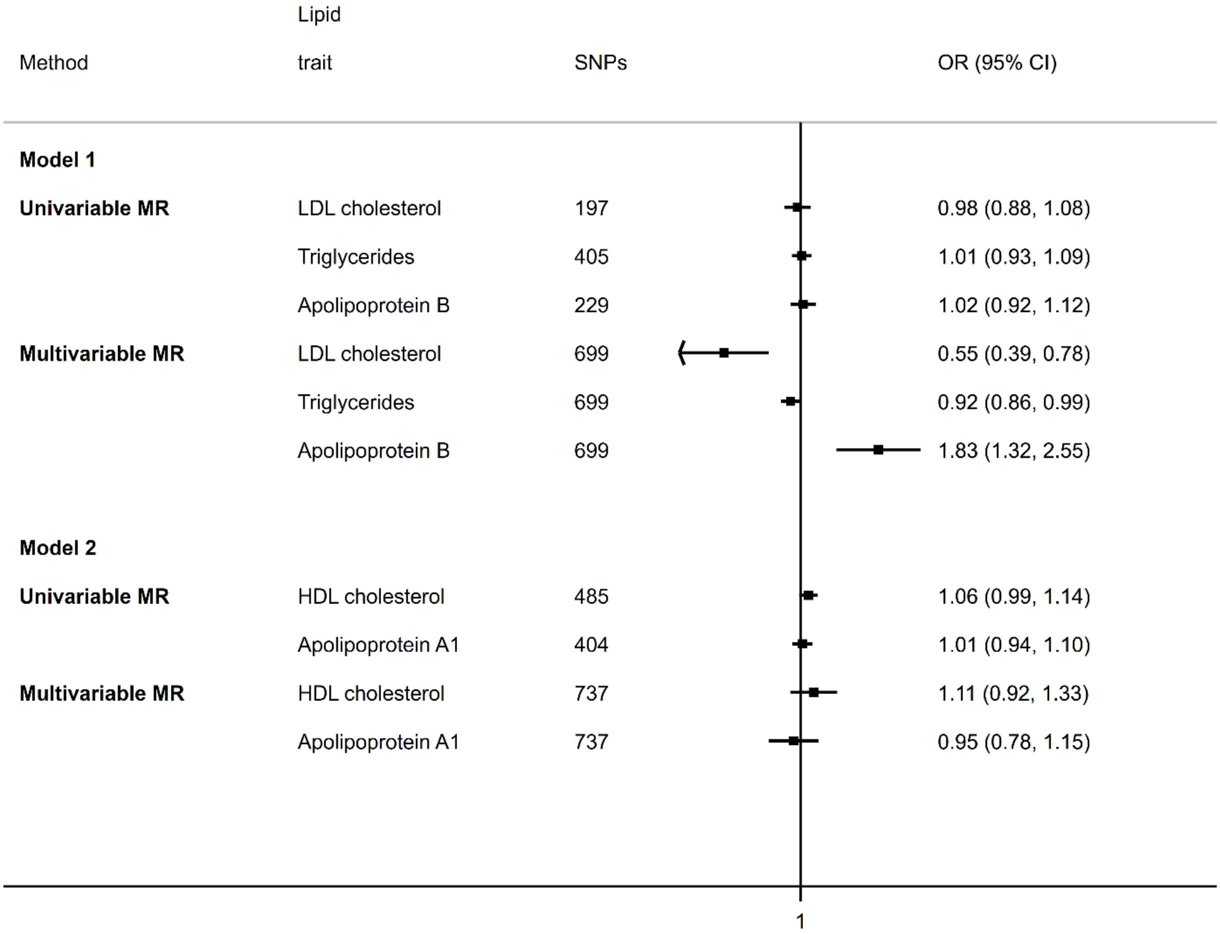

**Supplementary Figure 2:** Forest plot showing the associations between genetically instrumented lipid traits and dementia risk, adjusting for fasting time before blood sample collection or adjusting for history of lipid-lowering medication use. Estimates are shown for univariable and multivariable MR, using the IWVMR method.

| Method | | Lipid trait | SNPs | OR (95% CI) |
|---|---|---|---|---|
| **Fasting time** | | | | |
| **Univariable MR** | **Model 1** | LDL cholesterol | 173 | 1.01 (0.89, 1.15) |
| | | Triglycerides | 361 | 1.00 (0.92, 1.09) |
| | | Apolipoprotein B | 193 | 1.06 (0.94, 1.19) |
| | **Model 2** | HDL cholesterol | 423 | 1.04 (0.95, 1.13) |
| | | Apolipoprotein A1 | 338 | 1.01 (0.92, 1.11) |
| **Multivariable MR** | **Model 1** | LDL cholesterol | 612 | 0.63 (0.43, 0.94) |
| | | Triglycerides | 612 | 0.93 (0.86, 1.02) |
| | | Apolipoprotein B | 612 | 1.63 (1.12, 2.37) |
| | **Model 2** | HDL cholesterol | 635 | 1.20 (0.96, 1.49) |
| | | Apolipoprotein A1 | 635 | 0.87 (0.69, 1.10) |
| **Lipid-lowering medication use** | | | | |
| **Univariable MR** | **Model 1** | LDL cholesterol | 173 | 1.01 (0.91, 1.12) |
| | | Triglycerides | 361 | 1.00 (0.92, 1.09) |
| | | Apolipoprotein B | 193 | 1.04 (0.94, 1.15) |
| | **Model 2** | HDL cholesterol | 423 | 1.04 (0.95, 1.13) |
| | | Apolipoprotein A1 | 338 | 1.01 (0.92, 1.11) |
| **Multivariable MR** | **Model 1** | LDL cholesterol | 612 | 0.62 (0.42, 0.91) |
| | | Triglycerides | 612 | 0.94 (0.86, 1.02) |
| | | Apolipoprotein B | 612 | 1.66 (1.13, 2.43) |
| | **Model 2** | HDL cholesterol | 635 | 1.21 (0.96, 1.52) |
| | | Apolipoprotein A1 | 635 | 0.86 (0.68, 1.10) |

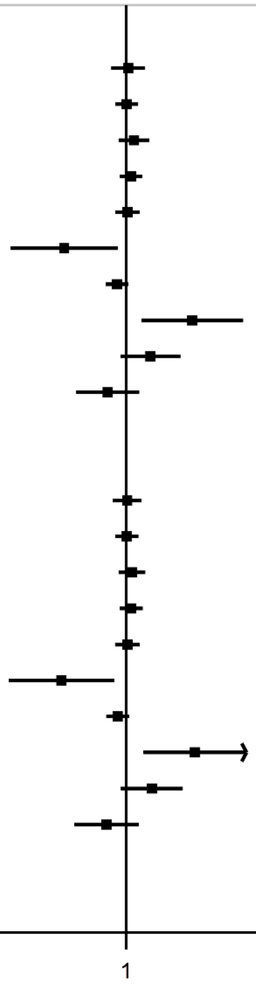

**Supplementary figure 3:** Forest plot showing the associations between genetically instrumented lipid traits and dementia risk. Model 1 uses SNPs associated with LDL-C, triglycerides and ApoB (while controlling for pathways through HDL-C and ApoA) and model 2 uses SNPs associated with HDL-C and ApoA (while controlling for pathways through LDL-C, triglycerides and ApoB). Estimates are shown for multivariable MR using IVWMR, MR-Egger, weighted median, and MR-Lasso.

| Lipid trait | MR Method | SNPs | OR (95% CI) |
|---|---|---|---|
| **Model 1** | | | |
| LDL-cholesterol | IVWMR | 612 | 0.62 (0.42, 0.92) |
| | MR-Egger | 612 | 0.63 (0.42, 0.95) |
| | Weighted median | 612 | 0.84 (0.53, 1.33) |
| | MR-Lasso | 612 | 0.81 (0.56, 1.17) |
| Triglycerides | IVWMR | 612 | 0.93 (0.86, 1.02) |
| | MR-Egger | 612 | 0.94 (0.86, 1.02) |
| | Weighted median | 612 | 0.96 (0.86, 1.07) |
| | MR-Lasso | 612 | 0.97 (0.89, 1.04) |
| Apolipoprotein B | IVWMR | 612 | 1.66 (1.14, 2.42) |
| | MR-Egger | 612 | 1.65 (1.13, 2.41) |
| | Weighted median | 612 | 1.24 (0.81, 1.91) |
| | MR-Lasso | 612 | 1.27 (0.89, 1.80) |
| HDL-cholesterol | IVWMR | 612 | 0.84 (0.69, 1.02) |
| | MR-Egger | 612 | 0.84 (0.69, 1.02) |
| | Weighted median | 612 | 0.94 (0.74, 1.21) |
| | MR-Lasso | 612 | 0.84 (0.71, 1.01) |
| Apolipoprotein A1 | IVWMR | 612 | 1.14 (0.91, 1.41) |
| | MR-Egger | 612 | 1.13 (0.91, 1.41) |
| | Weighted median | 612 | 1.00 (0.75, 1.33) |
| | MR-Lasso | 612 | 1.14 (0.93, 1.40) |
| **Model 2** | | | |
| LDL-cholesterol | IVWMR | 635 | 1.02 (0.67, 1.57) |
| | MR-Egger | 635 | 1.03 (0.67, 1.58) |
| | Weighted median | 635 | 0.90 (0.55, 1.47) |
| | MR-Lasso | 635 | 1.11 (0.75, 1.65) |
| Triglycerides | IVWMR | 635 | 1.02 (0.91, 1.13) |
| | MR-Egger | 635 | 1.02 (0.91, 1.13) |
| | Weighted median | 635 | 0.97 (0.84, 1.11) |
| | MR-Lasso | 635 | 1.00 (0.91, 1.10) |
| Apolipoprotein B | IVWMR | 635 | 0.98 (0.64, 1.49) |
| | MR-Egger | 635 | 0.98 (0.64, 1.48) |
| | Weighted median | 635 | 1.12 (0.70, 1.78) |
| | MR-Lasso | 635 | 0.92 (0.62, 1.35) |
| HDL-cholesterol | IVWMR | 635 | 1.18 (0.94, 1.49) |
| | MR-Egger | 635 | 1.16 (0.92, 1.48) |
| | Weighted median | 635 | 0.98 (0.71, 1.33) |
| | MR-Lasso | 635 | 1.02 (0.83, 1.25) |
| Apolipoprotein A1 | IVWMR | 635 | 0.88 (0.69, 1.12) |
| | MR-Egger | 635 | 0.87 (0.68, 1.11) |
| | Weighted median | 635 | 1.13 (0.81, 1.59) |
| | MR-Lasso | 635 | 1.05 (0.85, 1.31) |

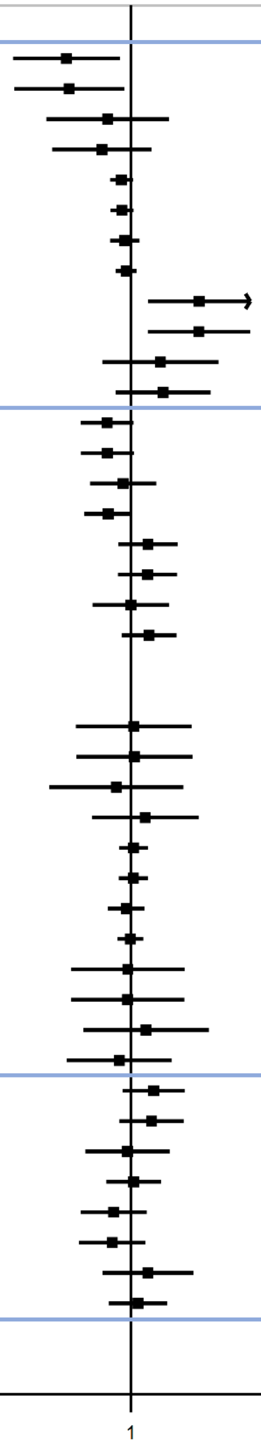

**Supplementary Figure 4:** Forest plot showing the associations between genetically instrumented lipid traits and dementia risk, including SNPs in *APOE* and neighbouring genes (additional SNPs for model 1: rs6857, rs4452060, rs1551891, rs483082, rs12691088; additional SNPs for model 2: rs429358, rs2965169). Estimates are shown for multivariable MR using IVWMR, MR-Egger, weighted median, and MR-Lasso.

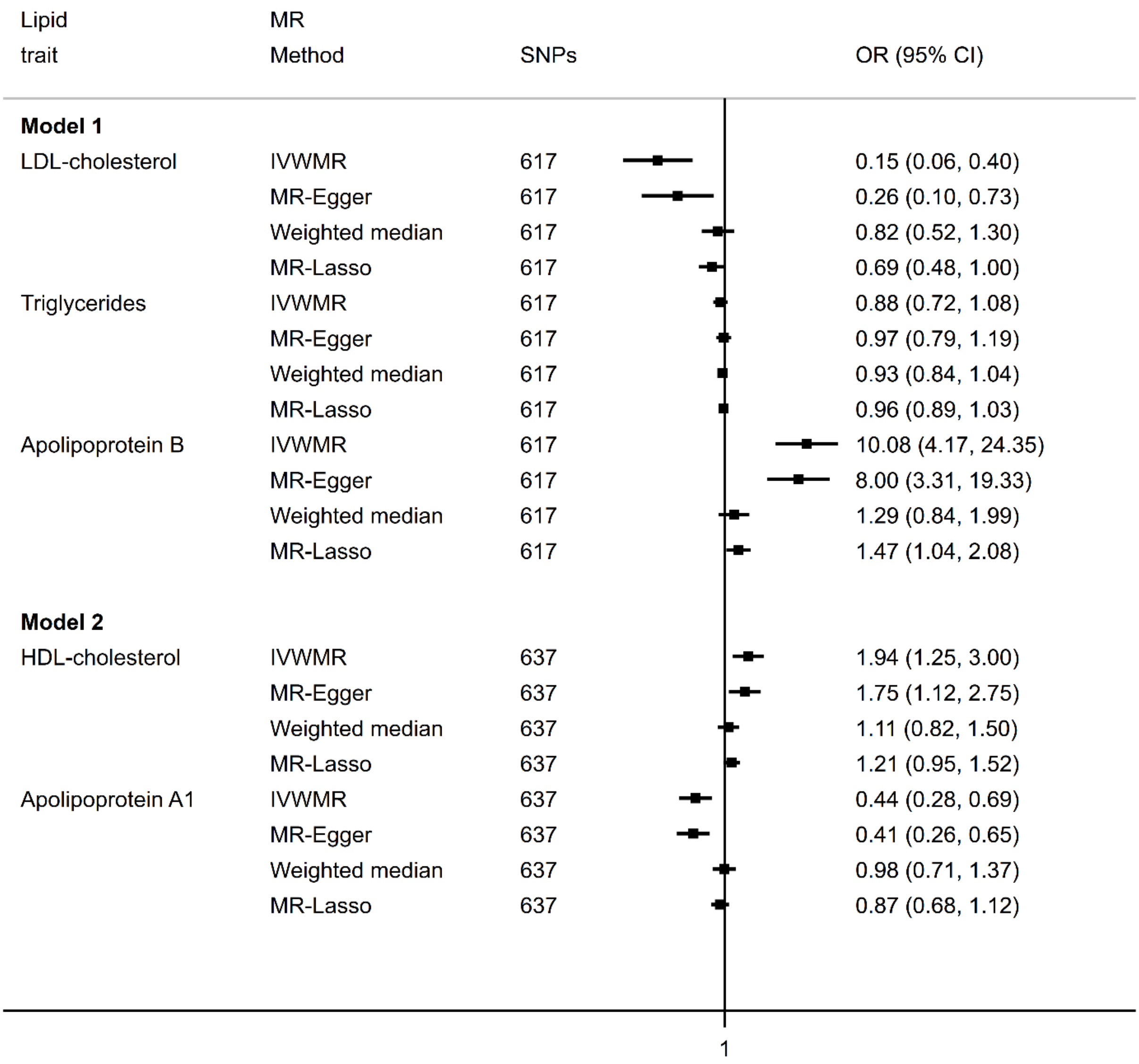
